# Development and Evaluation of Machine Learning Models to Predict Mechanical Restraint and Related Coercive Measures in Hospital Psychiatry

**DOI:** 10.64898/2025.12.15.25342272

**Authors:** Sara Kolding, Jakob Grøhn Damgaard, Martin Bernstorff, Lasse Hansen, Søren Dinesen Østergaard, Andreas Aalkjær Danielsen

**Author notes:** Corresponding author: Sara Kolding, MSc, Department of Affective Disorders, Aarhus University Hospital – Psychiatry, Palle Juul-Jensens Boulevard 175, 8200 Aarhus N, Denmark.

## Abstract

**Introduction:** Use of coercive measures in psychiatric hospitals is clinically and ethically challenging. Aiming to support prevention, we developed and evaluated machine learning models to predict both mechanical restraint and a broader composite outcome that includes related coercive measures.

**Methods:** The dataset comprised electronic health records (EHR) from adults (≥18 years) who had at least one admission to the Psychiatric Services in the Central Denmark Region between 2015 and 2021. For each inpatient day, XGBoost machine learning models were trained to predict mechanical restraint or composite (mechanical, chemical, or manual) restraint within 48 hours. Hyperparameters were optimised for the area under the receiver operating characteristic curve (AUROC) using five-fold cross validation on 85% of the data, with performance validated on a held-out 15% test set.

**Results:** The cohort included 16,834 patients with 45,179 inpatient stays, covering 687,388 prediction days. Of these, 2,736 days were followed by a restraint episode within 48 hours, including 983 episodes of mechanical restraint. The final models were trained on 2,389 EHR-based predictors, derived from demographics, diagnoses, medications, and clinical notes. The mechanical restraint model achieved an AUROC of 0.921 (95% CI: [0.918–0.922]) and a positive predictive value (PPV) of 4.9% when classifying the top 1% of risk scores as positive. The composite model achieved an AUROC of 0.912 (95% CI: [0.909–0.913]) and a PPV of 4.2% when predicting mechanical restraint, and 0.900 (95% CI: [0.898–0.900]) with a PPV of 10.4% when predicting composite restraint.

**Conclusion:** The results indicate that incorporating related coercive measures into model training did not improve discrimination (AUROC) for predicting mechanical restraint but did increase PPV when predicting composite restraint, reflecting the higher outcome prevalence. This suggests that leveraging related outcomes can inform prediction of rare events, emphasising the importance of problem framing in clinical prediction modelling. Future work should include external validation across temporal, geographic, and demographic contexts.

**Significant Outcomes:**

– A machine learning model trained solely for predicting mechanical restraint achieved strong performance (AUROC 0.92), identifying nearly one-third of restraint cases at high specificity.
– Training on a broader composite outcome yielded similar discriminatory performance when predicting mechanical restraint, while the higher base rate resulted in a higher positive predictive value for predicting composite restraint.
– Broadening the outcome to include multiple restraint types increased the number of at-risk patients detected due to the higher prevalence, without compromising accuracy for mechanical restraint, supporting shared underlying risk factors.

**Limitations:**

– The model requires more extensive external validation to assess generalisability across time, demographic groups, and settings, which may be limited by regional/national differences in legislation and clinical documentation.
– Prediction performance was highest near the restraint event, limiting early forecasting and suggesting that limiting predictions to the early phase of hospitalisation, where most restraint occurs, could elevate the base rate and improve model performance.

## 1. Introduction

At psychiatric hospitals, physical coercive measures can be used as a last resort to prevent inpatients from causing harm to themselves or others (1). While the use of such measures varies across countries, most rely on a combination of manual restraint (physical holding by staff), mechanical restraint (fixation to a bed using belts and, in some instances, also cuffs), and chemical restraint (administration of tranquilising medication not intended as treatment of illness) (2, 3).

Restraint practices challenge fundamental ethical principles of patient autonomy (4) and have been associated with numerous adverse outcomes, including cardiovascular events (5), thromboembolisms (5, 6), worsening of psychiatric symptoms (7), and even death (8).

Despite these significant risks, research across multiple countries indicates that restraint is relatively common (9). The proportion of inpatients subjected to mechanical or physical restraint averages 8% in North America and 12% in Europe, with the use of chemical restraint reported to be even more prevalent (10). As a result, it is a common goal to minimise the use of restraint whenever possible (11–14).

One strategy to reduce the use of restraint is to implement early, non-coercive preventive interventions aimed at high-risk patients (15). To identify these high-risk individuals, previous research has demonstrated that prognostic machine learning models trained on electronic health record (EHR) data can predict the use of incident mechanical restraint (16). If sufficiently predictive and clinically validated, such models could serve as clinical decision support tools to identify high-risk patients and prompt timely preventive interventions.

The rarity of the use of mechanical restraint presents a significant challenge for model development, as highly imbalanced datasets can lead to overfitting and poor generalisability (17). However, mechanical restraint shares purpose with other forms of restraint—namely prevention of harm—and is typically also regulated under the same legislation (i.e., subject to the same legal criteria for use) (18). In practice, adherence to the “least intrusive means” principle may lead to use of, e.g., manual or chemical restraint rather than mechanical restraint. However, documentation often reflects only the primary intervention, even when multiple measures are used concurrently (19–21). Given this overlap, it seems reasonable to include related types of restraint when training predictive models. Consequently, we first investigated whether a machine learning model trained on a composite outcome—including manual, chemical, and mechanical restraint—could enhance the prediction of mechanical restraint compared to a model trained solely on the mechanical restraint outcome. Secondarily, we evaluated the performance of the models in predicting composite restraint. The model is intended as a clinical decision support tool to identify high-risk individuals and prompt non-coercive, preventive interventions.

## 2. Materials and Methods

Reporting was in general agreement with the Transparent Reporting of multivariable prediction models for Individual Prognosis Or Diagnosis with Artificial Intelligence (TRIPOD+AI) guidelines (22). The accompanying checklist can be found in Supplementary table 1. The code used to complete the analysis is available at https://github.com/Aarhus-Psychiatry-Research/psycop-common.

### 2.1 Data source and setting

Both training and validation data stem from the PSYchiatric Clinical Outcome Prediction (PSYCOP) cohort (23), which includes EHR data from all individuals in the Central Denmark Region (catchment area serving approximately 1.3 million people across 6 centres) who had at least one contact with Psychiatric Services between January 1, 2011, and November 22, 2021. EHR data is produced as product of routine care, enabling model development without the need for additional data collection. The dataset comprises both structured and unstructured (free-text notes) data from all service contacts—encompassing both psychiatric and general hospital contacts—at public hospitals in the Central Denmark Region. Service contacts include inpatient admissions, outpatient visits, home visits, and consultations by phone (Figure 1A). Data from all patients meeting the eligibility criteria were extracted. Data prior to 2013 were dropped due to instability after the introduction of a new digital EHR-system in 2011 (24, 25). Hence, restricting the cohort to patients with a contact after 2015, ensures that stable data were available for generating predictors based on data from the two years preceding an admission. The end date is November 22, 2021. In line with our prior studies based on this cohort (26–30), formal sample size calculation was not conducted, and the sample size was determined by the available data.

**Figure 1.**
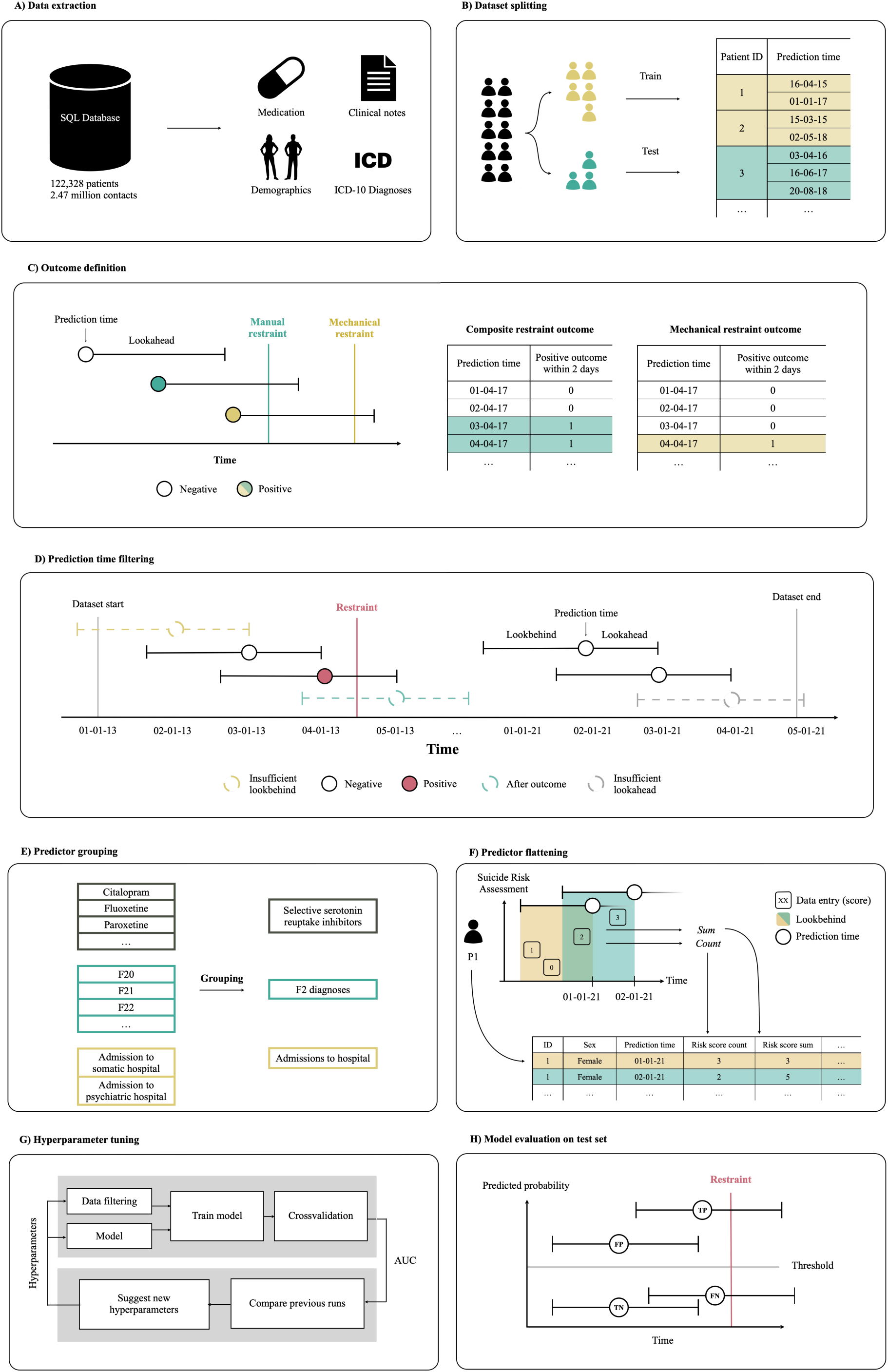
Overview of the study design. (A) Data were extracted from electronic health records. (B) The dataset was randomly divided into independent training (85%) and test (15%) sets, ensuring no patient appeared in both. (C) A prediction time was labelled as a positive outcome if restraint occurred within 48 hours: mechanical restraint for the mechanical model and any manual, chemical, or mechanical restraint for the composite model. (D) Prediction times were excluded if their lookbehind window extended before the start of the dataset, their lookahead extended beyond the dataset end, or if they occurred after the patient had already experienced restraint. (E) Certain predictors were synthesised by renaming, while the number of observations was preserved. (F) For each prediction time, a single value was calculated by aggregating multiple observations within the lookbehind window using various aggregation functions. (G) Models were trained and optimised for the area under the receiver operating characteristic curve using 5-fold cross-validation on the training set. (H) The final model was evaluated on the independent test set, with predictions classified as positive or negative according to the decision threshold. The figure was adapted from Bernstorff et al. (28).

### 2.2 Eligibility criteria

For the present study, the dataset comprises all patients meeting the following criteria:

1. Having at least one psychiatric admission initiated between 1 January 2015 and 20 November 2021.
2. Being ≥ 18 years at the time of admission*.

*As the application of coercive measures to minors is rare and governed by additional legislation, potentially leading to distinct underlying processes and risk factors (31).

### 2.3 Dataset splitting

The data was split into a training and a held-out test set of 85% and 15%, respectively, by randomly sampling unique anonymised patient identifiers into each group (see Figure 1B). Patient assignment to data splits was determined using an a priori stratification based on multiple outcomes defined by the author group, ensuring consistent train–test splits across related studies (26–30). The training set was then further split into five folds for cross validation during training and hyperparameter tuning, stratified by outcome (either mechanical or composite restraint) while still ensuring that data from one individual only occurred in a single fold. The test set was not examined before the final phase of model evaluation, after optimal and final model parameters had been established for all models.

### 2.4 Terminology

To enhance clinical utility, we employed the “landmark model” framework for dynamic prediction (32, 33). This involves selecting specific time points, known as “landmarks” or “prediction times” from which future outcomes are predicted. For consistency, we use the terminology defined in our *timeseriesflattener* Python package throughout, where these time points are defined as prediction times (34). The *lookbehind* window refers to the period preceding the prediction time used for predictor generation, while the *lookahead* window refers to the period succeeding the prediction time, in which we predict whether the outcome occurs. This approach offers several advantages for clinical prediction modelling, as it ensures predictions are made at clinically relevant times, as defined by the authors, and aligns the model’s training, validation, and performance with real-world clinical settings (35).

### 2.5 Outcome definition

For the composite restraint outcome, prediction times for which the patient was either manually, chemically, or mechanically restrained in the following 48 hours were assigned positive outcome labels. For the mechanical restraint outcome, a prediction time was assigned a positive outcome label only when a patient was mechanically restrained in the following 48 hours (see Figure 1C). All prediction times that did not adhere to the criteria for a positive outcome label were assigned negative outcome labels.

### 2.6 Prediction time filtering and exclusion criteria

The model issued predictions every day at 6 a.m., for every calendar day of the inpatient stay following an admission. Prediction times were excluded if the lookbehind (maximum 2 years preceding the prediction time) or lookahead (48 hours) window exceeded the bounds of the dataset (see Figure 1D). Additionally, prediction times following a positive outcome during an inpatient stay are excluded, as the likelihood of repeated restraint is greatly increased, and prediction is hence less informative from a clinical perspective (36, 37). Similarly, all prediction times for inpatient stays where the patient had been restrained in the 365 days preceding the admission start date were excluded. By excluding patients with recent history of restraint, we aimed to identify at-risk patients whose early indicators of heightened aggression or violence are less familiar to hospital staff and, thus, of higher clinical utility.

### 2.7 Predictor engineering

Predictors were derived from several groups of variables selected based on evidence from systematic reviews of risk factors (38–40), findings from nationwide Danish register studies (41), insights from prior predictive models in similar cohorts (16), and clinical experience. These predictors encompassed demographic characteristics, laboratory results, diagnostic codes, and other relevant variables (see Figure 1E).

Various lookbehind windows were used to encode different temporal aspects of the data, with the longest being 2 years. Multiple values within a lookbehind window were condensed to a single summary statistic using specified aggregation functions (e.g., minimum value, mean value, etc.) using the *timeseriesflattener v.2.0.1* Python package (34) (Figure 1F). In instances where no values were present in the records in the specified lookbehind window, a fallback value was imputed instead (e.g., 0 or not a number (NaN)). A comprehensive list of all predictors can be found in Supplementary table 2.

Predictors derived from free text were selected based on the subset of EHR clinical note types considered most informative and stable over time, such as “Subjective Mental State” and “Current Objective Mental State” (see Supplementary table 3 for the full list of note types) (42). The term frequency–inverse document frequency (TF-IDF) statistic was used to create structured predictors from clinical notes, implemented using Scikit-learn (43). For details, see the Supplementary Methods and Supplementary figure 1.

### 2.8 Model training and hyperparameter tuning

For the primary question, two models were trained. Specifically, for each day as an inpatient: i) one model was trained to predict the risk of mechanical restraint in the 48 hours following the prediction, and ii) another model was trained to predict the risk of composite restraint in the 48 hours following the prediction. Both models were then evaluated on an independent dataset, on the task of predicting mechanical restraint in the 48 hours following prediction. As a secondary analysis, the composite restraint model was evaluated on predicting composite restraint.

In addition to the two primary models, a third model was trained using composite restraint outcomes but evaluated out-of-fold on mechanical restraint during five-fold cross-validation. As the composite model was trained and validated solely on the composite outcome, its performance on mechanical restraint reflects an indirect generalization, while the composite-to-mechanical model directly optimizes for mechanical restraint prediction while leveraging the broader composite outcome for training signal. All other aspects of model development, including predictor set, hyperparameter tuning procedures (see below), and evaluation metrics, were kept consistent with the primary models.

To assess whether predictive performance could be maintained with a reduced set of clinically meaningful variables, we trained additional models using a minimal, aetiologically motivated predictor set. These predictors were selected based on clinical expertise and salient predictors in previous work (16), and the models were trained with the same cross-validation procedures and outcome definitions as the primary models. Hyperparameter tuning and performance metrics were kept consistent to enable comparison.

XGBoost models were trained using scikit-learn version 1.2.1 (43). XGBoost was selected due to its strong performance for structured data, fast training capabilities, and inherent ability to handle missing values (44). While we have previously used elastic net regularised logistic regression as a benchmark model for comparison (45, 46), it was not used here, due to results from previous studies suggesting that it is generally inferior to XGBoost for this type of task (27–30). To establish the optimal model parameters, hyperparameter tuning was conducted using the training split to perform 5-fold cross-validation, with outcome stratified folds grouped by anonymous patient identifier (see Figure 1G). See Supplementary Methods and Supplementary table 4 for specifications of hyperparameter tuning.

### 2.9 Model evaluation

The best-performing models according to the area under the receiver operating characteristic curve (AUROC) from hyperparameter tuning, were retrained on the entire training split and evaluated on the held-out test set (see Figure 1H). For sensitivity, specificity, positive predictive value (PPV), and negative predictive value (NPV), predicted probabilities of restraint risk were converted to binary classifications across a range of different predicted positive rates. The predicted positive rate denotes the proportion of the highest probability predictions that are labelled positive. The best predicted positive rate was determined according to F1 score.

We assessed predictor importance by calculating information gain on the best performing model. For XGBoost, information gain reflects the change in predicted probability at a specific node split, averaged across all trees in the model.

### 2.10 Robustness analyses

A series of robustness analyses (across time, demographics, types of restraint, and geographical location of hospitals) were carried out. Please see the Supplementary Methods for a description.

## 3. Results

### 3.1 Dataset

A total of 16,834 patients (median [IQR] age at time of admission, 39.5 [27.7-53.2] years; 8,335 female [49.5%]) were included for analyses, with 45,179 inpatient stays spanning 687,388 days. Of these, 2,736 days were followed by a restraint episode within 48 hours, including 983 mechanical restraint cases. For additional characteristics stratified by data split, see Table 1.

**Table 1.**
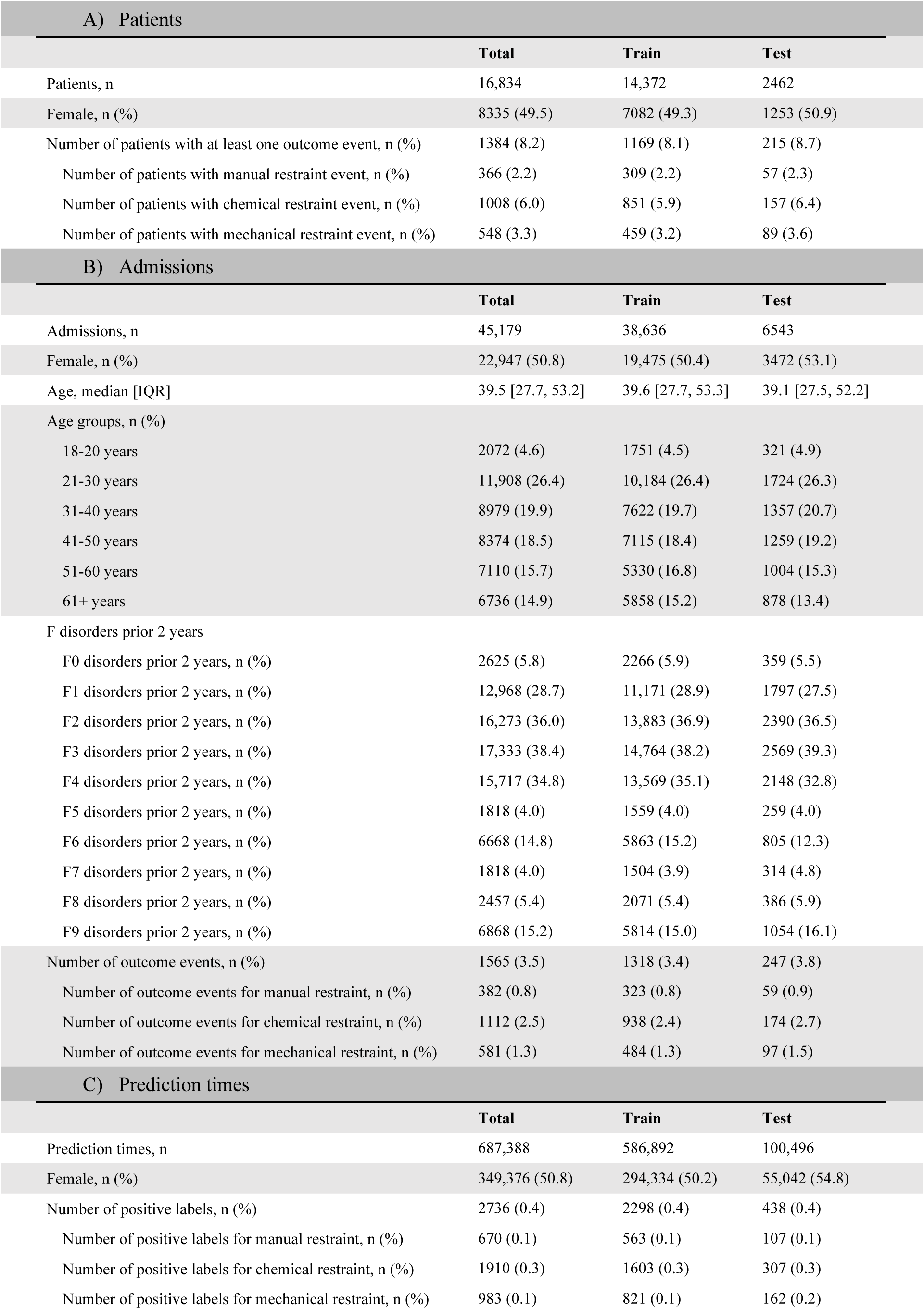
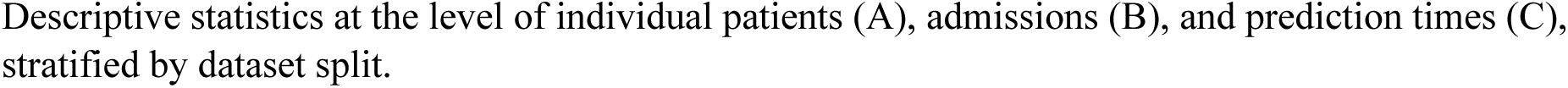
Descriptive cohort statistics.

| A) Patients |  |  |  |
| --- | --- | --- | --- |
|  | Total | Train | Test |
| Patients, n | 16,834 | 14,372 | 2462 |
| Female, n (%) | 8335 (49.5) | 7082 (49.3) | 1253 (50.9) |
| Number of patients with at least one outcome event, n (%) | 1384 (8.2) | 1169 (8.1) | 215 (8.7) |
| Number of patients with manual restraint event, n (%) | 366 (2.2) | 309 (2.2) | 57 (2.3) |
| Number of patients with chemical restraint event, n (%) | 1008 (6.0) | 851 (5.9) | 157 (6.4) |
| Number of patients with mechanical restraint event, n (%) | 548 (3.3) | 459 (3.2) | 89 (3.6) |
| B) Admissions |  |  |  |
|  | Total | Train | Test |
| Admissions, n | 45,179 | 38,636 | 6543 |
| Female, n (%) | 22,947 (50.8) | 19,475 (50.4) | 3472 (53.1) |
| Age, median [IQR] | 39.5 [27.7, 53.2] | 39.6 [27.7, 53.3] | 39.1 [27.5, 52.2] |
| Age groups, n (%) |  |  |  |
| 18-20 years | 2072 (4.6) | 1751 (4.5) | 321 (4.9) |
| 21-30 years | 11,908 (26.4) | 10,184 (26.4) | 1724 (26.3) |
| 31-40 years | 8979 (19.9) | 7622 (19.7) | 1357 (20.7) |
| 41-50 years | 8374 (18.5) | 7115 (18.4) | 1259 (19.2) |
| 51-60 years | 7110 (15.7) | 5330 (16.8) | 1004 (15.3) |
| 61+ years | 6736 (14.9) | 5858 (15.2) | 878 (13.4) |
| F disorders prior 2 years |  |  |  |
| F0 disorders prior 2 years, n (%) | 2625 (5.8) | 2266 (5.9) | 359 (5.5) |
| F1 disorders prior 2 years, n (%) | 12,968 (28.7) | 11,171 (28.9) | 1797 (27.5) |
| F2 disorders prior 2 years, n (%) | 16,273 (36.0) | 13,883 (36.9) | 2390 (36.5) |
| F3 disorders prior 2 years, n (%) | 17,333 (38.4) | 14,764 (38.2) | 2569 (39.3) |
| F4 disorders prior 2 years, n (%) | 15,717 (34.8) | 13,569 (35.1) | 2148 (32.8) |
| F5 disorders prior 2 years, n (%) | 1818 (4.0) | 1559 (4.0) | 259 (4.0) |
| F6 disorders prior 2 years, n (%) | 6668 (14.8) | 5863 (15.2) | 805 (12.3) |
| F7 disorders prior 2 years, n (%) | 1818 (4.0) | 1504 (3.9) | 314 (4.8) |
| F8 disorders prior 2 years, n (%) | 2457 (5.4) | 2071 (5.4) | 386 (5.9) |
| F9 disorders prior 2 years, n (%) | 6868 (15.2) | 5814 (15.0) | 1054 (16.1) |
| Number of outcome events, n (%) | 1565 (3.5) | 1318 (3.4) | 247 (3.8) |
| Number of outcome events for manual restraint, n (%) | 382 (0.8) | 323 (0.8) | 59 (0.9) |
| Number of outcome events for chemical restraint, n (%) | 1112 (2.5) | 938 (2.4) | 174 (2.7) |
| Number of outcome events for mechanical restraint, n (%) | 581 (1.3) | 484 (1.3) | 97 (1.5) |
| C) Prediction times |  |  |  |
|  | Total | Train | Test |
| Prediction times, n | 687,388 | 586,892 | 100,496 |
| Female, n (%) | 349,376 (50.8) | 294,334 (50.2) | 55,042 (54.8) |
| Number of positive labels, n (%) | 2736 (0.4) | 2298 (0.4) | 438 (0.4) |
| Number of positive labels for manual restraint, n (%) | 670 (0.1) | 563 (0.1) | 107 (0.1) |
| Number of positive labels for chemical restraint, n (%) | 1910 (0.3) | 1603 (0.3) | 307 (0.3) |
| Number of positive labels for mechanical restraint, n (%) | 983 (0.1) | 821 (0.1) | 162 (0.2) |

Descriptive statistics at the level of individual patients (A), admissions (B), and prediction times (C), stratified by dataset split.

### 3.2 Model selection

The available predictors included 2,389 predictors collected through standard clinical routine, including 1,639 structured predictors and 750 predictors derived from clinical text. For a comprehensive list of distribution and proportion of missing values for all predictors, see Supplementary table 2. The optimal hyperparameters for each model, determined by AUROC performance during hyperparameter tuning, are listed in Supplementary table 5.

The minimal, aetiologically motivated predictor set contained 17 predictors, including age, sex, status of the admission (voluntary or involuntary), Brøset violence checklist (BVC) scores, past psychiatric disorders, and an admission day count variable (see Supplementary table 2 for full predictor specifications).

### 3.3 Model evaluation

The reported performance estimates are based on the held-out test set. Confidence intervals are estimated from 100 bootstraps. Performance estimates for hyperparameter tuning on the training data can be found in Supplementary table 6.

As seen in Figure 2a, the mechanical restraint model achieved an AUROC of 0.921 (95% CI: [0.918–0.922]) and at a specificity of 99.0%, the sensitivity was 30.2%, the PPV was 4.9%, and the NPV was 99.9% at a predicted positive rate of 1%.

**Figure 2.**
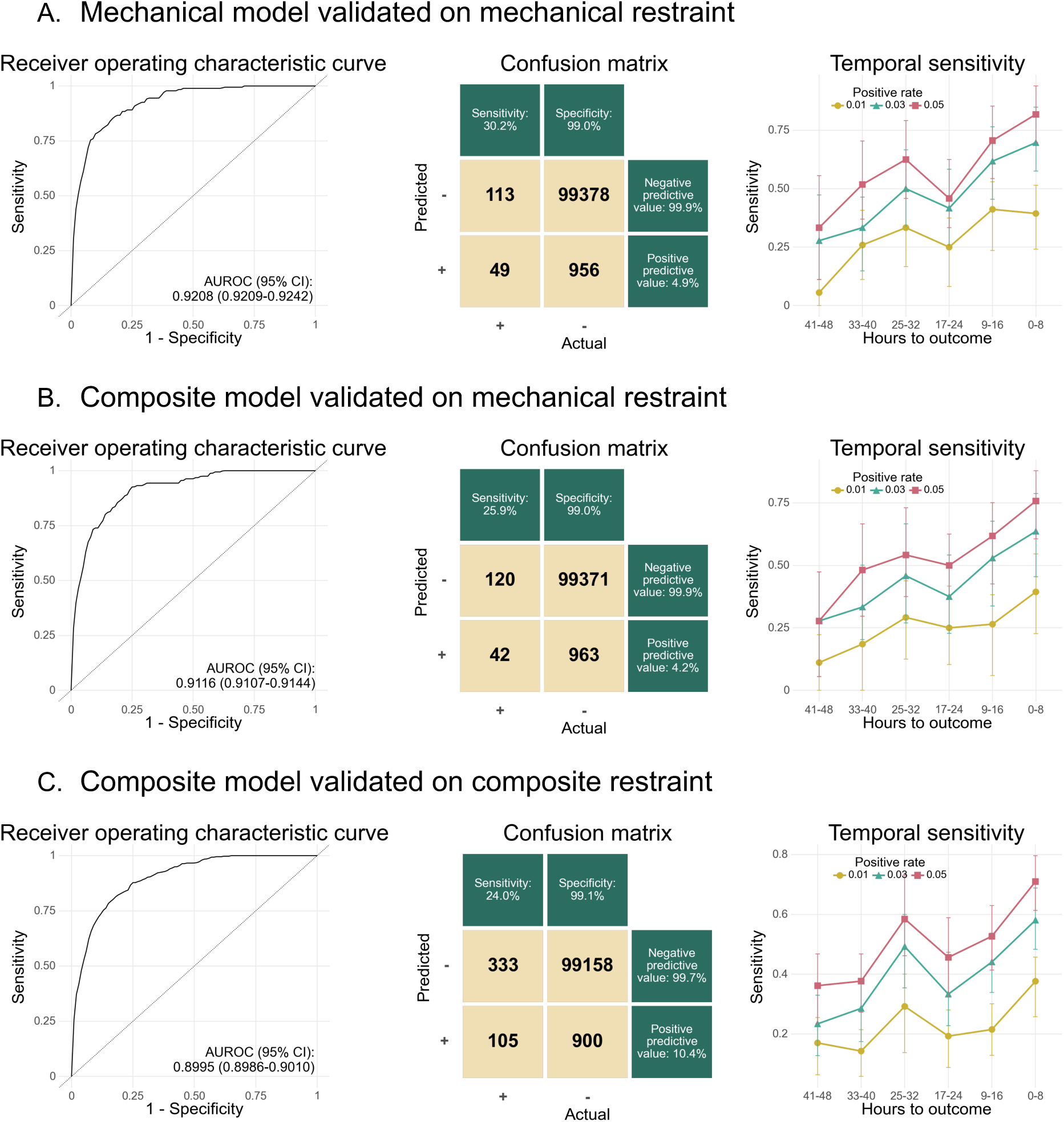
Model performance. Receiver operating characteristic curve of the best performing mechanical restraint model on the test set. Confidence intervals estimated using 100 bootstraps. Confusion matrix with the top 1% predictions classified as positive. Sensitivity by hours from prediction time to outcome event, stratified by positive rates.

The composite restraint model predicted mechanical restraint with an AUROC of 0.912 (95% CI: [0.909–0.913]) and at a specificity of 99.0%, the sensitivity was 25.9%, the PPV was 4.2%, while the NPV was 99.9% at a predicted positive rate of 1% (Figure 2b).

When predicting composite restraint, the composite restraint model achieved an AUROC of 0.900 (95% CI: [0.998–0.900]) and at a specificity of 99.1%, the sensitivity was 24.2%, the PPV was 10.4%, and the NPV was 99.7% at a predicted positive rate of 1% (Figure 2c).

Additionally, for both models and outcomes, model sensitivity increases the closer in time the prediction is made to the outcome (Figure 2). For performance on remaining metrics, see Table 2.

**Table 2.** Model performance metrics.

Mechanical model validated on mechanical restraint
| Predicted positive rate | True prevalence | PPV | NPV | Sens | Spec | FPR | FNR | Acc | TP | TN | FP | FN | FI | MCC | Unique outcome events | Positive outcomes in test set (TP+FN) | Number of unique outcome events detected $\geq 1$ | Prop. of unique outcome events detected $\geq 1$ | Median days from first positive to outcome |
| --- | --- | --- | --- | --- | --- | --- | --- | --- | --- | --- | --- | --- | --- | --- | --- | --- | --- | --- | --- |
| 50.0% | 0.2% | 0.3% | 100.0% | 98.8% | 50.1% | 49.9% | 1.2% | 50.2% | 160 | 50,246 | 50,088 | 2 | 0.6% | 3.9% | 97 | 162 | 96 | 99.0% | 5 |
| 20.0% | 0.2% | 0.7% | 100.0% | 86.4% | 80.1% | 19.9% | 13.6% | 80.1% | 140 | 80,372 | 19,962 | 22 | 1.4% | 6.7% | 97 | 162 | 90 | 92.8% | 5 |
| 10.0% | 0.2% | 1.3% | 100.0% | 77.8% | 90.1% | 9.9% | 22.2% | 90.1% | 126 | 90,410 | 9,924 | 36 | 2.5% | 9.1% | 97 | 162 | 81 | 83.5% | 4 |
| 7.5% | 0.2% | 1.6% | 100.0% | 73.5% | 92.6% | 7.4% | 26.5% | 92.6% | 119 | 92,915 | 7,419 | 43 | 3.1% | 10.1% | 97 | 162 | 79 | 81.4% | 4 |
| 5.0% | 0.2% | 1.9% | 99.9% | 59.9% | 95.1% | 4.9% | 40.1% | 95.0% | 97 | 95,406 | 4,928 | 65 | 3.7% | 10.1% | 97 | 162 | 66 | 68.0% | 4 |
| 4.0% | 0.2% | 2.2% | 99.9% | 53.7% | 96.1% | 3.9% | 46.3% | 96.0% | 87 | 96,401 | 3,933 | 75 | 4.2% | 10.2% | 97 | 162 | 63 | 64.9% | 4 |
| 3.0% | 0.2% | 2.7% | 99.9% | 49.4% | 97.1% | 2.9% | 50.6% | 97.0% | 80 | 97,399 | 2,935 | 82 | 5.0% | 10.9% | 97 | 162 | 59 | 60.8% | 4 |
| 2.0% | 0.2% | 3.4% | 99.9% | 42.0% | 98.1% | 1.9% | 58.0% | 98.0% | 68 | 98,392 | 1,942 | 94 | 6.3% | 11.5% | 97 | 162 | 50 | 51.5% | 4 |
| 1.0% | 0.2% | 4.9% | 99.9% | 30.2% | 99.0% | 1.0% | 69.8% | 98.9% | 49 | 99,378 | 956 | 113 | 8.4% | 11.8% | 97 | 162 | 38 | 39.2% | 2 |

**B. Composite model validated on mechanical restraint**
| Predicted positive rate | True prevalence | PPV | NPV | Sens | Spec | FPR | FNR | Acc | TP | TN | FP | FN | FI | MCC | Unique outcome events | Positive outcomes in test set (TP+FN) | Number of unique outcome events detected $\geq 1$ | Prop. of unique outcome events detected $\geq 1$ | Median days from first positive to outcome |
| --- | --- | --- | --- | --- | --- | --- | --- | --- | --- | --- | --- | --- | --- | --- | --- | --- | --- | --- | --- |
| 50.0% | 0.2% | 0.3% | 100.0% | 96.3% | 50.1% | 49.9% | 3.7% | 50.1% | 156 | 50,241 | 50,093 | 6 | 0.6% | 3.7% | 97 | 162 | 95 | 97.9% | 5 |
| 20.0% | 0.2% | 0.7% | 100.0% | 85.8% | 80.1% | 19.9% | 14.2% | 80.1% | 139 | 80,372 | 19,962 | 23 | 1.4% | 6.6% | 97 | 162 | 87 | 89.7% | 5 |
| 10.0% | 0.2% | 1.2% | 100.0% | 73.5% | 90.1% | 9.9% | 26.5% | 90.1% | 119 | 90,403 | 9,931 | 43 | 2.3% | 8.5% | 97 | 162 | 79 | 81.4% | 4,5 |
| 7.5% | 0.2% | 1.5% | 99.9% | 69.1% | 92.6% | 7.4% | 30.9% | 92.6% | 112 | 92,908 | 7,426 | 50 | 2.9% | 9.4% | 97 | 162 | 77 | 79.4% | 4 |
| 5.0% | 0.2% | 1.8% | 99.9% | 54.9% | 95.1% | 4.9% | 45.1% | 95.0% | 89 | 95,398 | 4,936 | 73 | 3.4% | 9.2% | 97 | 162 | 62 | 63.9% | 4 |
| 4.0% | 0.2% | 2.0% | 99.9% | 50.0% | 96.1% | 3.9% | 50.0% | 96.0% | 81 | 96,395 | 3,939 | 81 | 3.9% | 9.4% | 97 | 162 | 56 | 57.7% | 4 |
| 3.0% | 0.2% | 2.4% | 99.9% | 45.1% | 97.1% | 2.9% | 54.9% | 97.0% | 73 | 97,392 | 2,942 | 89 | 4.6% | 9.9% | 97 | 162 | 50 | 51.5% | 4 |
| 2.0% | 0.2% | 3.1% | 99.9% | 38.9% | 98.1% | 1.9% | 61.1% | 98.0% | 63 | 98,387 | 1,947 | 99 | 5.8% | 10.6% | 97 | 162 | 45 | 46.4% | 3 |
| 1.0% | 0.2% | 4.2% | 99.9% | 25.9% | 99.0% | 1.0% | 74.1% | 98.9% | 42 | 99,371 | 963 | 120 | 7.2% | 10.1% | 97 | 162 | 32 | 33.0% | 4 |

**C. Composite model validated on composite restraint**
| Predicted positive rate | True prevalence | PPV | NPV | Sens | Spec | FPR | FNR | Acc | TP | TN | FP | FN | FI | MCC | Unique outcome events | Positive outcomes (TP+FN) | Number of unique outcome events detected $\geq 1$ | Prop. of unique outcome events detected $\geq 1$ | Median days from first positive to outcome |
| --- | --- | --- | --- | --- | --- | --- | --- | --- | --- | --- | --- | --- | --- | --- | --- | --- | --- | --- | --- |
| 50.0% | 0.4% | 0.8% | 100.0% | 96.6% | 50.2% | 49.8% | 3.4% | 50.4% | 423 | 50,232 | 49,826 | 15 | 1.7% | 6.2% | 236 | 438 | 230 | 97.5% | 5 |
| 20.0% | 0.4% | 1.8% | 99.9% | 82.2% | 80.3% | 19.7% | 17.8% | 80.3% | 360 | 80,317 | 19,741 | 78 | 3.5% | 10.3% | 236 | 438 | 203 | 86.0% | 5 |
| 10.0% | 0.4% | 3.1% | 99.9% | 70.1% | 90.3% | 9.7% | 29.9% | 90.2% | 307 | 90,315 | 9,743 | 131 | 5.9% | 13.3% | 236 | 438 | 180 | 76.3% | 4,5 |
| 7.5% | 0.4% | 3.7% | 99.8% | 63.5% | 92.7% | 7.3% | 36.5% | 92.6% | 278 | 92,798 | 7,260 | 160 | 7.0% | 14.1% | 236 | 438 | 172 | 72.9% | 4 |
| 5.0% | 0.4% | 4.5% | 99.8% | 51.6% | 95.2% | 4.8% | 48.4% | 95.0% | 226 | 95,259 | 4,799 | 212 | 8.3% | 14.1% | 236 | 438 | 144 | 61.0% | 4 |
| 4.0% | 0.4% | 5.1% | 99.8% | 46.8% | 96.2% | 3.8% | 53.2% | 96.0% | 205 | 96,243 | 3,815 | 233 | 9.2% | 14.5% | 236 | 438 | 133 | 56.4% | 4 |
| 3.0% | 0.4% | 6.0% | 99.7% | 41.1% | 97.2% | 2.8% | 58.9% | 96.9% | 180 | 97,223 | 2,835 | 258 | 10.4% | 14.8% | 236 | 438 | 119 | 50.4% | 4 |
| 2.0% | 0.4% | 8.1% | 99.7% | 37.0% | 98.2% | 1.8% | 63.0% | 97.9% | 162 | 98,210 | 1,848 | 276 | 13.2% | 16.5% | 236 | 438 | 107 | 45.3% | 3 |
| 1.0% | 0.4% | 10.4% | 99.7% | 24.0% | 99.1% | 0.9% | 76.0% | 98.8% | 105 | 99,158 | 900 | 333 | 14.6% | 15.3% | 236 | 438 | 78 | 33.1% | 4 |

Predicted positive rate (PPR): the proportion of predicted risk scores that are classified as positive. True prevalence: the proportion of prediction times that followed by an outcome within the lookahead window. AUROC: area under the receiver operator characteristic curve. PPV: positive predictive value. NPV: negative predictive value. FPR: false positive rate. FNR: false negative rate. TP: true positives. Numbers are based on prediction times (end of
psychiatric admission). TN: true negatives. Numbers are based on prediction times (end of psychiatric admission). FP: false positives. Numbers are based on prediction times (end of psychiatric admission). FN: false negatives. Numbers are based on prediction times (end of psychiatric admission). F1: the harmonic mean of the precision and recall. MCC: Matthew's correlation coefficient. Unique outcome events: number of restraint episodes. Positive outcomes: number of positive outcomes (up to two positive outcomes per unique outcome event—true positives + false negatives). Number of unique outcome events detected: number of restraint episodes that are flagged by a least one true positive prediction. Prop. Of unique outcome events detected: proportion of restraint episodes that are flagged by a least one true positive prediction. Median days from first positive to outcome: median number of days from first positive prediction to restraint episode.

Figure 3 shows the 10 most important predictors for each model according to information gain. BVC scores featured heavily in both, in addition to predictors containing data on medication and clinical notes.

**Figure 3.**
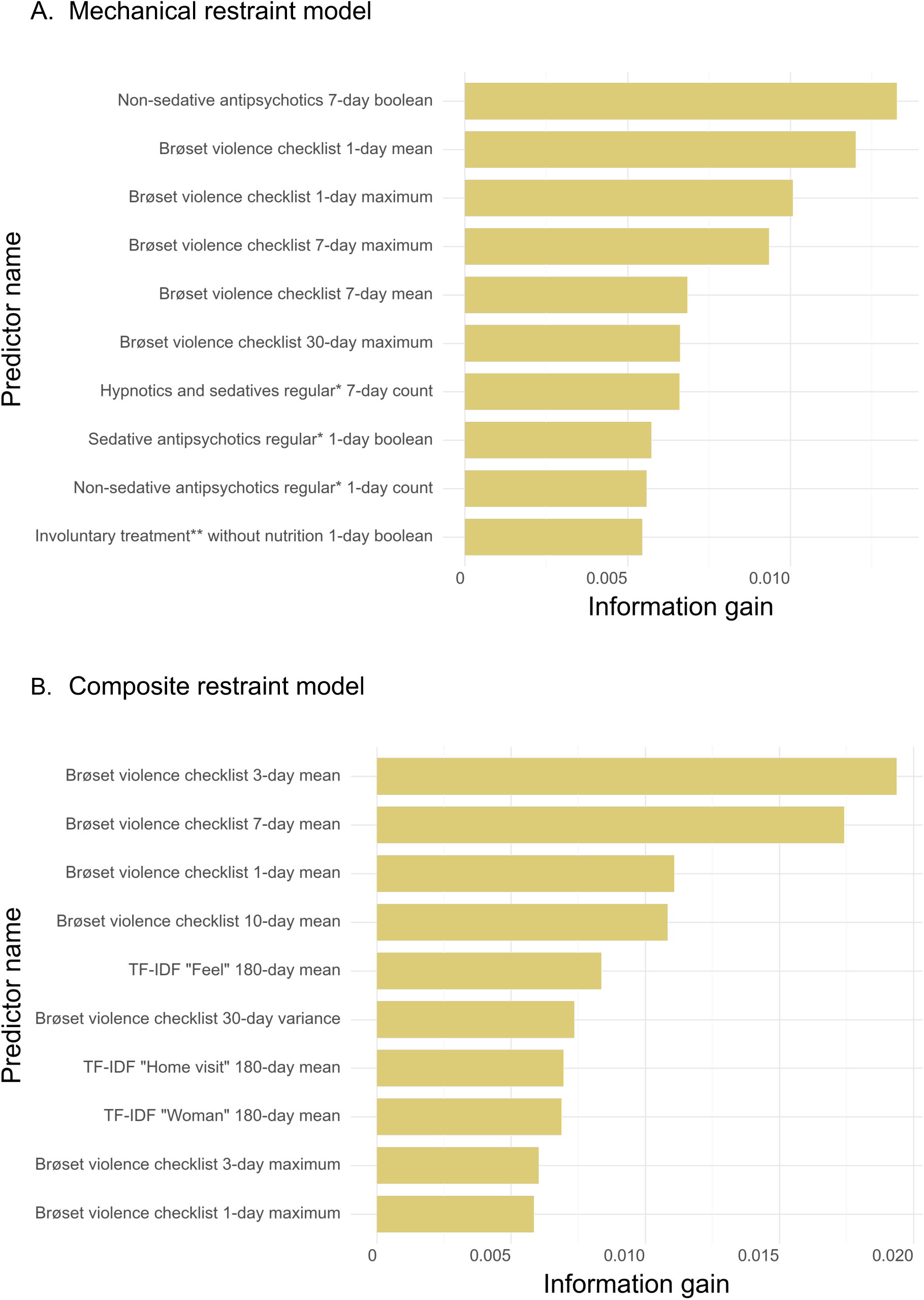
Predictor importance. Predictor importance measured by information gain for the (A) mechanical restraint model and (B) composite restraint model.

For estimates of performance robustness across time, demographics, geographical location of hospitals, and types of restraint, see Supplementary figures 2-6.

The mechanical restraint model trained on a minimal predictor set predicted mechanical restraint with an AUROC of 0.874 (95% CI: [0.874–0.879]), while the composite restraint model trained on the same predictor set obtained an AUROC of 0.876 (95% CI: [0.875–0.880]) when predicting mechanical restraint, and an AUROC of 0.851 (95% CI: [0.850–0.853]) when predicting composite restraint.

The composite-to-mechanical model, trained using composite restraint outcomes but evaluated out-of-fold on mechanical restraint during five-fold cross-validation, predicted mechanical restraint with an AUROC of 0.913 (95% CI: [0.913–0.917]), and at a specificity of 99.0%, the sensitivity was 25.3%, the PPV was 4.1%, and the NPV was 99.9% at a predicted positive rate of 1%.

## 4. Discussion

We examined whether a machine learning model trained on predicting a composite outcome covering manual, chemical, and mechanical restraint (i.e. the composite restraint model), could improve prediction of mechanical restraint, compared to a model trained only to predict mechanical restraint (i.e. the mechanical restraint model). The mechanical restraint model demonstrated strong discriminatory performance, with an AUROC of 0.92. At a high specificity threshold (99.0%), the model identified 30.2% of mechanical restraint incidents with a PPV of 4.9%. The composite restraint model performance slightly decreased when predicting mechanical restraint (AUROC 0.91; specificity 99.9%; sensitivity 25.9%; PPV 4.2%), suggesting that augmentation of the outcome with related restraint types did not improve prediction of mechanical restraint. However, even though the model trained on composite restraint obtained similar discriminatory performance in the prediction of composite restraint (AUROC of 0.90 and sensitivity of 24.2% at a specificity of 99.7%), it had a much higher PPV of 10.4%.

The composite restraint model more than doubled the PPV when predicting composite restraint, compared to its performance on mechanical restraint alone, due to the higher base rate of the broader outcome. Notably, this increase did not appear to be at the expense of prediction of mechanical restraint: the sensitivity of the model was approximately 20% across restraint types (Supplementary figure 6). This indicates that broadening the outcome to include multiple forms of restraint increased the number of at-risk patients detected, without compromising performance on any single restraint type. The same applies to the mechanical restraint model, where some patients flagged as high-risk for mechanical restraint ultimately receive other forms of coercion—appearing as false positives in the evaluation (Supplementary figure 6). This supports the notion that practically and legally related forms of restraint may share a common set of risk factors and underlying mechanisms, and that considering a broader spectrum of coercive interventions offers a more comprehensive understanding of physical coercion in the inpatient psychiatric setting.

Model performance appeared to be primarily driven by BVC-related predictors, as indicated by their high information gain. This was true for both models, while the mechanical restraint model also featured predictors related to use of antipsychotics and sedatives, and the composite restraint model—which included text-based predictors—attributed greater weight to the terms “feel”, “home visit”, and “woman”. These results align with previous findings of BVC scores as strong predictors of restraint in Danish psychiatry (16). Meta-analysis of BVC scores indicate that BVC scores alone are highly predictive of prospective violence (pooled area under the curve (AUC) 0.83 (95% CI 0.78–0.87), 47). Excluding text predictors, the lookbehind periods for top predictors were short, aligning with clinical intuition about recency of information and the temporal dynamics of physical restraint. Since we only created text predictors with 180-days lookbehinds, it is possible that text-based predictors with shorter lookbehind periods would have been equally, or more informative.

Models trained using minimal, aetiologically motivated predictor sets performed worse than models with access to the full predictor set, though they still obtained somewhat high performance with an AUC of 0.87. This indicates that while a limited number of clinically meaningful variables capture a substantial portion of the predictive signal, additional routinely collected data contributes complementary information that enhances model discrimination.

While prior studies have addressed prediction of physical coercion using machine learning, they have primarily focused on specific subpopulations, such as children or adolescents, (48) older patients (49), patients admitted involuntary (50), patients with schizophrenia admitted in forensic settings (51), incident admissions of patients with diagnoses of bipolar disorder, depressive disorder, or schizophrenia (52), or patients at emergency departments (53).

Moreover, the operationalisation of physical coercion differs substantially between these studies, from physical restraint alone (49, 52), to composite outcomes consisting of physical and mechanical restraint, seclusion, and forced medication (51), to unspecified binary coercion outcomes (50). In addition to imprecisely defined outcomes, some studies consider predictors that use data recorded at discharge, thus being incompatible with early detection of at-risk patients (50, 51). Taken together, these aspects complicate meaningful comparison of the results of the present study with those from prior studies.

The present study was based on the approach from Danielsen et al. (16), excluding patients who are frequently subjected to restraint, focusing instead on a more challenging yet clinically valuable task: identifying risk in patients whose need for restraint is likely less apparent to hospital staff (more incident rather than prevalent cases). By including a broad range of inpatients but restricting to those who are not already obvious at-risk cases to clinicians, the model aims to support decision-making where clinical uncertainty is greatest. Additionally, to further enhance practical and clinical utility, the model was developed using only routinely collected clinical data and based on the ‘landmark model’ to help facilitate prospective deployment.

Across all models, sensitivity improved as the prediction time approached the restraint event, highlighting the importance of temporal proximity in forecasting clinical outcomes. This could reflect both a genuine rise in predictive signal as the restraint approaches, or a higher concentration of restraint events in the early days of admission. In practice, this temporal pattern suggests that predictions may be most useful in the initial phase of inpatient stays. Limiting predictions to, for example, the first few days of admission could not only improve efficiency but also help address the extreme class imbalance while still retaining clinical utility, as patients are more likely to be identified and known to staff later in their stay.

Indeed, the low base rate fundamentally constrains PPV and, thus, likely clinical utility. At a high-specificity threshold of 99.7%, only ten in 100 positive predictions were correct, even for the composite restraint outcome. However, this still represents nearly a 25-fold improvement over random chance. Whether this is sufficient for clinical use depends on the trade-off the cost of intervening unnecessarily—as well as the nature of the intervention—and of preventing restraint. A cost-benefit analysis is essential to evaluate whether the clinical value of avoiding restraint outweighs the resource implications of acting on false positives.

Beyond performance metrics, careful consideration must be given to how predictive models are introduced into clinical practice. In the present case, while the intended use would be to allocate preventive resources to patients at risk, there is a theoretical risk that a prediction of “high risk of restraint” could hypothetically be misinterpreted as “high need for restraint” and lead to unlawful restraint. Thus, when implementation of prediction models of this type is considered, it should be preceded/accompanied by teaching of staff with regard to appropriate clinical interpretation of- and action upon model predictions. Specifically, in the present case, a prediction of high risk of restraint for a specific patient should inform allocation of staff resources to reduce the very risk of restraint. Future research should evaluate how such tools affect clinical decisions, patient outcomes, and ward culture in real-world settings. Prospective implementation and cost–benefit studies are needed to investigate their potential to support safer, more patient-centred care.

There are limitations to this study which should be taken into account by readers. First, in addition to potentially limiting clinical utility, the low base rate of restraint events increases the risk of overfitting. However, the stable performance across training and testing, even when predicting mechanical restraint, would suggest that overfitting was minimal. Second, there is need for more extensive external validation to assess the model’s generalisability, particularly across temporally distinct samples. We plan to do so in future work. Third, we did not conduct a fairness audit as we are investigating fairness across a range of prediction models (26–30), including the present one, in an ongoing study. Fairness evaluation is an important next step for model validation and enhancing understanding of model behaviour, as existing literature indicates potential biases in the use of physical and chemical restraint (54). Fourth, while restraint is intended as a last resort for avoiding harm towards self or others, its use is likely mediated by a combination of clinical and contextual factors (55, 56). Indeed, systematic reviews indicate that contextual factors, such as hospital location, ward occupancy, admission rates, and staffing levels, are also associated with the frequency of aggression and restraint events (39, 57). Consequently, predictive models may reflect both clinical risk factors and broader contextual patterns, which may resultantly render predictions imprecise in regard to the individual’s true clinical risk. Recognising this complexity is important when interpreting model outputs and considering their potential role in clinical decision support, to prevent unintended, self-reinforcing feedback loops. Fifth, generalisability across regions or countries may be inherently limited due to differences in patient populations, legislation and clinical practice, e.g., mechanical restraint is not used in some countries (58, 59), while data for certain predictors considered in the present study might not be collected or recorded in others. While predictive performance often drops substantially for models applied to unseen datasets, updating the model with data from the target setting can improve performance (60). Our findings suggest that predicting a broader category of coercive interventions could be useful in other contexts, provided it is adapted to local clinical practice and legislation.

In conclusion, this study demonstrates that machine learning models can predict mechanical restraint with good discriminatory performance in a general psychiatric inpatient population using routinely collected clinical data. Training on a broader composite outcome that included manual, chemical, and mechanical restraint did not improve discrimination for mechanical restraint but did enhance identification of patients at risk of any form of restraint, expanding the pool of at-risk patients identified without loss of performance for any single restraint type. Model performance increased as the predicted restraint event approached, indicating that early prediction remains challenging and that clinical utility may be greatest in the initial phase of admissions. Although the low base rate limits positive predictive value, predictions still performed markedly better than chance and may hold clinical value if coupled with appropriate interventions, as well as careful consideration of limitations and potential challenges.

## Data availability statement

Due to the personally sensitive nature of the data used for this study, it cannot be shared according to Danish law.

## Author contributions

All authors contributed to the design of the study. The data was procured by Østergaard. The analyses were carried out by Kolding, Damgaard, Bernstorff, and Hansen. The results were interpreted by all authors. Kolding drafted the manuscript, which was subsequently revised for important intellectual content by the remaining authors. All authors approved the final version of the manuscript prior to submission.

## Acknowledgements

The study is supported by grants from the Lundbeck Foundation (grant no. R344-2020-1073 to Dr Østergaard), the Danish Cancer Society (grant no. R283-A16461 to Østergaard), the Central Denmark Region Fund for Strengthening of Health Science (grant no. 1-36-72-4-20 to Østergaard), the Danish Agency for Digitisation Investment Fund for New Technologies (grant no. 2020-6720 to Østergaard), the Independent Research Fund Denmark (grant no. 4309-00028B to Østergaard).

The authors are grateful to Professor Ole Mors from the Psychosis Research Unit, Aarhus University Hospital - Psychiatry, for his financial support, valuable input to the study design and research aims, and supervision during the analyses. The authors are grateful to Bettina Nørremark from the Psychiatric Services of the Central Denmark Region for her support with data extraction.

## Ethics statement

The study was approved by the Legal Office of the Central Denmark Region in agreement with the Danish Health Care Act §46, Section 2. The Danish Committee Act exempts studies based solely on EHR data from ethical review board assessment (waiver for this project: 1-10-72-1-22). Handling and storage of data complied with the European Union General Data Protection Regulation. The project is registered on the list of research projects having the Central Denmark Region as data steward. There was no patient nor public involvement in this study.

## Conflicts of interest

Østergaard reported receiving grants from The Lundbeck Foundation (grant No. R358-2020-2341), and Independent Research Fund Denmark (grant Nos. 7016-00048B and 2096-00055A); receiving the 2020 Lundbeck Foundation Young Investigator Prize; owning or having owned units of mutual funds with stock tickers DKIGI, IAIMWC, SPIC25KL and WEKAFKI; and owning or having owned units of exchange traded funds with stock tickers BATE, TRET, QDV5, QDVH, QDVE, SADM, IQQH, USPY, EXH2, 2B76, IS4S, OM3X, EUNL and SXRV outside the submitted work. The other authors report no conflicts of interest.

## Notes

### Competing Interest Statement

Ostergaard reported receiving grants from The Lundbeck Foundation (grant No. R358-2020-2341), and Independent Research Fund Denmark (grant Nos. 7016-00048B and 2096-00055A); receiving the 2020 Lundbeck Foundation Young Investigator Prize; owning or having owned units of mutual funds with stock tickers DKIGI, IAIMWC, SPIC25KL and WEKAFKI; and owning or having owned units of exchange traded funds with stock tickers BATE, TRET, QDV5, QDVH, QDVE, SADM, IQQH, USPY, EXH2, 2B76, IS4S, OM3X, EUNL and SXRV outside the submitted work. The other authors report no conflicts of interest.

### Funding Statement

The study is supported by grants from the Lundbeck Foundation (grant no. R344-2020-1073
to Ostergaard), the Danish Cancer Society (grant no. R283-A16461 to Ostergaard), the Central Denmark Region Fund for Strengthening of Health Science (grant no. 1-36-72-4-20 to Ostergaard), the Danish Agency for Digitisation Investment Fund for New Technologies (grant no. 2020-6720 to Ostergaard), the Independent Research Fund Denmark (grant no. 4309-00028B to Ostergaard).

### Author Declarations

The Central Denmark Region Committees on Health Research Ethics waived ethical approval for this work.

