## Supplementary Material for "Development and Evaluation of Machine Learning Models to Predict Mechanical Restraint and Related Coercive Measures in Hospital Psychiatry"

**Corresponding author**

### Table of Contents

#### SUPPLEMENTARY METHODS 2

**Text predictors** 2

**Hyperparameter tuning** 2

**Robustness analyses** 2

#### SUPPLEMENTARY TABLES AND FIGURES 4

**Supplementary table 1: TRIPOD+AI checklist** 4

**Supplementary table 2: Comprehensive predictor list** 7

**Supplementary table 3: Description of note content by type** 40

**Supplementary table 4: Hyperparameter search space** 42

**Supplementary table 5: Optimal hyperparameters** 43

**Supplementary table 6: Performance estimates for models on the training data** 44

**Supplementary figure 1: Text vectorisation** 45

**Supplementary figure 2: Performance across demographic characteristics** 46

**Supplementary figure 3: Performance across temporal characteristics** 47

**Supplementary figure 4: Performance across geographical location** 48

**Supplementary figure 5: Temporal stability** 49

**Supplementary figure 6: Sensitivity across outcome types** 50

#### REFERENCES 52

### Supplementary Methods

##### Text predictors

For the term frequency-inverse document frequency (TF-IDF) approach, clinical notes from the training data were preprocessed by lowercasing and removing stop words (1), punctuation, symbols, and other non-alphanumerical characters. Unigrams and bigrams were extracted, while the top 10% most frequent n-grams (i.e., terms) were excluded, since these are more common and thus less likely to provide predictive value. From the remaining terms, the 750 most frequent were selected as predictors. For each prediction time, the patient's clinical notes from the preceding 180 days were concatenated into a single document, from which the TF-IDF predictors were constructed. TF-IDF values were calculated for each of the 750 terms; if the term was not present, the predictor was coded as missing (NaN).

##### Hyperparameter tuning

To identify the parameters, we employed the tree structured Parzen estimator algorithm available in Optuna version 2.10.1 to maximise AUROC over 250 model trials, resulting in three distinct, optimal model configurations for the three models (see Supplementary Table 5 for the parameters). All analyses were performed in Python v. 3.11.9.

Data preprocessing steps were also optimised, including imputation, scaling, and predictor reduction. For missing values imputation, the values were estimated from the training data and then applied to both the training and test data.

##### Robustness analyses

To assess stability of model predictions over time, demographic attributes, and individual outcomes (manual restraint, chemical restraint, or mechanical restraint), we performed stratified analyses of AUROC performance on the test set. Geographical stability was investigated by grouping hospitals based on location within the Central Denmark Region: west (including Herning and Holstebro municipalities), east (including Aarhus, Randers, and Horsens municipalities), and central (including Viborg, Skive, and Silkeborg municipalities).

To assess temporal stability, we performed cross-validation on the full dataset using expanding training windows (2016–2020) with validation on data from subsequent years (2016–2021), simulating real-world deployment and prospective performance evaluation.

### Supplementary Tables and Figures

#### Supplementary table 1: TRIPOD+AI checklist

TRIPOD+AI

**AUTHOR NOTE**
We distinguish between two types of omissions:
***N/A:*** Items that are not applicable to the current study.
**-:** Items that could be relevant, but were not performed or reported due to, e.g., prioritisation, scope, or focus.

Version: 11-January-2024

| Section/Topic | Item | Development / evaluation[1] | Checklist item | Reported on page |
|---|---|---|---|---|
| **TITLE** | | | | |
| *Title* | 1 | D;E | Identify the study as developing or evaluating the performance of a multivariable prediction model, the target population, and the outcome to be predicted | 1 |
| **ABSTRACT** | | | | |
| *Abstract* | 2 | D;E | See TRIPOD+AI for Abstracts checklist | |
| **INTRODUCTION** | | | | |
| *Background* | 3a | D;E | Explain the healthcare context (including whether diagnostic or prognostic) and rationale for developing or evaluating the prediction model, including references to existing models | 4 |
| | 3b | D;E | Describe the target population and the intended purpose of the prediction model in the context of the care pathway, including its intended users (e.g., healthcare professionals, patients, public) | 5 |
| | 3c | D;E | Describe any known health inequalities between sociodemographic groups | - (will be addressed in future paper) |
| *Objectives* | 4 | D;E | Specify the study objectives, including whether the study describes the development or validation of a prediction model (or both) | 5 |
| **METHODS** | | | | |
| *Data* | 5a | D;E | Describe the sources of data separately for the development and evaluation datasets (e.g., randomised trial, cohort, routine care or registry data), the rationale for using these data, and representativeness of the data | 6 |
| | 5b | D;E | Specify the dates of the collected participant data, including start and end of participant accrual; and, if applicable, end of follow-up | 6 |
| *Participants* | 6a | D;E | Specify key elements of the study setting (e.g., primary care, secondary care, general population) including the number and location of centres | 6 |
| | 6b | D;E | Describe the eligibility criteria for study participants | 6-7 |
| | 6c | D;E | Give details of any treatments received, and how they were handled during model development or evaluation, if relevant | *N/A* |
| *Data preparation* | 7 | D;E | Describe any data pre-processing and quality checking, including whether this was similar across relevant sociodemographic groups | 8 |
| *Outcome* | 8a | D;E | Clearly define the outcome that is being predicted and the time horizon, including how and when assessed, the rationale for choosing this outcome, and whether the method of outcome assessment is consistent across sociodemographic groups | 7-8 |
| | 8b | D;E | If outcome assessment requires subjective interpretation, describe the qualifications and demographic characteristics of the outcome assessors | *N/A* |
| | 8c | D;E | Report any actions to blind assessment of the outcome to be predicted | *N/A* |
| *Predictors* | 9a | D | Describe the choice of initial predictors (e.g., literature, previous models, all available predictors) and any pre-selection of predictors before model building | 8-9 |
| | 9b | D;E | Clearly define all predictors, including how and when they were measured (and any actions to blind assessment of predictors for the outcome and other predictors) | Supplementary Table 2 |
| | 9c | D;E | If predictor measurement requires subjective interpretation, describe the qualifications and demographic characteristics of the predictor assessors | *N/A* |
| *Sample size* | 10 | D;E | Explain how the study size was arrived at (separately for development and evaluation), and justify that the study size was sufficient to answer the research question. Include details of any sample size calculation | 6 |
| *Missing data* | 11 | D;E | Describe how missing data were handled. Provide reasons for omitting any data | 9 |
| *Analytical methods* | 12a | D | Describe how the data were used (e.g., for development and evaluation of model performance) in the analysis, including whether the data were partitioned, considering any sample size requirements | 7 |
| | 12b | D | Depending on the type of model, describe how predictors were handled in the analyses (functional form, rescaling, transformation, or any standardisation). | Supplementary Table 2 |
| | 12c | D | Specify the type of model, rationale[2], all model-building steps, including any hyperparameter tuning, and method for internal validation | 9-10 |
| | 12d | D;E | Describe if and how any heterogeneity in estimates of model parameter values and model performance was handled and quantified across clusters (e.g., hospitals, countries). See TRIPOD-Cluster for additional considerations[3] | Supplementary Methods |
| | 12e | D;E | Specify all measures and plots used (and their rationale) to evaluate model performance (e.g., discrimination, calibration, clinical utility) and, if relevant, to compare multiple models | 10 |
| | 12f | E | Describe any model updating (e.g., recalibration) arising from the model evaluation, either overall or for particular sociodemographic groups or settings | - |
| | 12g | E | For model evaluation, describe how the model predictions were calculated (e.g., formula, code, object, application programming interface) | 6 |
| *Class imbalance* | 13 | D;E | If class imbalance methods were used, state why and how this was done, and any subsequent methods to recalibrate the model or the model predictions | - |
| *Fairness* | 14 | D;E | Describe any approaches that were used to address model fairness and their rationale | - (will be addressed in future paper) |
| *Model output* | 15 | D | Specify the output of the prediction model (e.g., probabilities, classification). Provide details and rationale for any classification and how the thresholds were identified | 10 |

[1] D=items relevant only to the development of a prediction model; E=items relating solely to the evaluation of a prediction model; D;E=items applicable to both the development and evaluation of a prediction model
[2] Separately for all model building approaches.
[3] TRIPOD-Cluster is a checklist of reporting recommendations for studies developing or validating models that explicitly account for clustering or explore heterogeneity in model performance (eg, at different hospitals or centres). Debray et al, BMJ 2023; 380: e071018 [DOI: 10.1136/bmj-2022-071018]

| | | | | |
|---|---|---|---|---|
| *Training versus evaluation* | 16 | D;E | Identify any differences between the development and evaluation data in healthcare setting, eligibility criteria, outcome, and predictors | 10 |
| *Ethical approval* | 17 | D;E | Name the institutional research board or ethics committee that approved the study and describe the participant-informed consent or the ethics committee waiver of informed consent | 19 |
| **OPEN SCIENCE** | | | | |
| *Funding* | 18a | D;E | Give the source of funding and the role of the funders for the present study | 19 |
| *Conflicts of interest* | 18b | D;E | Declare any conflicts of interest and financial disclosures for all authors | 19-20 |
| *Protocol* | 18c | D;E | Indicate where the study protocol can be accessed or state that a protocol was not prepared | - |
| *Registration* | 18d | D;E | Provide registration information for the study, including register name and registration number, or state that the study was not registered | 19 |
| *Data sharing* | 18e | D;E | Provide details of the availability of the study data | 19 |
| *Code sharing* | 18f | D;E | Provide details of the availability of the analytical code[4] | 6 |
| **PATIENT & PUBLIC INVOLVEMENT** | | | | |
| *Patient & Public Involvement* | 19 | D;E | Provide details of any patient and public involvement during the design, conduct, reporting, interpretation, or dissemination of the study or state no involvement. | 19 |
| **RESULTS** | | | | |
| *Participants* | 20a | D;E | Describe the flow of participants through the study, including the number of participants with and without the outcome and, if applicable, a summary of the follow-up time. A diagram may be helpful. | - |
| | 20b | D;E | Report the characteristics overall and, where applicable, for each data source or setting, including the key dates, key predictors (including demographics), treatments received, sample size, number of outcome events, follow-up time, and amount of missing data. A table may be helpful. Report any differences across key demographic groups. | - |
| | 20c | E | For model evaluation, show a comparison with the development data of the distribution of important predictors (demographics, predictors, and outcome). | - |
| *Model development* | 21 | D;E | Specify the number of participants and outcome events in each analysis (e.g., for model development, hyperparameter tuning, model evaluation) | 12 |
| *Model specification* | 22 | D | Provide details of the full prediction model (e.g., formula, code, object, application programming interface) to allow predictions in new individuals and to enable third-party evaluation and implementation, including any restrictions to access or re-use (e.g., freely available, proprietary)[5] | - |
| *Model performance* | 23a | D;E | Report model performance estimates with confidence intervals, including for any key subgroups (e.g., sociodemographic). Consider plots to aid presentation. | 12-13 |
| | 23b | D;E | If examined, report results of any heterogeneity in model performance across clusters. See TRIPOD Cluster for additional details[3]. | Supplementary figures 2-6 |
| *Model updating* | 24 | E | Report the results from any model updating, including the updated model and subsequent performance | - |
| **DISCUSSION** | | | | |
| *Interpretation* | 25 | D;E | Give an overall interpretation of the main results, including issues of fairness in the context of the objectives and previous studies | 14 |
| *Limitations* | 26 | D;E | Discuss any limitations of the study (such as a non-representative sample, sample size, overfitting, missing data) and their effects on any biases, statistical uncertainty, and generalizability | 17 |
| *Usability of the model in the context of current care* | 27a | D | Describe how poor quality or unavailable input data (e.g., predictor values) should be assessed and handled when implementing the prediction model | - |
| | 27b | D | Specify whether users will be required to interact in the handling of the input data or use of the model, and what level of expertise is required of users | - |
| | 27c | D;E | Discuss any next steps for future research, with a specific view to applicability and generalizability of the model | 16 |

From: Collins GS, Moons KGM, Dhiman P, et al. *BMJ* 2024;385:e078378. doi:10.1136/bmj-2023-078378

[4] This relates to the analysis code, for example, any data cleaning, feature engineering, model building, evaluation.
[5] This relates to the code to implement the model to get estimates of risk for a new individual.

**Supplementary table 2: Comprehensive predictor list**

| Feature name | Lookbehind period | Resolve multiple strategy | Fallback strategy | Static | Mean | Number of unique values | Proportion using fallback | Mechanical model | Composite model | Minimal models |
|---|---|---|---|---|---|---|---|---|---|---|
| adm_day_count | - | - | - | FALSE | 51.897 | 1796 | 0 | X | X | X |
| admissions | 365 | count | 0 | FALSE | 3.018 | 99 | 0.274 | | | |
| admissions | 365 | boolean | 0 | FALSE | 0.726 | 2 | 0.274 | | X | |
| admissions | 365 | summed | 0 | FALSE | 3.018 | 99 | 0.274 | | | |
| admissions | 730 | boolean | 0 | FALSE | 0.806 | 2 | 0.194 | X | X | |
| admissions | 730 | summed | 0 | FALSE | 5.034 | 149 | 0.194 | X | | |
| admissions | 730 | count | 0 | FALSE | 5.034 | 149 | 0.194 | X | | |
| admissions | 10 | summed | 0 | FALSE | 0.169 | 12 | 0.865 | | X | |
| admissions | 10 | count | 0 | FALSE | 0.169 | 12 | 0.865 | | X | |
| admissions | 10 | boolean | 0 | FALSE | 0.135 | 2 | 0.865 | | X | |
| admissions | 30 | summed | 0 | FALSE | 0.455 | 23 | 0.7 | X | X | |
| admissions | 30 | boolean | 0 | FALSE | 0.3 | 2 | 0.7 | X | | |
| admissions | 30 | count | 0 | FALSE | 0.455 | 23 | 0.7 | X | X | |
| admissions | 180 | boolean | 0 | FALSE | 0.63 | 2 | 0.37 | | X | |
| admissions | 180 | count | 0 | FALSE | 1.783 | 63 | 0.37 | | | |
| admissions | 180 | summed | 0 | FALSE | 1.783 | 63 | 0.37 | | | |
| admissions_to_psychiatry | 180 | boolean | 0 | FALSE | 0.523 | 2 | 0.477 | | X | |
| admissions_to_psychiatry | 180 | summed | 0 | FALSE | 1.202 | 41 | 0.477 | | | |
| admissions_to_psychiatry | 180 | count | 0 | FALSE | 1.202 | 41 | 0.477 | | | |
| admissions_to_psychiatry | 365 | boolean | 0 | FALSE | 0.62 | 2 | 0.38 | | X | |
| admissions_to_psychiatry | 365 | summed | 0 | FALSE | 2.069 | 50 | 0.38 | | | |
| admissions_to_psychiatry | 365 | count | 0 | FALSE | 2.069 | 50 | 0.38 | | | |
| admissions_to_psychiatry | 730 | boolean | 0 | FALSE | 0.698 | 2 | 0.302 | X | X | |
| admissions_to_psychiatry | 730 | count | 0 | FALSE | 3.474 | 92 | 0.302 | X | | |
| admissions_to_psychiatry | 730 | summed | 0 | FALSE | 3.474 | 92 | 0.302 | X | | |
| admissions_to_psychiatry | 10 | boolean | 0 | FALSE | 0.088 | 2 | 0.912 | | X | |
| admissions_to_psychiatry | 10 | summed | 0 | FALSE | 0.097 | 7 | 0.912 | | X | |
| admissions_to_psychiatry | 10 | count | 0 | FALSE | 0.097 | 7 | 0.912 | | X | |
| admissions_to_psychiatry | 30 | boolean | 0 | FALSE | 0.22 | 2 | 0.78 | X | | |
| admissions_to_psychiatry | 30 | count | 0 | FALSE | 0.282 | 14 | 0.78 | X | | |
| admissions_to_psychiatry | 30 | summed | 0 | FALSE | 0.282 | 14 | 0.78 | X | | |
| admissions_to_somatic | 180 | boolean | 0 | FALSE | 0.269 | 2 | 0.731 | | | |
| admissions_to_somatic | 180 | count | 0 | FALSE | 0.582 | 55 | 0.731 | | | |
| admissions_to_somatic | 180 | summed | 0 | FALSE | 0.582 | 55 | 0.731 | | | |
| admissions_to_somatic | 365 | summed | 0 | FALSE | 0.949 | 79 | 0.652 | | | |
| admissions_to_somatic | 365 | count | 0 | FALSE | 0.949 | 79 | 0.652 | | | |
| admissions_to_somatic | 365 | boolean | 0 | FALSE | 0.348 | 2 | 0.652 | | | |
| admissions_to_somatic | 730 | count | 0 | FALSE | 1.561 | 117 | 0.549 | X | | |
| admissions_to_somatic | 730 | summed | 0 | FALSE | 1.561 | 117 | 0.549 | X | | |
| admissions_to_somatic | 730 | boolean | 0 | FALSE | 0.451 | 2 | 0.549 | X | | |
| admissions_to_somatic | 10 | summed | 0 | FALSE | 0.072 | 10 | 0.94 | | X | |
| admissions_to_somatic | 10 | count | 0 | FALSE | 0.072 | 10 | 0.94 | | X | |
| admissions_to_somatic | 10 | boolean | 0 | FALSE | 0.06 | 2 | 0.94 | | X | |
| admissions_to_somatic | 30 | boolean | 0 | FALSE | 0.122 | 2 | 0.878 | X | | |
| admissions_to_somatic | 30 | count | 0 | FALSE | 0.173 | 21 | 0.878 | X | X | |
| admissions_to_somatic | 30 | summed | 0 | FALSE | 0.173 | 21 | 0.878 | X | X | |
| af_helbredsmaessige_grunde | 10 | count | 0 | FALSE | 0.015 | 3 | 0.985 | | | |
| af_helbredsmaessige_grunde | 10 | boolean | 0 | FALSE | 0.015 | 2 | 0.985 | | | |
| af_helbredsmaessige_grunde | 10 | summed | 0 | FALSE | 6.164 | 131 | 0.985 | | | |
| af_helbredsmaessige_grunde | 30 | summed | 0 | FALSE | 12.355 | 132 | 0.971 | X | | |
| af_helbredsmaessige_grunde | 30 | count | 0 | FALSE | 0.029 | 4 | 0.971 | X | | |
| af_helbredsmaessige_grunde | 30 | boolean | 0 | FALSE | 0.029 | 2 | 0.971 | X | | |
| af_helbredsmaessige_grunde | 180 | boolean | 0 | FALSE | 0.06 | 2 | 0.94 | | | |
| af_helbredsmaessige_grunde | 180 | summed | 0 | FALSE | 36.412 | 156 | 0.94 | | | |
| af_helbredsmaessige_grunde | 180 | count | 0 | FALSE | 0.069 | 6 | 0.94 | | | |
| af_helbredsmaessige_grunde | 365 | summed | 0 | FALSE | 57.571 | 189 | 0.921 | | X | |
| af_helbredsmaessige_grunde | 365 | boolean | 0 | FALSE | 0.079 | 2 | 0.921 | | X | |
| af_helbredsmaessige_grunde | 365 | count | 0 | FALSE | 0.099 | 7 | 0.921 | | X | |
| af_helbredsmaessige_grunde | 730 | boolean | 0 | FALSE | 0.115 | 2 | 0.885 | X | X | |
| af_helbredsmaessige_grunde | 730 | count | 0 | FALSE | 0.163 | 10 | 0.885 | X | X | |
| af_helbredsmaessige_grunde | 730 | summed | 0 | FALSE | 110.64 | 246 | 0.885 | X | X | |
| af_helbredsmaessige_grunde | 1 | boolean | 0 | FALSE | 0.064 | 2 | 0.936 | X | X | |
| af_helbredsmaessige_grunde | 3 | boolean | 0 | FALSE | 0.067 | 2 | 0.933 | | X | |
| af_helbredsmaessige_grunde | 7 | boolean | 0 | FALSE | 0.011 | 2 | 0.989 | X | | |
| af_helbredsmaessige_grunde | 7 | count | 0 | FALSE | 0.011 | 3 | 0.989 | X | | |
| af_helbredsmaessige_grunde | 7 | summed | 0 | FALSE | 4.878 | 131 | 0.989 | X | | |
| af_legemlig_lidelse | 7 | boolean | 0 | FALSE | 0.001 | 2 | 0.999 | X | X | |
| af_legemlig_lidelse | 7 | count | 0 | FALSE | 0.001 | 5 | 0.999 | X | X | |
| af_legemlig_lidelse | 7 | summed | 0 | FALSE | 0.179 | 26 | 0.999 | X | | |
| af_legemlig_lidelse | 10 | boolean | 0 | FALSE | 0.001 | 2 | 0.999 | | X | |
| af_legemlig_lidelse | 10 | count | 0 | FALSE | 0.001 | 5 | 0.999 | | X | |
| af_legemlig_lidelse | 10 | summed | 0 | FALSE | 0.241 | 26 | 0.999 | | | |
| af_legemlig_lidelse | 30 | boolean | 0 | FALSE | 0.002 | 2 | 0.998 | X | | |

| | | | | | | | | | | |
|---|---|---|---|---|---|---|---|---|---|---|
| af_legemlig_lidelse | 30 | count | 0 | FALSE | 0.002 | 7 | 0.998 | X | | |
| af_legemlig_lidelse | 30 | summed | 0 | FALSE | 0.584 | 27 | 0.998 | X | | |
| af_legemlig_lidelse | 180 | summed | 0 | FALSE | 1.124 | 32 | 0.996 | | | |
| af_legemlig_lidelse | 180 | boolean | 0 | FALSE | 0.004 | 2 | 0.996 | | | |
| af_legemlig_lidelse | 180 | count | 0 | FALSE | 0.005 | 9 | 0.996 | | | |
| af_legemlig_lidelse | 365 | boolean | 0 | FALSE | 0.005 | 2 | 0.995 | | | |
| af_legemlig_lidelse | 365 | count | 0 | FALSE | 0.007 | 9 | 0.995 | | | |
| af_legemlig_lidelse | 365 | summed | 0 | FALSE | 1.656 | 33 | 0.995 | | | |
| af_legemlig_lidelse | 730 | boolean | 0 | FALSE | 0.009 | 2 | 0.991 | X | | |
| af_legemlig_lidelse | 730 | summed | 0 | FALSE | 2.964 | 40 | 0.991 | X | | |
| af_legemlig_lidelse | 730 | count | 0 | FALSE | 0.013 | 9 | 0.991 | X | | |
| af_legemlig_lidelse | 1 | boolean | 0 | FALSE | 0.002 | 2 | 0.998 | X | X | |
| af_legemlig_lidelse | 3 | boolean | 0 | FALSE | 0.002 | 2 | 0.998 | | X | |
| age_in_years | - | - | - | FALSE | 42.727 | 7429 | 0 | X | X | X |
| alcohol_abstinence | 1 | boolean | 0 | FALSE | 0.1 | 2 | 0.9 | X | | |
| alcohol_abstinence | 1 | count | 0 | FALSE | 0.201 | 18 | 0.9 | X | | |
| alcohol_abstinence | 3 | boolean | 0 | FALSE | 0.104 | 2 | 0.896 | | | |
| alcohol_abstinence | 3 | count | 0 | FALSE | 0.553 | 40 | 0.896 | | | |
| alcohol_abstinence | 7 | boolean | 0 | FALSE | 0.108 | 2 | 0.892 | X | | |
| alcohol_abstinence | 7 | count | 0 | FALSE | 1.146 | 68 | 0.892 | X | | |
| alcohol_abstinence | 10 | boolean | 0 | FALSE | 0.111 | 2 | 0.889 | | | |
| alcohol_abstinence | 10 | count | 0 | FALSE | 1.529 | 82 | 0.889 | | | |
| alcohol_abstinence | 30 | boolean | 0 | FALSE | 0.122 | 2 | 0.878 | X | | |
| alcohol_abstinence | 30 | count | 0 | FALSE | 3.326 | 167 | 0.878 | X | | |
| alcohol_abstinence | 180 | boolean | 0 | FALSE | 0.151 | 2 | 0.849 | | | |
| alcohol_abstinence | 180 | count | 0 | FALSE | 8.516 | 590 | 0.849 | | | |
| alcohol_abstinence | 365 | boolean | 0 | FALSE | 0.167 | 2 | 0.833 | | | |
| alcohol_abstinence | 365 | count | 0 | FALSE | 12.122 | 985 | 0.833 | | | |
| alcohol_abstinence | 730 | boolean | 0 | FALSE | 0.191 | 2 | 0.809 | X | | |
| alcohol_abstinence | 730 | count | 0 | FALSE | 16.793 | 1295 | 0.809 | X | | |
| ambulatory_visits | 180 | summed | 0 | FALSE | 6.215 | 183 | 0.257 | | X | |
| ambulatory_visits | 180 | boolean | 0 | FALSE | 0.743 | 2 | 0.257 | | X | |
| ambulatory_visits | 180 | count | 0 | FALSE | 6.215 | 183 | 0.257 | | X | |
| ambulatory_visits | 365 | summed | 0 | FALSE | 11.758 | 329 | 0.174 | | X | |
| ambulatory_visits | 365 | count | 0 | FALSE | 11.758 | 329 | 0.174 | | X | |
| ambulatory_visits | 365 | boolean | 0 | FALSE | 0.826 | 2 | 0.174 | | X | |
| ambulatory_visits | 730 | count | 0 | FALSE | 21.019 | 523 | 0.112 | X | X | |
| ambulatory_visits | 730 | summed | 0 | FALSE | 21.019 | 523 | 0.112 | X | X | |
| ambulatory_visits | 730 | boolean | 0 | FALSE | 0.888 | 2 | 0.112 | X | X | |
| ambulatory_visits | 10 | count | 0 | FALSE | 0.291 | 22 | 0.81 | | | |
| ambulatory_visits | 10 | boolean | 0 | FALSE | 0.19 | 2 | 0.81 | | | |
| ambulatory_visits | 10 | summed | 0 | FALSE | 0.291 | 22 | 0.81 | | | |
| ambulatory_visits | 30 | boolean | 0 | FALSE | 0.408 | 2 | 0.592 | X | X | |
| ambulatory_visits | 30 | summed | 0 | FALSE | 1.002 | 58 | 0.592 | X | | |
| ambulatory_visits | 30 | count | 0 | FALSE | 1.002 | 58 | 0.592 | X | | |
| ambulatory_visits_to_psychiatry | 180 | count | 0 | FALSE | 5.133 | 173 | 0.387 | | X | |
| ambulatory_visits_to_psychiatry | 180 | summed | 0 | FALSE | 5.133 | 173 | 0.387 | | X | |
| ambulatory_visits_to_psychiatry | 180 | boolean | 0 | FALSE | 0.613 | 2 | 0.387 | | X | |
| ambulatory_visits_to_psychiatry | 365 | boolean | 0 | FALSE | 0.692 | 2 | 0.308 | | X | |
| ambulatory_visits_to_psychiatry | 365 | count | 0 | FALSE | 9.705 | 314 | 0.308 | | X | |
| ambulatory_visits_to_psychiatry | 365 | summed | 0 | FALSE | 9.705 | 314 | 0.308 | | X | |
| ambulatory_visits_to_psychiatry | 730 | boolean | 0 | FALSE | 0.757 | 2 | 0.243 | X | X | |
| ambulatory_visits_to_psychiatry | 730 | count | 0 | FALSE | 17.324 | 488 | 0.243 | X | | |
| ambulatory_visits_to_psychiatry | 730 | summed | 0 | FALSE | 17.324 | 488 | 0.243 | X | | |
| ambulatory_visits_to_psychiatry | 10 | count | 0 | FALSE | 0.224 | 22 | 0.856 | | | |
| ambulatory_visits_to_psychiatry | 10 | summed | 0 | FALSE | 0.224 | 22 | 0.856 | | | |
| ambulatory_visits_to_psychiatry | 10 | boolean | 0 | FALSE | 0.144 | 2 | 0.856 | | | |
| ambulatory_visits_to_psychiatry | 30 | count | 0 | FALSE | 0.806 | 57 | 0.677 | X | | |
| ambulatory_visits_to_psychiatry | 30 | boolean | 0 | FALSE | 0.323 | 2 | 0.677 | X | X | |
| ambulatory_visits_to_psychiatry | 30 | summed | 0 | FALSE | 0.806 | 57 | 0.677 | X | | |
| ambulatory_visits_to_somatic | 180 | boolean | 0 | FALSE | 0.342 | 2 | 0.658 | | | |
| ambulatory_visits_to_somatic | 180 | summed | 0 | FALSE | 1.082 | 82 | 0.658 | | | |
| ambulatory_visits_to_somatic | 180 | count | 0 | FALSE | 1.082 | 82 | 0.658 | | | |
| ambulatory_visits_to_somatic | 365 | count | 0 | FALSE | 2.053 | 125 | 0.545 | | | |
| ambulatory_visits_to_somatic | 365 | boolean | 0 | FALSE | 0.455 | 2 | 0.545 | | X | |
| ambulatory_visits_to_somatic | 365 | summed | 0 | FALSE | 2.053 | 125 | 0.545 | | | |
| ambulatory_visits_to_somatic | 730 | count | 0 | FALSE | 3.696 | 177 | 0.428 | X | | |
| ambulatory_visits_to_somatic | 730 | summed | 0 | FALSE | 3.696 | 177 | 0.428 | X | | |
| ambulatory_visits_to_somatic | 730 | boolean | 0 | FALSE | 0.572 | 2 | 0.428 | X | X | |
| ambulatory_visits_to_somatic | 10 | count | 0 | FALSE | 0.067 | 14 | 0.948 | | | |
| ambulatory_visits_to_somatic | 10 | boolean | 0 | FALSE | 0.052 | 2 | 0.948 | | | |
| ambulatory_visits_to_somatic | 10 | summed | 0 | FALSE | 0.067 | 14 | 0.948 | | | |
| ambulatory_visits_to_somatic | 30 | boolean | 0 | FALSE | 0.122 | 2 | 0.878 | X | | |
| ambulatory_visits_to_somatic | 30 | count | 0 | FALSE | 0.196 | 32 | 0.878 | X | | |
| ambulatory_visits_to_somatic | 30 | summed | 0 | FALSE | 0.196 | 32 | 0.878 | X | | |
| antidepressives | 1 | boolean | 0 | FALSE | 0.368 | 2 | 0.632 | X | X | |

| | | | | | | | | | |
|---|---|---|---|---|---|---|---|---|---|
| antidepressives | 1 | count | 0 | FALSE | 0.427 | 6 | 0.632 | X | X |
| antidepressives | 3 | count | 0 | FALSE | 1.215 | 13 | 0.619 | | X |
| antidepressives | 3 | boolean | 0 | FALSE | 0.381 | 2 | 0.619 | | X |
| antidepressives | 7 | count | 0 | FALSE | 2.588 | 29 | 0.61 | X | X |
| antidepressives | 7 | boolean | 0 | FALSE | 0.39 | 2 | 0.61 | X | X |
| antidepressives | 10 | boolean | 0 | FALSE | 0.395 | 2 | 0.605 | | X |
| antidepressives | 10 | count | 0 | FALSE | 3.477 | 41 | 0.605 | | X |
| antidepressives | 30 | count | 0 | FALSE | 7.53 | 110 | 0.584 | X | X |
| antidepressives | 30 | boolean | 0 | FALSE | 0.416 | 2 | 0.584 | X | X |
| antidepressives | 180 | boolean | 0 | FALSE | 0.459 | 2 | 0.541 | | X |
| antidepressives | 180 | count | 0 | FALSE | 18.144 | 359 | 0.541 | | X |
| antidepressives | 365 | count | 0 | FALSE | 25.776 | 643 | 0.513 | | X |
| antidepressives | 365 | boolean | 0 | FALSE | 0.487 | 2 | 0.513 | | X |
| antidepressives | 730 | boolean | 0 | FALSE | 0.523 | 2 | 0.477 | X | X |
| antidepressives | 730 | count | 0 | FALSE | 36.839 | 975 | 0.477 | X | X |
| antipsychotics | 1 | boolean | 0 | FALSE | 0.717 | 2 | 0.283 | X | |
| antipsychotics | 1 | count | 0 | FALSE | 1.457 | 13 | 0.283 | X | X |
| antipsychotics | 3 | count | 0 | FALSE | 4.163 | 33 | 0.242 | | |
| antipsychotics | 3 | boolean | 0 | FALSE | 0.758 | 2 | 0.242 | | |
| antipsychotics | 7 | boolean | 0 | FALSE | 0.784 | 2 | 0.216 | X | |
| antipsychotics | 7 | count | 0 | FALSE | 8.936 | 65 | 0.216 | X | X |
| antipsychotics | 10 | count | 0 | FALSE | 12.084 | 89 | 0.205 | | X |
| antipsychotics | 10 | boolean | 0 | FALSE | 0.795 | 2 | 0.205 | | |
| antipsychotics | 30 | boolean | 0 | FALSE | 0.825 | 2 | 0.175 | X | |
| antipsychotics | 30 | count | 0 | FALSE | 27.265 | 247 | 0.175 | X | X |
| antipsychotics | 180 | boolean | 0 | FALSE | 0.853 | 2 | 0.147 | | |
| antipsychotics | 180 | count | 0 | FALSE | 73.766 | 1014 | 0.147 | | X |
| antipsychotics | 365 | boolean | 0 | FALSE | 0.864 | 2 | 0.136 | | |
| antipsychotics | 365 | count | 0 | FALSE | 109.69 | 1771 | 0.136 | | X |
| antipsychotics | 730 | boolean | 0 | FALSE | 0.876 | 2 | 0.124 | X | |
| antipsychotics | 730 | count | 0 | FALSE | 168.27 | 2714 | 0.124 | X | X |
| antipsychotics_engangs | 1 | boolean | 0 | FALSE | 0.015 | 2 | 0.985 | X | X |
| antipsychotics_engangs | 1 | count | 0 | FALSE | 0.016 | 6 | 0.985 | X | X |
| antipsychotics_engangs | 3 | count | 0 | FALSE | 0.046 | 9 | 0.961 | | X |
| antipsychotics_engangs | 3 | boolean | 0 | FALSE | 0.039 | 2 | 0.961 | | X |
| antipsychotics_engangs | 7 | boolean | 0 | FALSE | 0.074 | 2 | 0.926 | X | X |
| antipsychotics_engangs | 7 | count | 0 | FALSE | 0.097 | 11 | 0.926 | X | X |
| antipsychotics_engangs | 10 | count | 0 | FALSE | 0.13 | 12 | 0.905 | | X |
| antipsychotics_engangs | 10 | boolean | 0 | FALSE | 0.095 | 2 | 0.905 | | X |
| antipsychotics_engangs | 30 | count | 0 | FALSE | 0.282 | 18 | 0.82 | X | X |
| antipsychotics_engangs | 30 | boolean | 0 | FALSE | 0.18 | 2 | 0.82 | X | X |
| antipsychotics_engangs | 180 | boolean | 0 | FALSE | 0.324 | 2 | 0.676 | | X |
| antipsychotics_engangs | 180 | count | 0 | FALSE | 0.649 | 20 | 0.676 | | X |
| antipsychotics_engangs | 365 | count | 0 | FALSE | 0.908 | 35 | 0.611 | | X |
| antipsychotics_engangs | 365 | boolean | 0 | FALSE | 0.389 | 2 | 0.611 | | X |
| antipsychotics_engangs | 730 | count | 0 | FALSE | 1.441 | 51 | 0.534 | X | X |
| antipsychotics_engangs | 730 | boolean | 0 | FALSE | 0.466 | 2 | 0.534 | X | X |
| antipsychotics_fast | 1 | count | 0 | FALSE | 1.018 | 10 | 0.363 | | |
| antipsychotics_fast | 1 | boolean | 0 | FALSE | 0.637 | 2 | 0.363 | | |
| antipsychotics_fast | 3 | count | 0 | FALSE | 2.914 | 23 | 0.339 | | X |
| antipsychotics_fast | 3 | boolean | 0 | FALSE | 0.661 | 2 | 0.339 | | |
| antipsychotics_fast | 7 | boolean | 0 | FALSE | 0.68 | 2 | 0.32 | | |
| antipsychotics_fast | 7 | count | 0 | FALSE | 6.266 | 49 | 0.32 | X | X |
| antipsychotics_fast | 10 | boolean | 0 | FALSE | 0.691 | 2 | 0.309 | | |
| antipsychotics_fast | 10 | count | 0 | FALSE | 8.481 | 65 | 0.309 | | X |
| antipsychotics_fast | 30 | count | 0 | FALSE | 19.22 | 179 | 0.277 | X | X |
| antipsychotics_fast | 30 | boolean | 0 | FALSE | 0.723 | 2 | 0.277 | X | |
| antipsychotics_fast | 180 | count | 0 | FALSE | 53.222 | 820 | 0.242 | | X |
| antipsychotics_fast | 180 | boolean | 0 | FALSE | 0.758 | 2 | 0.242 | | |
| antipsychotics_fast | 365 | boolean | 0 | FALSE | 0.774 | 2 | 0.226 | | |
| antipsychotics_fast | 365 | count | 0 | FALSE | 79.78 | 1308 | 0.226 | | X |
| antipsychotics_fast | 730 | boolean | 0 | FALSE | 0.792 | 2 | 0.208 | X | |
| antipsychotics_fast | 730 | count | 0 | FALSE | 123.65 | 1959 | 0.208 | X | X |
| antipsychotics_im | 1 | count | 0 | FALSE | 0.01 | 6 | 0.99 | X | |
| antipsychotics_im | 1 | boolean | 0 | FALSE | 0.01 | 2 | 0.99 | X | |
| antipsychotics_im | 3 | boolean | 0 | FALSE | 0.026 | 2 | 0.974 | | |
| antipsychotics_im | 3 | count | 0 | FALSE | 0.03 | 12 | 0.974 | | X |
| antipsychotics_im | 7 | count | 0 | FALSE | 0.064 | 21 | 0.947 | X | |
| antipsychotics_im | 7 | boolean | 0 | FALSE | 0.053 | 2 | 0.947 | X | |
| antipsychotics_im | 10 | boolean | 0 | FALSE | 0.069 | 2 | 0.931 | | |
| antipsychotics_im | 10 | count | 0 | FALSE | 0.088 | 25 | 0.931 | | |
| antipsychotics_im | 30 | boolean | 0 | FALSE | 0.116 | 2 | 0.884 | X | |
| antipsychotics_im | 30 | count | 0 | FALSE | 0.215 | 52 | 0.884 | X | |
| antipsychotics_im | 180 | boolean | 0 | FALSE | 0.151 | 2 | 0.849 | | |
| antipsychotics_im | 180 | count | 0 | FALSE | 0.751 | 72 | 0.849 | | |
| antipsychotics_im | 365 | boolean | 0 | FALSE | 0.171 | 2 | 0.829 | | |

| | | | | | | | | | |
|---|---|---|---|---|---|---|---|---|---|
| antipsychotics_im | 365 | count | 0 | FALSE | 1.277 | 107 | 0.829 | | |
| antipsychotics_im | 730 | boolean | 0 | FALSE | 0.207 | 2 | 0.793 | X | X |
| antipsychotics_im | 730 | count | 0 | FALSE | 2.319 | 126 | 0.793 | | |
| antipsychotics_pn | 1 | boolean | 0 | FALSE | 0.259 | 2 | 0.741 | X | X |
| antipsychotics_pn | 1 | count | 0 | FALSE | 0.423 | 11 | 0.741 | X | X |
| antipsychotics_pn | 3 | boolean | 0 | FALSE | 0.36 | 2 | 0.64 | | X |
| antipsychotics_pn | 3 | count | 0 | FALSE | 1.207 | 26 | 0.64 | | X |
| antipsychotics_pn | 7 | boolean | 0 | FALSE | 0.433 | 2 | 0.567 | X | X |
| antipsychotics_pn | 7 | count | 0 | FALSE | 2.582 | 53 | 0.567 | X | |
| antipsychotics_pn | 10 | boolean | 0 | FALSE | 0.463 | 2 | 0.537 | | X |
| antipsychotics_pn | 10 | count | 0 | FALSE | 3.486 | 70 | 0.537 | | |
| antipsychotics_pn | 30 | boolean | 0 | FALSE | 0.547 | 2 | 0.453 | X | |
| antipsychotics_pn | 30 | count | 0 | FALSE | 7.791 | 164 | 0.453 | X | X |
| antipsychotics_pn | 180 | boolean | 0 | FALSE | 0.641 | 2 | 0.359 | | |
| antipsychotics_pn | 180 | count | 0 | FALSE | 19.976 | 527 | 0.359 | | X |
| antipsychotics_pn | 365 | boolean | 0 | FALSE | 0.677 | 2 | 0.323 | | |
| antipsychotics_pn | 365 | count | 0 | FALSE | 29.126 | 970 | 0.323 | | X |
| antipsychotics_pn | 730 | boolean | 0 | FALSE | 0.719 | 2 | 0.281 | X | |
| antipsychotics_pn | 730 | count | 0 | FALSE | 43.373 | 1567 | 0.281 | X | X |
| antipsychotics_po | 1 | boolean | 0 | FALSE | 0.001 | 2 | 0.999 | X | |
| antipsychotics_po | 1 | count | 0 | FALSE | 0.001 | 4 | 0.999 | | |
| antipsychotics_po | 3 | count | 0 | FALSE | 0.003 | 10 | 0.999 | | |
| antipsychotics_po | 3 | boolean | 0 | FALSE | 0.001 | 2 | 0.999 | | |
| antipsychotics_po | 7 | boolean | 0 | FALSE | 0.001 | 2 | 0.999 | X | |
| antipsychotics_po | 7 | count | 0 | FALSE | 0.008 | 22 | 0.999 | | |
| antipsychotics_po | 10 | count | 0 | FALSE | 0.01 | 31 | 0.999 | | |
| antipsychotics_po | 10 | boolean | 0 | FALSE | 0.001 | 2 | 0.999 | | |
| antipsychotics_po | 30 | boolean | 0 | FALSE | 0.001 | 2 | 0.999 | X | |
| antipsychotics_po | 30 | count | 0 | FALSE | 0.024 | 89 | 0.999 | | |
| antipsychotics_po | 180 | boolean | 0 | FALSE | 0.001 | 2 | 0.999 | | |
| antipsychotics_po | 180 | count | 0 | FALSE | 0.047 | 174 | 0.999 | | |
| antipsychotics_po | 365 | count | 0 | FALSE | 0.055 | 178 | 0.998 | | |
| antipsychotics_po | 365 | boolean | 0 | FALSE | 0.002 | 2 | 0.998 | | |
| antipsychotics_po | 730 | count | 0 | FALSE | 0.099 | 217 | 0.997 | X | |
| antipsychotics_po | 730 | boolean | 0 | FALSE | 0.003 | 2 | 0.997 | | |
| anxiolytics | 1 | boolean | 0 | FALSE | 0.237 | 2 | 0.763 | X | X |
| anxiolytics | 1 | count | 0 | FALSE | 0.472 | 17 | 0.763 | X | X |
| anxiolytics | 3 | boolean | 0 | FALSE | 0.299 | 2 | 0.701 | | X |
| anxiolytics | 3 | count | 0 | FALSE | 1.361 | 34 | 0.701 | | |
| anxiolytics | 7 | count | 0 | FALSE | 2.946 | 60 | 0.649 | | |
| anxiolytics | 7 | boolean | 0 | FALSE | 0.351 | 2 | 0.649 | X | X |
| anxiolytics | 10 | boolean | 0 | FALSE | 0.375 | 2 | 0.625 | | X |
| anxiolytics | 10 | count | 0 | FALSE | 4.002 | 80 | 0.625 | | |
| anxiolytics | 30 | count | 0 | FALSE | 9.058 | 217 | 0.547 | X | X |
| anxiolytics | 30 | boolean | 0 | FALSE | 0.453 | 2 | 0.547 | X | X |
| anxiolytics | 180 | count | 0 | FALSE | 21.33 | 720 | 0.447 | | X |
| anxiolytics | 180 | boolean | 0 | FALSE | 0.553 | 2 | 0.447 | | |
| anxiolytics | 365 | boolean | 0 | FALSE | 0.597 | 2 | 0.403 | | |
| anxiolytics | 365 | count | 0 | FALSE | 29.34 | 965 | 0.403 | | X |
| anxiolytics | 730 | boolean | 0 | FALSE | 0.647 | 2 | 0.353 | X | |
| anxiolytics | 730 | count | 0 | FALSE | 42.119 | 1275 | 0.353 | X | X |
| anxiolytics_engangs | 1 | count | 0 | FALSE | 0.011 | 7 | 0.99 | X | X |
| anxiolytics_engangs | 1 | boolean | 0 | FALSE | 0.01 | 2 | 0.99 | X | X |
| anxiolytics_engangs | 3 | boolean | 0 | FALSE | 0.027 | 2 | 0.973 | | X |
| anxiolytics_engangs | 3 | count | 0 | FALSE | 0.033 | 9 | 0.973 | | X |
| anxiolytics_engangs | 7 | boolean | 0 | FALSE | 0.052 | 2 | 0.948 | X | X |
| anxiolytics_engangs | 7 | count | 0 | FALSE | 0.069 | 9 | 0.948 | X | X |
| anxiolytics_engangs | 10 | boolean | 0 | FALSE | 0.067 | 2 | 0.933 | | X |
| anxiolytics_engangs | 10 | count | 0 | FALSE | 0.093 | 12 | 0.933 | | X |
| anxiolytics_engangs | 30 | count | 0 | FALSE | 0.204 | 17 | 0.87 | X | X |
| anxiolytics_engangs | 30 | boolean | 0 | FALSE | 0.13 | 2 | 0.87 | X | X |
| anxiolytics_engangs | 180 | count | 0 | FALSE | 0.472 | 20 | 0.765 | | |
| anxiolytics_engangs | 180 | boolean | 0 | FALSE | 0.235 | 2 | 0.765 | | |
| anxiolytics_engangs | 365 | boolean | 0 | FALSE | 0.288 | 2 | 0.712 | | |
| anxiolytics_engangs | 365 | count | 0 | FALSE | 0.651 | 24 | 0.712 | | |
| anxiolytics_engangs | 730 | boolean | 0 | FALSE | 0.353 | 2 | 0.647 | X | |
| anxiolytics_engangs | 730 | count | 0 | FALSE | 0.965 | 39 | 0.647 | X | |
| anxiolytics_fast | 1 | count | 0 | FALSE | 0.21 | 9 | 0.904 | X | |
| anxiolytics_fast | 1 | boolean | 0 | FALSE | 0.096 | 2 | 0.904 | | |
| anxiolytics_fast | 3 | boolean | 0 | FALSE | 0.103 | 2 | 0.897 | | |
| anxiolytics_fast | 3 | count | 0 | FALSE | 0.607 | 22 | 0.897 | | |
| anxiolytics_fast | 7 | count | 0 | FALSE | 1.32 | 46 | 0.889 | X | |
| anxiolytics_fast | 7 | boolean | 0 | FALSE | 0.111 | 2 | 0.889 | | |
| anxiolytics_fast | 10 | count | 0 | FALSE | 1.796 | 66 | 0.884 | | |
| anxiolytics_fast | 10 | boolean | 0 | FALSE | 0.116 | 2 | 0.884 | | |
| anxiolytics_fast | 30 | count | 0 | FALSE | 4.061 | 182 | 0.862 | X | X |

| | | | | | | | | | | |
|---|---|---|---|---|---|---|---|---|---|---|
| anxiolytics_fast | 30 | boolean | 0 | FALSE | 0.138 | 2 | 0.862 | X | | |
| anxiolytics_fast | 180 | count | 0 | FALSE | 9.502 | 671 | 0.815 | | | |
| anxiolytics_fast | 180 | boolean | 0 | FALSE | 0.185 | 2 | 0.815 | | | |
| anxiolytics_fast | 365 | boolean | 0 | FALSE | 0.214 | 2 | 0.786 | | | |
| anxiolytics_fast | 365 | count | 0 | FALSE | 12.798 | 867 | 0.786 | | | |
| anxiolytics_fast | 730 | boolean | 0 | FALSE | 0.253 | 2 | 0.747 | X | | |
| anxiolytics_fast | 730 | count | 0 | FALSE | 18.056 | 1020 | 0.747 | X | | |
| anxiolytics_pn | 1 | boolean | 0 | FALSE | 0.157 | 2 | 0.843 | X | X | |
| anxiolytics_pn | 1 | count | 0 | FALSE | 0.251 | 15 | 0.843 | X | X | |
| anxiolytics_pn | 3 | boolean | 0 | FALSE | 0.222 | 2 | 0.778 | | X | |
| anxiolytics_pn | 3 | count | 0 | FALSE | 0.72 | 30 | 0.778 | | X | |
| anxiolytics_pn | 7 | boolean | 0 | FALSE | 0.275 | 2 | 0.725 | X | X | |
| anxiolytics_pn | 7 | count | 0 | FALSE | 1.556 | 45 | 0.725 | X | | |
| anxiolytics_pn | 10 | boolean | 0 | FALSE | 0.299 | 2 | 0.701 | | X | |
| anxiolytics_pn | 10 | count | 0 | FALSE | 2.112 | 60 | 0.701 | | | |
| anxiolytics_pn | 30 | boolean | 0 | FALSE | 0.376 | 2 | 0.624 | X | | |
| anxiolytics_pn | 30 | count | 0 | FALSE | 4.789 | 130 | 0.624 | X | | |
| anxiolytics_pn | 180 | boolean | 0 | FALSE | 0.474 | 2 | 0.526 | | | |
| anxiolytics_pn | 180 | count | 0 | FALSE | 11.352 | 391 | 0.526 | | X | |
| anxiolytics_pn | 365 | boolean | 0 | FALSE | 0.518 | 2 | 0.482 | | | |
| anxiolytics_pn | 365 | count | 0 | FALSE | 15.887 | 481 | 0.482 | | X | |
| anxiolytics_pn | 730 | boolean | 0 | FALSE | 0.569 | 2 | 0.431 | X | | |
| anxiolytics_pn | 730 | count | 0 | FALSE | 23.096 | 717 | 0.431 | X | X | |
| baelte | 730 | count | 0 | FALSE | 0.069 | 41 | 0.97 | X | X | |
| baelte | 730 | summed | 0 | FALSE | 1.555 | 551 | 0.97 | X | X | |
| baelte | 730 | boolean | 0 | FALSE | 0.03 | 2 | 0.97 | X | X | |
| beroligende_medicin | 730 | count | 0 | FALSE | 0.088 | 26 | 0.964 | X | X | |
| beroligende_medicin | 730 | boolean | 0 | FALSE | 0.036 | 2 | 0.964 | X | X | |
| bmi | 10 | latest | NaN | FALSE | 25.984 | 484 | 0.754 | | | |
| bmi | 365 | latest | NaN | FALSE | 26.917 | 505 | 0.213 | | | |
| bmi | 730 | latest | NaN | FALSE | 26.943 | 507 | 0.141 | | | |
| bmi | 180 | latest | NaN | FALSE | 26.81 | 500 | 0.299 | | X | |
| bmi | 30 | latest | NaN | FALSE | 26.351 | 485 | 0.558 | X | X | |
| broeset_violence_checklist | 1 | maximum | NaN | FALSE | 0.433 | 8 | 0.652 | X | X | X |
| broeset_violence_checklist | 1 | mean | NaN | FALSE | 0.298 | 84 | 0.652 | X | X | |
| broeset_violence_checklist | 1 | day | NaN | FALSE | -0.249 | 22421 | 0.792 | | X | |
| broeset_violence_checklist | 1 | variance | NaN | FALSE | 0.352 | 233 | 0.792 | X | X | |
| broeset_violence_checklist | 1 | minimum | NaN | FALSE | 0.187 | 8 | 0.652 | X | X | |
| broeset_violence_checklist | 3 | mean | NaN | FALSE | 0.309 | 284 | 0.512 | | X | |
| broeset_violence_checklist | 3 | maximum | NaN | FALSE | 0.629 | 8 | 0.512 | | X | |
| broeset_violence_checklist | 3 | day | NaN | FALSE | -0.2 | 77134 | 0.641 | | X | |
| broeset_violence_checklist | 3 | variance | NaN | FALSE | 0.388 | 1582 | 0.641 | | X | |
| broeset_violence_checklist | 3 | minimum | NaN | FALSE | 0.131 | 8 | 0.512 | | X | |
| broeset_violence_checklist | 7 | maximum | NaN | FALSE | 0.784 | 8 | 0.376 | X | X | |
| broeset_violence_checklist | 7 | mean | NaN | FALSE | 0.309 | 763 | 0.376 | X | X | |
| broeset_violence_checklist | 7 | minimum | NaN | FALSE | 0.103 | 8 | 0.376 | X | X | |
| broeset_violence_checklist | 7 | variance | NaN | FALSE | 0.402 | 7417 | 0.513 | X | X | |
| broeset_violence_checklist | 7 | day | NaN | FALSE | -0.177 | 115044 | 0.513 | | X | |
| broeset_violence_checklist | 10 | mean | NaN | FALSE | 0.308 | 1223 | 0.314 | | X | |
| broeset_violence_checklist | 10 | maximum | NaN | FALSE | 0.85 | 8 | 0.314 | | X | |
| broeset_violence_checklist | 10 | day | NaN | FALSE | -0.147 | 128865 | 0.454 | | X | |
| broeset_violence_checklist | 10 | minimum | NaN | FALSE | 0.093 | 8 | 0.314 | | X | |
| broeset_violence_checklist | 10 | variance | NaN | FALSE | 0.406 | 13323 | 0.454 | | X | |
| broeset_violence_checklist | 30 | maximum | NaN | FALSE | 1.05 | 8 | 0.143 | X | X | |
| broeset_violence_checklist | 30 | mean | NaN | FALSE | 0.304 | 4681 | 0.143 | X | X | |
| broeset_violence_checklist | 30 | day | NaN | FALSE | -0.12 | 158315 | 0.275 | | X | |
| broeset_violence_checklist | 30 | variance | NaN | FALSE | 0.416 | 43700 | 0.275 | X | X | |
| broeset_violence_checklist | 30 | minimum | NaN | FALSE | 0.063 | 8 | 0.143 | X | X | |
| broeset_violence_checklist | 180 | maximum | NaN | FALSE | 1.397 | 8 | 0.049 | | X | |
| broeset_violence_checklist | 180 | mean | NaN | FALSE | 0.298 | 21659 | 0.049 | | X | |
| broeset_violence_checklist | 180 | variance | NaN | FALSE | 0.422 | 92284 | 0.134 | | X | |
| broeset_violence_checklist | 180 | day | NaN | FALSE | -0.139 | 179021 | 0.134 | | X | |
| broeset_violence_checklist | 180 | minimum | NaN | FALSE | 0.036 | 8 | 0.049 | | X | |
| broeset_violence_checklist | 365 | mean | NaN | FALSE | 0.295 | 32533 | 0.038 | | X | |
| broeset_violence_checklist | 365 | maximum | NaN | FALSE | 1.551 | 8 | 0.038 | | X | |
| broeset_violence_checklist | 365 | variance | NaN | FALSE | 0.418 | 110721 | 0.109 | | X | |
| broeset_violence_checklist | 365 | day | NaN | FALSE | -0.133 | 189007 | 0.109 | | X | |
| broeset_violence_checklist | 365 | minimum | NaN | FALSE | 0.031 | 8 | 0.038 | | X | |
| broeset_violence_checklist | 730 | maximum | NaN | FALSE | 1.767 | 8 | 0.032 | X | X | |
| broeset_violence_checklist | 730 | mean | NaN | FALSE | 0.294 | 46780 | 0.032 | X | X | |
| broeset_violence_checklist | 730 | variance | NaN | FALSE | 0.433 | 129814 | 0.091 | X | X | |
| broeset_violence_checklist | 730 | minimum | NaN | FALSE | 0.025 | 8 | 0.032 | X | X | |
| broeset_violence_checklist | 730 | day | NaN | FALSE | -0.11 | 196086 | 0.091 | | X | |
| cancelled_standard_lab_results | 10 | maximum | NaN | FALSE | 1 | 2 | 0.948 | | | |
| cancelled_standard_lab_results | 10 | mean | NaN | FALSE | 1 | 2 | 0.948 | | | |
| cancelled_standard_lab_results | 10 | latest | NaN | FALSE | 1 | 2 | 0.948 | | | |

| | | | | | | | | | |
|---|---|---|---|---|---|---|---|---|---|
| cancelled_standard_lab_results | 10 | minimum | NaN | FALSE | 1 | 2 | 0.948 | | |
| cancelled_standard_lab_results | 30 | maximum | NaN | FALSE | 1 | 2 | 0.892 | | |
| cancelled_standard_lab_results | 30 | minimum | NaN | FALSE | 1 | 2 | 0.892 | | |
| cancelled_standard_lab_results | 30 | mean | NaN | FALSE | 1 | 2 | 0.892 | | |
| cancelled_standard_lab_results | 30 | latest | NaN | FALSE | 1 | 2 | 0.892 | | |
| cancelled_standard_lab_results | 180 | mean | NaN | FALSE | 1 | 2 | 0.77 | | |
| cancelled_standard_lab_results | 180 | maximum | NaN | FALSE | 1 | 2 | 0.77 | | |
| cancelled_standard_lab_results | 180 | minimum | NaN | FALSE | 1 | 2 | 0.77 | | |
| cancelled_standard_lab_results | 180 | latest | NaN | FALSE | 1 | 2 | 0.77 | | |
| cancelled_standard_lab_results | 365 | mean | NaN | FALSE | 1 | 2 | 0.706 | | |
| cancelled_standard_lab_results | 365 | maximum | NaN | FALSE | 1 | 2 | 0.706 | | |
| cancelled_standard_lab_results | 365 | minimum | NaN | FALSE | 1 | 2 | 0.706 | | |
| cancelled_standard_lab_results | 365 | latest | NaN | FALSE | 1 | 2 | 0.706 | | |
| cancelled_standard_lab_results | 730 | maximum | NaN | FALSE | 1 | 2 | 0.631 | | |
| cancelled_standard_lab_results | 730 | minimum | NaN | FALSE | 1 | 2 | 0.631 | | |
| cancelled_standard_lab_results | 730 | latest | NaN | FALSE | 1 | 2 | 0.631 | | |
| cancelled_standard_lab_results | 730 | mean | NaN | FALSE | 1 | 2 | 0.631 | | |
| clozapine | 1 | count | 0 | FALSE | 0.124 | 7 | 0.918 | X | X |
| clozapine | 1 | boolean | 0 | FALSE | 0.082 | 2 | 0.918 | X | X |
| clozapine | 3 | count | 0 | FALSE | 0.355 | 18 | 0.916 | | X |
| clozapine | 3 | boolean | 0 | FALSE | 0.084 | 2 | 0.916 | | X |
| clozapine | 7 | boolean | 0 | FALSE | 0.084 | 2 | 0.916 | X | X |
| clozapine | 7 | count | 0 | FALSE | 0.77 | 33 | 0.916 | X | X |
| clozapine | 10 | count | 0 | FALSE | 1.051 | 43 | 0.915 | | X |
| clozapine | 10 | boolean | 0 | FALSE | 0.085 | 2 | 0.915 | | X |
| clozapine | 30 | boolean | 0 | FALSE | 0.087 | 2 | 0.913 | X | X |
| clozapine | 30 | count | 0 | FALSE | 2.552 | 121 | 0.913 | X | X |
| clozapine | 180 | boolean | 0 | FALSE | 0.093 | 2 | 0.907 | | X |
| clozapine | 180 | count | 0 | FALSE | 8.735 | 586 | 0.907 | | X |
| clozapine | 365 | count | 0 | FALSE | 13.919 | 892 | 0.903 | | X |
| clozapine | 365 | boolean | 0 | FALSE | 0.097 | 2 | 0.903 | | X |
| clozapine | 730 | count | 0 | FALSE | 22.528 | 1578 | 0.895 | X | |
| clozapine | 730 | boolean | 0 | FALSE | 0.105 | 2 | 0.895 | X | |
| clozapine_engangs | 1 | count | 0 | FALSE | 0 | 3 | 1 | | |
| clozapine_engangs | 1 | boolean | 0 | FALSE | 0 | 2 | 1 | | |
| clozapine_engangs | 3 | count | 0 | FALSE | 0.001 | 4 | 0.999 | | |
| clozapine_engangs | 3 | boolean | 0 | FALSE | 0.001 | 2 | 0.999 | | |
| clozapine_engangs | 7 | boolean | 0 | FALSE | 0.002 | 2 | 0.998 | X | |
| clozapine_engangs | 7 | count | 0 | FALSE | 0.002 | 5 | 0.998 | X | |
| clozapine_engangs | 10 | count | 0 | FALSE | 0.002 | 5 | 0.998 | | |
| clozapine_engangs | 10 | boolean | 0 | FALSE | 0.002 | 2 | 0.998 | | |
| clozapine_engangs | 30 | boolean | 0 | FALSE | 0.005 | 2 | 0.995 | X | |
| clozapine_engangs | 30 | count | 0 | FALSE | 0.006 | 5 | 0.995 | X | |
| clozapine_engangs | 180 | boolean | 0 | FALSE | 0.013 | 2 | 0.987 | | |
| clozapine_engangs | 180 | count | 0 | FALSE | 0.015 | 6 | 0.987 | | |
| clozapine_engangs | 365 | count | 0 | FALSE | 0.021 | 6 | 0.983 | | |
| clozapine_engangs | 365 | boolean | 0 | FALSE | 0.017 | 2 | 0.983 | | |
| clozapine_engangs | 730 | count | 0 | FALSE | 0.032 | 7 | 0.976 | X | |
| clozapine_engangs | 730 | boolean | 0 | FALSE | 0.024 | 2 | 0.976 | X | |
| clozapine_fast | 1 | boolean | 0 | FALSE | 0.082 | 2 | 0.918 | X | X |
| clozapine_fast | 1 | count | 0 | FALSE | 0.123 | 7 | 0.918 | X | X |
| clozapine_fast | 3 | boolean | 0 | FALSE | 0.083 | 2 | 0.917 | | X |
| clozapine_fast | 3 | count | 0 | FALSE | 0.353 | 18 | 0.917 | | X |
| clozapine_fast | 7 | count | 0 | FALSE | 0.765 | 33 | 0.916 | X | X |
| clozapine_fast | 7 | boolean | 0 | FALSE | 0.084 | 2 | 0.916 | X | X |
| clozapine_fast | 10 | count | 0 | FALSE | 1.044 | 43 | 0.915 | | X |
| clozapine_fast | 10 | boolean | 0 | FALSE | 0.085 | 2 | 0.915 | | X |
| clozapine_fast | 30 | count | 0 | FALSE | 2.537 | 121 | 0.914 | X | X |
| clozapine_fast | 30 | boolean | 0 | FALSE | 0.086 | 2 | 0.914 | X | X |
| clozapine_fast | 180 | boolean | 0 | FALSE | 0.093 | 2 | 0.907 | | X |
| clozapine_fast | 180 | count | 0 | FALSE | 8.693 | 579 | 0.907 | | X |
| clozapine_fast | 365 | count | 0 | FALSE | 13.863 | 892 | 0.903 | | X |
| clozapine_fast | 365 | boolean | 0 | FALSE | 0.097 | 2 | 0.903 | | X |
| clozapine_fast | 730 | count | 0 | FALSE | 22.443 | 1580 | 0.896 | X | |
| clozapine_fast | 730 | boolean | 0 | FALSE | 0.104 | 2 | 0.896 | X | |
| clozapine_pn | 1 | boolean | 0 | FALSE | 0 | 2 | 1 | | |
| clozapine_pn | 1 | count | 0 | FALSE | 0 | 3 | 1 | | |
| clozapine_pn | 3 | boolean | 0 | FALSE | 0 | 2 | 1 | | |
| clozapine_pn | 3 | count | 0 | FALSE | 0 | 7 | 1 | | |
| clozapine_pn | 7 | boolean | 0 | FALSE | 0 | 2 | 1 | | |
| clozapine_pn | 7 | count | 0 | FALSE | 0.001 | 11 | 1 | | |
| clozapine_pn | 10 | boolean | 0 | FALSE | 0 | 2 | 1 | | |
| clozapine_pn | 10 | count | 0 | FALSE | 0.001 | 13 | 1 | | |
| clozapine_pn | 30 | boolean | 0 | FALSE | 0 | 2 | 1 | | |
| clozapine_pn | 30 | count | 0 | FALSE | 0.002 | 17 | 1 | | |
| clozapine_pn | 180 | boolean | 0 | FALSE | 0.001 | 2 | 0.999 | | |

| | | | | | | | | | |
|---|---|---|---|---|---|---|---|---|---|
| clozapine_pn | 180 | count | 0 | FALSE | 0.003 | 18 | 0.999 | | |
| clozapine_pn | 365 | boolean | 0 | FALSE | 0.001 | 2 | 0.999 | | |
| clozapine_pn | 365 | count | 0 | FALSE | 0.004 | 18 | 0.999 | | |
| clozapine_pn | 730 | boolean | 0 | FALSE | 0.002 | 2 | 0.998 | X | |
| clozapine_pn | 730 | count | 0 | FALSE | 0.016 | 45 | 0.998 | X | |
| clozapine_po | 1 | count | 0 | FALSE | 0.001 | 4 | 1 | | |
| clozapine_po | 1 | boolean | 0 | FALSE | 0 | 2 | 1 | | |
| clozapine_po | 3 | count | 0 | FALSE | 0.003 | 10 | 1 | | |
| clozapine_po | 3 | boolean | 0 | FALSE | 0 | 2 | 1 | | |
| clozapine_po | 7 | count | 0 | FALSE | 0.006 | 22 | 1 | | |
| clozapine_po | 7 | boolean | 0 | FALSE | 0 | 2 | 1 | | |
| clozapine_po | 10 | boolean | 0 | FALSE | 0 | 2 | 1 | | |
| clozapine_po | 10 | count | 0 | FALSE | 0.008 | 31 | 1 | | |
| clozapine_po | 30 | boolean | 0 | FALSE | 0 | 2 | 1 | | |
| clozapine_po | 30 | count | 0 | FALSE | 0.02 | 88 | 1 | | |
| clozapine_po | 180 | boolean | 0 | FALSE | 0 | 2 | 1 | | |
| clozapine_po | 180 | count | 0 | FALSE | 0.036 | 170 | 1 | | |
| clozapine_po | 365 | count | 0 | FALSE | 0.039 | 176 | 0.999 | | |
| clozapine_po | 365 | boolean | 0 | FALSE | 0.001 | 2 | 0.999 | | |
| clozapine_po | 730 | boolean | 0 | FALSE | 0.001 | 2 | 0.999 | X | |
| clozapine_po | 730 | count | 0 | FALSE | 0.06 | 209 | 0.999 | | |
| depressive_disorders | 10 | boolean | 0 | FALSE | 0.062 | 2 | 0.938 | | X |
| depressive_disorders | 30 | boolean | 0 | FALSE | 0.129 | 2 | 0.871 | X | X |
| depressive_disorders | 180 | boolean | 0 | FALSE | 0.214 | 2 | 0.786 | | X |
| depressive_disorders | 365 | boolean | 0 | FALSE | 0.246 | 2 | 0.754 | | X |
| depressive_disorders | 730 | boolean | 0 | FALSE | 0.284 | 2 | 0.716 | X | X |
| ect | 10 | boolean | 0 | FALSE | 0.001 | 2 | 0.999 | | |
| ect | 10 | count | 0 | FALSE | 0.001 | 2 | 0.999 | | |
| ect | 10 | summed | 0 | FALSE | 0.173 | 24 | 0.999 | | |
| ect | 30 | boolean | 0 | FALSE | 0.001 | 2 | 0.999 | X | |
| ect | 30 | summed | 0 | FALSE | 0.313 | 24 | 0.999 | X | |
| ect | 30 | count | 0 | FALSE | 0.001 | 3 | 0.999 | X | |
| ect | 180 | count | 0 | FALSE | 0.002 | 3 | 0.998 | | |
| ect | 180 | boolean | 0 | FALSE | 0.002 | 2 | 0.998 | | |
| ect | 180 | summed | 0 | FALSE | 0.625 | 24 | 0.998 | | |
| ect | 365 | summed | 0 | FALSE | 0.674 | 24 | 0.998 | | |
| ect | 365 | count | 0 | FALSE | 0.002 | 3 | 0.998 | | |
| ect | 365 | boolean | 0 | FALSE | 0.002 | 2 | 0.998 | | |
| ect | 730 | boolean | 0 | FALSE | 0.004 | 2 | 0.996 | X | |
| ect | 730 | summed | 0 | FALSE | 1.259 | 32 | 0.996 | | |
| ect | 730 | count | 0 | FALSE | 0.004 | 5 | 0.996 | X | |
| ect | 1 | boolean | 0 | FALSE | 0.001 | 2 | 0.999 | X | X |
| ect | 3 | boolean | 0 | FALSE | 0.001 | 2 | 0.999 | | X |
| ect | 7 | boolean | 0 | FALSE | 0 | 2 | 1 | | |
| ect | 7 | summed | 0 | FALSE | 0.128 | 24 | 1 | | |
| ect | 7 | count | 0 | FALSE | 0 | 2 | 1 | | |
| emergency_visits | 180 | count | 0 | FALSE | 1.219 | 108 | 0.5 | | |
| emergency_visits | 180 | boolean | 0 | FALSE | 0.5 | 2 | 0.5 | | |
| emergency_visits | 180 | summed | 0 | FALSE | 1.219 | 108 | 0.5 | | |
| emergency_visits | 365 | boolean | 0 | FALSE | 0.575 | 2 | 0.425 | | |
| emergency_visits | 365 | count | 0 | FALSE | 1.918 | 159 | 0.425 | | |
| emergency_visits | 365 | summed | 0 | FALSE | 1.918 | 159 | 0.425 | | |
| emergency_visits | 730 | boolean | 0 | FALSE | 0.66 | 2 | 0.34 | X | |
| emergency_visits | 730 | count | 0 | FALSE | 3.093 | 171 | 0.34 | X | |
| emergency_visits | 730 | summed | 0 | FALSE | 3.093 | 171 | 0.34 | X | |
| emergency_visits | 10 | summed | 0 | FALSE | 0.201 | 14 | 0.838 | | X |
| emergency_visits | 10 | boolean | 0 | FALSE | 0.162 | 2 | 0.838 | | X |
| emergency_visits | 10 | count | 0 | FALSE | 0.201 | 14 | 0.838 | | X |
| emergency_visits | 30 | boolean | 0 | FALSE | 0.307 | 2 | 0.693 | X | X |
| emergency_visits | 30 | count | 0 | FALSE | 0.444 | 32 | 0.693 | X | X |
| emergency_visits | 30 | summed | 0 | FALSE | 0.444 | 32 | 0.693 | X | X |
| emergency_visits_to_psychiatry | 180 | boolean | 0 | FALSE | 0.301 | 2 | 0.699 | | |
| emergency_visits_to_psychiatry | 180 | count | 0 | FALSE | 0.656 | 94 | 0.699 | | |
| emergency_visits_to_psychiatry | 180 | summed | 0 | FALSE | 0.656 | 94 | 0.699 | | |
| emergency_visits_to_psychiatry | 365 | boolean | 0 | FALSE | 0.335 | 2 | 0.665 | | |
| emergency_visits_to_psychiatry | 365 | summed | 0 | FALSE | 1.012 | 123 | 0.665 | | |
| emergency_visits_to_psychiatry | 365 | count | 0 | FALSE | 1.012 | 123 | 0.665 | | |
| emergency_visits_to_psychiatry | 730 | count | 0 | FALSE | 1.627 | 123 | 0.614 | | |
| emergency_visits_to_psychiatry | 730 | boolean | 0 | FALSE | 0.386 | 2 | 0.614 | X | |
| emergency_visits_to_psychiatry | 730 | summed | 0 | FALSE | 1.627 | 123 | 0.614 | | |
| emergency_visits_to_psychiatry | 10 | summed | 0 | FALSE | 0.124 | 12 | 0.892 | | X |
| emergency_visits_to_psychiatry | 10 | boolean | 0 | FALSE | 0.108 | 2 | 0.892 | | X |
| emergency_visits_to_psychiatry | 10 | count | 0 | FALSE | 0.124 | 12 | 0.892 | | X |
| emergency_visits_to_psychiatry | 30 | boolean | 0 | FALSE | 0.201 | 2 | 0.799 | X | |
| emergency_visits_to_psychiatry | 30 | summed | 0 | FALSE | 0.263 | 29 | 0.799 | X | |
| emergency_visits_to_psychiatry | 30 | count | 0 | FALSE | 0.263 | 29 | 0.799 | X | |

| | | | | | | | | | | |
|---|---|---|---|---|---|---|---|---|---|---|
| emergency_visits_to_somatic | 180 | boolean | 0 | FALSE | 0.272 | 2 | 0.728 | | | |
| emergency_visits_to_somatic | 180 | count | 0 | FALSE | 0.563 | 56 | 0.728 | | | |
| emergency_visits_to_somatic | 180 | summed | 0 | FALSE | 0.563 | 56 | 0.728 | | | |
| emergency_visits_to_somatic | 365 | count | 0 | FALSE | 0.906 | 88 | 0.648 | | | |
| emergency_visits_to_somatic | 365 | boolean | 0 | FALSE | 0.352 | 2 | 0.648 | | | |
| emergency_visits_to_somatic | 365 | summed | 0 | FALSE | 0.906 | 88 | 0.648 | | | |
| emergency_visits_to_somatic | 730 | count | 0 | FALSE | 1.468 | 132 | 0.548 | X | | |
| emergency_visits_to_somatic | 730 | boolean | 0 | FALSE | 0.452 | 2 | 0.548 | X | | |
| emergency_visits_to_somatic | 730 | summed | 0 | FALSE | 1.468 | 132 | 0.548 | X | | |
| emergency_visits_to_somatic | 10 | boolean | 0 | FALSE | 0.064 | 2 | 0.936 | | X | |
| emergency_visits_to_somatic | 10 | summed | 0 | FALSE | 0.078 | 10 | 0.936 | | X | |
| emergency_visits_to_somatic | 10 | count | 0 | FALSE | 0.078 | 10 | 0.936 | | X | |
| emergency_visits_to_somatic | 30 | boolean | 0 | FALSE | 0.131 | 2 | 0.869 | X | X | |
| emergency_visits_to_somatic | 30 | count | 0 | FALSE | 0.182 | 19 | 0.869 | X | X | |
| emergency_visits_to_somatic | 30 | summed | 0 | FALSE | 0.182 | 19 | 0.869 | X | X | |
| f0_disorders | 10 | boolean | 0 | FALSE | 0.007 | 2 | 0.993 | | X | |
| f0_disorders | 30 | boolean | 0 | FALSE | 0.015 | 2 | 0.985 | X | X | |
| f0_disorders | 180 | boolean | 0 | FALSE | 0.032 | 2 | 0.968 | | X | |
| f0_disorders | 365 | boolean | 0 | FALSE | 0.04 | 2 | 0.96 | | X | |
| f0_disorders | 730 | boolean | 0 | FALSE | 0.052 | 2 | 0.948 | X | X | X |
| f1_disorders | 10 | boolean | 0 | FALSE | 0.032 | 2 | 0.968 | | | |
| f1_disorders | 30 | boolean | 0 | FALSE | 0.067 | 2 | 0.933 | X | | |
| f1_disorders | 180 | boolean | 0 | FALSE | 0.161 | 2 | 0.839 | | | |
| f1_disorders | 365 | boolean | 0 | FALSE | 0.208 | 2 | 0.792 | | | |
| f1_disorders | 730 | boolean | 0 | FALSE | 0.249 | 2 | 0.751 | X | | X |
| f2_disorders | 10 | boolean | 0 | FALSE | 0.093 | 2 | 0.907 | | X | |
| f2_disorders | 30 | boolean | 0 | FALSE | 0.181 | 2 | 0.819 | X | X | |
| f2_disorders | 180 | boolean | 0 | FALSE | 0.32 | 2 | 0.68 | | | |
| f2_disorders | 365 | boolean | 0 | FALSE | 0.364 | 2 | 0.636 | | | |
| f2_disorders | 730 | boolean | 0 | FALSE | 0.394 | 2 | 0.606 | X | X | X |
| f3_disorders | 10 | boolean | 0 | FALSE | 0.101 | 2 | 0.899 | | | |
| f3_disorders | 30 | boolean | 0 | FALSE | 0.208 | 2 | 0.792 | X | X | |
| f3_disorders | 180 | boolean | 0 | FALSE | 0.331 | 2 | 0.669 | | X | |
| f3_disorders | 365 | boolean | 0 | FALSE | 0.37 | 2 | 0.63 | | X | |
| f3_disorders | 730 | boolean | 0 | FALSE | 0.41 | 2 | 0.59 | X | X | X |
| f4_disorders | 10 | boolean | 0 | FALSE | 0.045 | 2 | 0.955 | | | |
| f4_disorders | 30 | boolean | 0 | FALSE | 0.089 | 2 | 0.911 | X | | |
| f4_disorders | 180 | boolean | 0 | FALSE | 0.185 | 2 | 0.815 | | X | |
| f4_disorders | 365 | boolean | 0 | FALSE | 0.23 | 2 | 0.77 | | X | |
| f4_disorders | 730 | boolean | 0 | FALSE | 0.281 | 2 | 0.719 | X | X | X |
| f5_disorders | 10 | boolean | 0 | FALSE | 0.008 | 2 | 0.992 | | | |
| f5_disorders | 30 | boolean | 0 | FALSE | 0.018 | 2 | 0.982 | X | | |
| f5_disorders | 180 | boolean | 0 | FALSE | 0.041 | 2 | 0.959 | | | |
| f5_disorders | 365 | boolean | 0 | FALSE | 0.047 | 2 | 0.953 | | | |
| f5_disorders | 730 | boolean | 0 | FALSE | 0.052 | 2 | 0.948 | X | | X |
| f6_disorders | 10 | boolean | 0 | FALSE | 0.022 | 2 | 0.978 | | | |
| f6_disorders | 30 | boolean | 0 | FALSE | 0.041 | 2 | 0.959 | X | | |
| f6_disorders | 180 | boolean | 0 | FALSE | 0.08 | 2 | 0.92 | | | |
| f6_disorders | 365 | boolean | 0 | FALSE | 0.097 | 2 | 0.903 | | | |
| f6_disorders | 730 | boolean | 0 | FALSE | 0.114 | 2 | 0.886 | | | X |
| f7_disorders | 10 | boolean | 0 | FALSE | 0.005 | 2 | 0.995 | | X | |
| f7_disorders | 30 | boolean | 0 | FALSE | 0.01 | 2 | 0.99 | X | X | |
| f7_disorders | 180 | boolean | 0 | FALSE | 0.021 | 2 | 0.979 | | | |
| f7_disorders | 365 | boolean | 0 | FALSE | 0.026 | 2 | 0.974 | | | |
| f7_disorders | 730 | boolean | 0 | FALSE | 0.03 | 2 | 0.97 | X | | X |
| f8_disorders | 10 | boolean | 0 | FALSE | 0.007 | 2 | 0.993 | | | |
| f8_disorders | 30 | boolean | 0 | FALSE | 0.015 | 2 | 0.985 | | | |
| f8_disorders | 180 | boolean | 0 | FALSE | 0.035 | 2 | 0.965 | | | |
| f8_disorders | 365 | boolean | 0 | FALSE | 0.043 | 2 | 0.957 | | | |
| f8_disorders | 730 | boolean | 0 | FALSE | 0.054 | 2 | 0.946 | X | | X |
| f9_disorders | 10 | boolean | 0 | FALSE | 0.016 | 2 | 0.984 | | | |
| f9_disorders | 30 | boolean | 0 | FALSE | 0.033 | 2 | 0.967 | X | | |
| f9_disorders | 180 | boolean | 0 | FALSE | 0.075 | 2 | 0.925 | | | |
| f9_disorders | 365 | boolean | 0 | FALSE | 0.098 | 2 | 0.902 | | | |
| f9_disorders | 730 | boolean | 0 | FALSE | 0.126 | 2 | 0.874 | X | | X |
| farlighed | 730 | boolean | 0 | FALSE | 0.104 | 2 | 0.896 | X | X | |
| farlighed | 730 | count | 0 | FALSE | 0.234 | 49 | 0.896 | X | X | |
| farlighed | 730 | summed | 0 | FALSE | 52.497 | 996 | 0.898 | X | X | |
| fastholden | 730 | boolean | 0 | FALSE | 0.015 | 2 | 0.985 | X | X | |
| fastholden | 730 | count | 0 | FALSE | 0.025 | 14 | 0.985 | X | X | |
| fastholden | 730 | summed | 0 | FALSE | 0.005 | 35 | 0.989 | X | X | |
| hamilton_d17 | 1 | mean | NaN | FALSE | 18.553 | 58 | 0.987 | X | | |
| hamilton_d17 | 1 | maximum | NaN | FALSE | 18.558 | 47 | 0.987 | X | | |
| hamilton_d17 | 1 | day | NaN | FALSE | -10.971 | 24 | 1 | | | |
| hamilton_d17 | 1 | variance | NaN | FALSE | 7.672 | 10 | 1 | | | |
| hamilton_d17 | 1 | minimum | NaN | FALSE | 18.549 | 47 | 0.987 | X | | |

| | | | | | | | | | |
|---|---|---|---|---|---|---|---|---|---|
| hamilton_d17 | 3 | mean | NaN | FALSE | 19.077 | 77 | 0.963 | | |
| hamilton_d17 | 3 | maximum | NaN | FALSE | 19.101 | 47 | 0.963 | | |
| hamilton_d17 | 3 | minimum | NaN | FALSE | 19.052 | 47 | 0.963 | | |
| hamilton_d17 | 3 | variance | NaN | FALSE | 16.875 | 19 | 1 | | |
| hamilton_d17 | 3 | day | NaN | FALSE | -4.766 | 162 | 1 | | |
| hamilton_d17 | 7 | maximum | NaN | FALSE | 19.754 | 47 | 0.923 | X | |
| hamilton_d17 | 7 | mean | NaN | FALSE | 19.633 | 93 | 0.923 | X | |
| hamilton_d17 | 7 | day | NaN | FALSE | -0.692 | 912 | 0.997 | | |
| hamilton_d17 | 7 | variance | NaN | FALSE | 24.355 | 46 | 0.997 | X | |
| hamilton_d17 | 7 | minimum | NaN | FALSE | 19.511 | 47 | 0.923 | | |
| hamilton_d17 | 10 | maximum | NaN | FALSE | 20.301 | 47 | 0.903 | | |
| hamilton_d17 | 10 | mean | NaN | FALSE | 19.981 | 126 | 0.903 | | |
| hamilton_d17 | 10 | day | NaN | FALSE | -0.422 | 2224 | 0.989 | | |
| hamilton_d17 | 10 | variance | NaN | FALSE | 26.28 | 119 | 0.989 | | |
| hamilton_d17 | 10 | minimum | NaN | FALSE | 19.66 | 47 | 0.903 | | |
| hamilton_d17 | 30 | mean | NaN | FALSE | 20.971 | 289 | 0.835 | X | |
| hamilton_d17 | 30 | maximum | NaN | FALSE | 22.133 | 47 | 0.835 | | |
| hamilton_d17 | 30 | variance | NaN | FALSE | 36.463 | 851 | 0.95 | | |
| hamilton_d17 | 30 | day | NaN | FALSE | -0.192 | 4922 | 0.95 | | |
| hamilton_d17 | 30 | minimum | NaN | FALSE | 19.805 | 47 | 0.835 | X | |
| hamilton_d17 | 180 | mean | NaN | FALSE | 20.656 | 624 | 0.763 | | |
| hamilton_d17 | 180 | maximum | NaN | FALSE | 23.213 | 47 | 0.763 | | |
| hamilton_d17 | 180 | day | NaN | FALSE | -0.086 | 6784 | 0.884 | | |
| hamilton_d17 | 180 | variance | NaN | FALSE | 45.86 | 2116 | 0.884 | | |
| hamilton_d17 | 180 | minimum | NaN | FALSE | 18.009 | 47 | 0.763 | | |
| hamilton_d17 | 365 | maximum | NaN | FALSE | 23.366 | 47 | 0.733 | | |
| hamilton_d17 | 365 | mean | NaN | FALSE | 20.252 | 812 | 0.733 | | |
| hamilton_d17 | 365 | day | NaN | FALSE | -0.066 | 7607 | 0.856 | | |
| hamilton_d17 | 365 | minimum | NaN | FALSE | 17.042 | 46 | 0.733 | | |
| hamilton_d17 | 365 | variance | NaN | FALSE | 46.906 | 2797 | 0.856 | | |
| hamilton_d17 | 730 | mean | NaN | FALSE | 19.722 | 1012 | 0.698 | | |
| hamilton_d17 | 730 | maximum | NaN | FALSE | 23.365 | 47 | 0.698 | X | |
| hamilton_d17 | 730 | variance | NaN | FALSE | 47.396 | 3438 | 0.823 | | |
| hamilton_d17 | 730 | minimum | NaN | FALSE | 15.961 | 45 | 0.698 | X | |
| hamilton_d17 | 730 | day | NaN | FALSE | -0.051 | 8338 | 0.823 | | |
| height_in_cm | 10 | latest | NaN | FALSE | 167.26 | 489 | 0.715 | | X |
| height_in_cm | 365 | latest | NaN | FALSE | 166.57 | 542 | 0.153 | | X |
| height_in_cm | 730 | latest | NaN | FALSE | 166.56 | 553 | 0.101 | X | X |
| height_in_cm | 30 | latest | NaN | FALSE | 166.94 | 496 | 0.494 | X | X |
| height_in_cm | 180 | latest | NaN | FALSE | 166.62 | 527 | 0.226 | | X |
| hypnotics_and_sedatives | 1 | boolean | 0 | FALSE | 0.264 | 2 | 0.736 | | |
| hypnotics_and_sedatives | 1 | count | 0 | FALSE | 0.284 | 14 | 0.736 | | |
| hypnotics_and_sedatives | 3 | boolean | 0 | FALSE | 0.316 | 2 | 0.684 | | |
| hypnotics_and_sedatives | 3 | count | 0 | FALSE | 0.813 | 31 | 0.684 | | X |
| hypnotics_and_sedatives | 7 | boolean | 0 | FALSE | 0.356 | 2 | 0.644 | X | |
| hypnotics_and_sedatives | 7 | count | 0 | FALSE | 1.741 | 64 | 0.644 | X | X |
| hypnotics_and_sedatives | 10 | boolean | 0 | FALSE | 0.374 | 2 | 0.626 | | |
| hypnotics_and_sedatives | 10 | count | 0 | FALSE | 2.349 | 90 | 0.626 | | X |
| hypnotics_and_sedatives | 30 | boolean | 0 | FALSE | 0.434 | 2 | 0.566 | | |
| hypnotics_and_sedatives | 30 | count | 0 | FALSE | 5.192 | 179 | 0.566 | X | X |
| hypnotics_and_sedatives | 180 | boolean | 0 | FALSE | 0.532 | 2 | 0.468 | | X |
| hypnotics_and_sedatives | 180 | count | 0 | FALSE | 12.919 | 393 | 0.468 | | X |
| hypnotics_and_sedatives | 365 | boolean | 0 | FALSE | 0.581 | 2 | 0.419 | | X |
| hypnotics_and_sedatives | 365 | count | 0 | FALSE | 18.472 | 656 | 0.419 | | X |
| hypnotics_and_sedatives | 730 | boolean | 0 | FALSE | 0.636 | 2 | 0.364 | X | X |
| hypnotics_and_sedatives | 730 | count | 0 | FALSE | 26.993 | 1076 | 0.364 | X | X |
| hypnotics_and_sedatives_engangs | 1 | boolean | 0 | FALSE | 0.012 | 2 | 0.988 | X | X |
| hypnotics_and_sedatives_engangs | 1 | count | 0 | FALSE | 0.012 | 6 | 0.988 | X | X |
| hypnotics_and_sedatives_engangs | 3 | count | 0 | FALSE | 0.034 | 6 | 0.972 | | X |
| hypnotics_and_sedatives_engangs | 3 | boolean | 0 | FALSE | 0.028 | 2 | 0.972 | | X |
| hypnotics_and_sedatives_engangs | 7 | boolean | 0 | FALSE | 0.052 | 2 | 0.948 | X | X |
| hypnotics_and_sedatives_engangs | 7 | count | 0 | FALSE | 0.07 | 11 | 0.948 | X | X |
| hypnotics_and_sedatives_engangs | 10 | boolean | 0 | FALSE | 0.066 | 2 | 0.934 | | X |
| hypnotics_and_sedatives_engangs | 10 | count | 0 | FALSE | 0.092 | 12 | 0.934 | | X |
| hypnotics_and_sedatives_engangs | 30 | count | 0 | FALSE | 0.193 | 20 | 0.877 | X | |
| hypnotics_and_sedatives_engangs | 30 | boolean | 0 | FALSE | 0.123 | 2 | 0.877 | X | |
| hypnotics_and_sedatives_engangs | 180 | boolean | 0 | FALSE | 0.23 | 2 | 0.77 | | |
| hypnotics_and_sedatives_engangs | 180 | count | 0 | FALSE | 0.444 | 22 | 0.77 | | |
| hypnotics_and_sedatives_engangs | 365 | count | 0 | FALSE | 0.624 | 24 | 0.715 | | |
| hypnotics_and_sedatives_engangs | 365 | boolean | 0 | FALSE | 0.285 | 2 | 0.715 | | |
| hypnotics_and_sedatives_engangs | 730 | count | 0 | FALSE | 0.921 | 33 | 0.65 | | |
| hypnotics_and_sedatives_engangs | 730 | boolean | 0 | FALSE | 0.35 | 2 | 0.65 | X | |
| hypnotics_and_sedatives_fast | 1 | boolean | 0 | FALSE | 0.171 | 2 | 0.829 | X | X |
| hypnotics_and_sedatives_fast | 1 | count | 0 | FALSE | 0.175 | 8 | 0.829 | X | X |
| hypnotics_and_sedatives_fast | 3 | boolean | 0 | FALSE | 0.183 | 2 | 0.817 | | |
| hypnotics_and_sedatives_fast | 3 | count | 0 | FALSE | 0.5 | 20 | 0.817 | | X |

| | | | | | | | | | |
|---|---|---|---|---|---|---|---|---|---|
| hypnotics_and_sedatives_fast | 7 | count | 0 | FALSE | 1.071 | 43 | 0.808 | X | X |
| hypnotics_and_sedatives_fast | 7 | boolean | 0 | FALSE | 0.192 | 2 | 0.808 | X | X |
| hypnotics_and_sedatives_fast | 10 | count | 0 | FALSE | 1.447 | 54 | 0.803 | | X |
| hypnotics_and_sedatives_fast | 10 | boolean | 0 | FALSE | 0.197 | 2 | 0.803 | | X |
| hypnotics_and_sedatives_fast | 30 | boolean | 0 | FALSE | 0.218 | 2 | 0.782 | X | X |
| hypnotics_and_sedatives_fast | 30 | count | 0 | FALSE | 3.223 | 117 | 0.782 | X | X |
| hypnotics_and_sedatives_fast | 180 | boolean | 0 | FALSE | 0.265 | 2 | 0.735 | | X |
| hypnotics_and_sedatives_fast | 180 | count | 0 | FALSE | 8.176 | 222 | 0.735 | | X |
| hypnotics_and_sedatives_fast | 365 | count | 0 | FALSE | 11.686 | 400 | 0.704 | | X |
| hypnotics_and_sedatives_fast | 365 | boolean | 0 | FALSE | 0.296 | 2 | 0.704 | | X |
| hypnotics_and_sedatives_fast | 730 | count | 0 | FALSE | 16.818 | 771 | 0.666 | X | X |
| hypnotics_and_sedatives_fast | 730 | boolean | 0 | FALSE | 0.334 | 2 | 0.666 | X | X |
| hypnotics_and_sedatives_pn | 1 | count | 0 | FALSE | 0.098 | 13 | 0.906 | X | |
| hypnotics_and_sedatives_pn | 1 | boolean | 0 | FALSE | 0.094 | 2 | 0.906 | X | |
| hypnotics_and_sedatives_pn | 3 | boolean | 0 | FALSE | 0.135 | 2 | 0.865 | | |
| hypnotics_and_sedatives_pn | 3 | count | 0 | FALSE | 0.281 | 26 | 0.865 | | |
| hypnotics_and_sedatives_pn | 7 | count | 0 | FALSE | 0.603 | 52 | 0.833 | X | |
| hypnotics_and_sedatives_pn | 7 | boolean | 0 | FALSE | 0.167 | 2 | 0.833 | X | |
| hypnotics_and_sedatives_pn | 10 | count | 0 | FALSE | 0.813 | 68 | 0.818 | | X |
| hypnotics_and_sedatives_pn | 10 | boolean | 0 | FALSE | 0.182 | 2 | 0.818 | | |
| hypnotics_and_sedatives_pn | 30 | count | 0 | FALSE | 1.784 | 125 | 0.768 | X | X |
| hypnotics_and_sedatives_pn | 30 | boolean | 0 | FALSE | 0.232 | 2 | 0.768 | X | X |
| hypnotics_and_sedatives_pn | 180 | count | 0 | FALSE | 4.321 | 248 | 0.678 | | X |
| hypnotics_and_sedatives_pn | 180 | boolean | 0 | FALSE | 0.322 | 2 | 0.678 | | X |
| hypnotics_and_sedatives_pn | 365 | count | 0 | FALSE | 6.189 | 363 | 0.63 | | X |
| hypnotics_and_sedatives_pn | 365 | boolean | 0 | FALSE | 0.37 | 2 | 0.63 | | X |
| hypnotics_and_sedatives_pn | 730 | count | 0 | FALSE | 9.29 | 536 | 0.568 | X | X |
| hypnotics_and_sedatives_pn | 730 | boolean | 0 | FALSE | 0.432 | 2 | 0.568 | X | X |
| hypnotics_and_sedatives_with_rivotril | 1 | boolean | 0 | FALSE | 0.302 | 2 | 0.698 | X | |
| hypnotics_and_sedatives_with_rivotril | 1 | count | 0 | FALSE | 0.394 | 14 | 0.698 | X | X |
| hypnotics_and_sedatives_with_rivotril | 3 | count | 0 | FALSE | 1.128 | 31 | 0.645 | | |
| hypnotics_and_sedatives_with_rivotril | 3 | boolean | 0 | FALSE | 0.355 | 2 | 0.645 | | |
| hypnotics_and_sedatives_with_rivotril | 7 | count | 0 | FALSE | 2.42 | 64 | 0.606 | X | |
| hypnotics_and_sedatives_with_rivotril | 7 | boolean | 0 | FALSE | 0.394 | 2 | 0.606 | X | |
| hypnotics_and_sedatives_with_rivotril | 10 | count | 0 | FALSE | 3.271 | 90 | 0.588 | | |
| hypnotics_and_sedatives_with_rivotril | 10 | boolean | 0 | FALSE | 0.412 | 2 | 0.588 | | |
| hypnotics_and_sedatives_with_rivotril | 30 | count | 0 | FALSE | 7.303 | 225 | 0.53 | X | |
| hypnotics_and_sedatives_with_rivotril | 30 | boolean | 0 | FALSE | 0.47 | 2 | 0.53 | X | |
| hypnotics_and_sedatives_with_rivotril | 180 | count | 0 | FALSE | 18.599 | 631 | 0.434 | | X |
| hypnotics_and_sedatives_with_rivotril | 180 | boolean | 0 | FALSE | 0.566 | 2 | 0.434 | | |
| hypnotics_and_sedatives_with_rivotril | 365 | count | 0 | FALSE | 27.104 | 1087 | 0.387 | | X |
| hypnotics_and_sedatives_with_rivotril | 365 | boolean | 0 | FALSE | 0.613 | 2 | 0.387 | | X |
| hypnotics_and_sedatives_with_rivotril | 730 | boolean | 0 | FALSE | 0.664 | 2 | 0.336 | | |
| hypnotics_and_sedatives_with_rivotril | 730 | count | 0 | FALSE | 40.875 | 1818 | 0.336 | | |
| hypnotics_and_sedatives_with_rivotril_engangs | 1 | count | 0 | FALSE | 0.013 | 6 | 0.987 | X | X |
| hypnotics_and_sedatives_with_rivotril_engangs | 1 | boolean | 0 | FALSE | 0.013 | 2 | 0.987 | X | X |
| hypnotics_and_sedatives_with_rivotril_engangs | 3 | boolean | 0 | FALSE | 0.031 | 2 | 0.969 | | X |
| hypnotics_and_sedatives_with_rivotril_engangs | 3 | count | 0 | FALSE | 0.037 | 8 | 0.969 | | X |
| hypnotics_and_sedatives_with_rivotril_engangs | 7 | count | 0 | FALSE | 0.077 | 11 | 0.943 | X | X |
| hypnotics_and_sedatives_with_rivotril_engangs | 7 | boolean | 0 | FALSE | 0.057 | 2 | 0.943 | X | X |
| hypnotics_and_sedatives_with_rivotril_engangs | 10 | count | 0 | FALSE | 0.102 | 13 | 0.928 | | X |
| hypnotics_and_sedatives_with_rivotril_engangs | 10 | boolean | 0 | FALSE | 0.072 | 2 | 0.928 | | X |
| hypnotics_and_sedatives_with_rivotril_engangs | 30 | count | 0 | FALSE | 0.214 | 20 | 0.866 | X | |
| hypnotics_and_sedatives_with_rivotril_engangs | 30 | boolean | 0 | FALSE | 0.134 | 2 | 0.866 | X | X |
| hypnotics_and_sedatives_with_rivotril_engangs | 180 | count | 0 | FALSE | 0.492 | 23 | 0.752 | | |
| hypnotics_and_sedatives_with_rivotril_engangs | 180 | boolean | 0 | FALSE | 0.248 | 2 | 0.752 | | |
| hypnotics_and_sedatives_with_rivotril_engangs | 365 | count | 0 | FALSE | 0.695 | 24 | 0.696 | | |
| hypnotics_and_sedatives_with_rivotril_engangs | 365 | boolean | 0 | FALSE | 0.304 | 2 | 0.696 | | |
| hypnotics_and_sedatives_with_rivotril_engangs | 730 | count | 0 | FALSE | 1.036 | 34 | 0.63 | X | |
| hypnotics_and_sedatives_with_rivotril_engangs | 730 | boolean | 0 | FALSE | 0.37 | 2 | 0.63 | X | |
| hypnotics_and_sedatives_with_rivotril_fast | 1 | boolean | 0 | FALSE | 0.208 | 2 | 0.792 | | |
| hypnotics_and_sedatives_with_rivotril_fast | 1 | count | 0 | FALSE | 0.265 | 8 | 0.792 | X | |
| hypnotics_and_sedatives_with_rivotril_fast | 3 | boolean | 0 | FALSE | 0.221 | 2 | 0.779 | | |
| hypnotics_and_sedatives_with_rivotril_fast | 3 | count | 0 | FALSE | 0.759 | 20 | 0.779 | | |
| hypnotics_and_sedatives_with_rivotril_fast | 7 | boolean | 0 | FALSE | 0.231 | 2 | 0.769 | | |
| hypnotics_and_sedatives_with_rivotril_fast | 7 | count | 0 | FALSE | 1.632 | 43 | 0.769 | | |
| hypnotics_and_sedatives_with_rivotril_fast | 10 | boolean | 0 | FALSE | 0.237 | 2 | 0.763 | | |
| hypnotics_and_sedatives_with_rivotril_fast | 10 | count | 0 | FALSE | 2.21 | 58 | 0.763 | | |
| hypnotics_and_sedatives_with_rivotril_fast | 30 | boolean | 0 | FALSE | 0.261 | 2 | 0.739 | X | |
| hypnotics_and_sedatives_with_rivotril_fast | 30 | count | 0 | FALSE | 4.991 | 149 | 0.739 | X | |
| hypnotics_and_sedatives_with_rivotril_fast | 180 | boolean | 0 | FALSE | 0.313 | 2 | 0.687 | | |
| hypnotics_and_sedatives_with_rivotril_fast | 180 | count | 0 | FALSE | 13.048 | 598 | 0.687 | | |
| hypnotics_and_sedatives_with_rivotril_fast | 365 | boolean | 0 | FALSE | 0.347 | 2 | 0.653 | | X |
| hypnotics_and_sedatives_with_rivotril_fast | 365 | count | 0 | FALSE | 19.061 | 1048 | 0.653 | | |
| hypnotics_and_sedatives_with_rivotril_fast | 730 | boolean | 0 | FALSE | 0.388 | 2 | 0.612 | X | |
| hypnotics_and_sedatives_with_rivotril_fast | 730 | count | 0 | FALSE | 28.595 | 1727 | 0.612 | | |

| | | | | | | | | | |
|---|---|---|---|---|---|---|---|---|---|
| hypnotics_and_sedatives_with_rivotril_pn | 1 | count | 0 | FALSE | 0.116 | 13 | 0.895 | X | X |
| hypnotics_and_sedatives_with_rivotril_pn | 1 | boolean | 0 | FALSE | 0.105 | 2 | 0.895 | X | |
| hypnotics_and_sedatives_with_rivotril_pn | 3 | boolean | 0 | FALSE | 0.15 | 2 | 0.85 | | |
| hypnotics_and_sedatives_with_rivotril_pn | 3 | count | 0 | FALSE | 0.333 | 26 | 0.85 | | |
| hypnotics_and_sedatives_with_rivotril_pn | 7 | count | 0 | FALSE | 0.714 | 52 | 0.815 | X | |
| hypnotics_and_sedatives_with_rivotril_pn | 7 | boolean | 0 | FALSE | 0.185 | 2 | 0.815 | X | |
| hypnotics_and_sedatives_with_rivotril_pn | 10 | boolean | 0 | FALSE | 0.201 | 2 | 0.799 | | |
| hypnotics_and_sedatives_with_rivotril_pn | 10 | count | 0 | FALSE | 0.963 | 70 | 0.799 | | |
| hypnotics_and_sedatives_with_rivotril_pn | 30 | boolean | 0 | FALSE | 0.255 | 2 | 0.745 | X | |
| hypnotics_and_sedatives_with_rivotril_pn | 30 | count | 0 | FALSE | 2.109 | 147 | 0.745 | X | |
| hypnotics_and_sedatives_with_rivotril_pn | 180 | boolean | 0 | FALSE | 0.35 | 2 | 0.65 | | |
| hypnotics_and_sedatives_with_rivotril_pn | 180 | count | 0 | FALSE | 5.087 | 260 | 0.65 | | X |
| hypnotics_and_sedatives_with_rivotril_pn | 365 | boolean | 0 | FALSE | 0.399 | 2 | 0.601 | | |
| hypnotics_and_sedatives_with_rivotril_pn | 365 | count | 0 | FALSE | 7.381 | 371 | 0.601 | | X |
| hypnotics_and_sedatives_with_rivotril_pn | 730 | boolean | 0 | FALSE | 0.462 | 2 | 0.538 | X | |
| hypnotics_and_sedatives_with_rivotril_pn | 730 | count | 0 | FALSE | 11.289 | 652 | 0.538 | | |
| lithium | 1 | boolean | 0 | FALSE | 0.072 | 2 | 0.928 | X | |
| lithium | 1 | count | 0 | FALSE | 0.117 | 4 | 0.928 | X | |
| lithium | 3 | count | 0 | FALSE | 0.336 | 10 | 0.927 | | |
| lithium | 3 | boolean | 0 | FALSE | 0.073 | 2 | 0.927 | | |
| lithium | 7 | count | 0 | FALSE | 0.72 | 22 | 0.925 | X | |
| lithium | 7 | boolean | 0 | FALSE | 0.075 | 2 | 0.925 | X | |
| lithium | 10 | boolean | 0 | FALSE | 0.075 | 2 | 0.925 | | |
| lithium | 10 | count | 0 | FALSE | 0.969 | 31 | 0.925 | | |
| lithium | 30 | count | 0 | FALSE | 2.084 | 78 | 0.921 | X | |
| lithium | 30 | boolean | 0 | FALSE | 0.079 | 2 | 0.921 | X | |
| lithium | 180 | count | 0 | FALSE | 5.079 | 350 | 0.911 | | |
| lithium | 180 | boolean | 0 | FALSE | 0.089 | 2 | 0.911 | | |
| lithium | 365 | boolean | 0 | FALSE | 0.095 | 2 | 0.905 | | |
| lithium | 365 | count | 0 | FALSE | 7.555 | 564 | 0.905 | | |
| lithium | 730 | count | 0 | FALSE | 11.66 | 792 | 0.893 | X | |
| lithium | 730 | boolean | 0 | FALSE | 0.107 | 2 | 0.893 | X | |
| manic_and_bipolar | 10 | boolean | 0 | FALSE | 0.04 | 2 | 0.96 | | X |
| manic_and_bipolar | 30 | boolean | 0 | FALSE | 0.081 | 2 | 0.919 | | |
| manic_and_bipolar | 180 | boolean | 0 | FALSE | 0.128 | 2 | 0.872 | | |
| manic_and_bipolar | 365 | boolean | 0 | FALSE | 0.141 | 2 | 0.859 | | |
| manic_and_bipolar | 730 | boolean | 0 | FALSE | 0.154 | 2 | 0.846 | | |
| mas_m | 1 | maximum | NaN | FALSE | 4.345 | 18 | 0.965 | X | X |
| mas_m | 1 | mean | NaN | FALSE | 3.639 | 104 | 0.965 | X | X |
| mas_m | 1 | day | NaN | FALSE | -0.498 | 7074 | 0.98 | X | |
| mas_m | 1 | variance | NaN | FALSE | 4.115 | 239 | 0.98 | X | X |
| mas_m | 1 | minimum | NaN | FALSE | 2.933 | 18 | 0.965 | X | X |
| mas_m | 3 | mean | NaN | FALSE | 3.648 | 433 | 0.955 | | X |
| mas_m | 3 | maximum | NaN | FALSE | 5.174 | 18 | 0.955 | | X |
| mas_m | 3 | variance | NaN | FALSE | 3.957 | 2525 | 0.965 | | X |
| mas_m | 3 | day | NaN | FALSE | -0.451 | 19751 | 0.965 | | |
| mas_m | 3 | minimum | NaN | FALSE | 2.266 | 18 | 0.955 | | X |
| mas_m | 7 | maximum | NaN | FALSE | 6.032 | 18 | 0.947 | X | X |
| mas_m | 7 | mean | NaN | FALSE | 3.786 | 1529 | 0.947 | X | X |
| mas_m | 7 | minimum | NaN | FALSE | 1.949 | 18 | 0.947 | X | X |
| mas_m | 7 | day | NaN | FALSE | -0.35 | 24089 | 0.956 | X | X |
| mas_m | 7 | variance | NaN | FALSE | 4.469 | 9355 | 0.956 | X | X |
| mas_m | 10 | maximum | NaN | FALSE | 6.458 | 18 | 0.944 | | X |
| mas_m | 10 | mean | NaN | FALSE | 3.886 | 2538 | 0.944 | | X |
| mas_m | 10 | day | NaN | FALSE | -0.345 | 25067 | 0.952 | | X |
| mas_m | 10 | minimum | NaN | FALSE | 1.858 | 18 | 0.944 | | X |
| mas_m | 10 | variance | NaN | FALSE | 4.811 | 12936 | 0.952 | | X |
| mas_m | 30 | maximum | NaN | FALSE | 7.538 | 18 | 0.935 | X | X |
| mas_m | 30 | mean | NaN | FALSE | 4.209 | 6672 | 0.935 | X | X |
| mas_m | 30 | minimum | NaN | FALSE | 1.776 | 18 | 0.935 | X | X |
| mas_m | 30 | day | NaN | FALSE | -0.331 | 24754 | 0.944 | X | X |
| mas_m | 30 | variance | NaN | FALSE | 5.859 | 17882 | 0.944 | X | X |
| mas_m | 180 | mean | NaN | FALSE | 4.132 | 8428 | 0.915 | | X |
| mas_m | 180 | maximum | NaN | FALSE | 7.833 | 18 | 0.915 | | X |
| mas_m | 180 | minimum | NaN | FALSE | 1.621 | 18 | 0.915 | | X |
| mas_m | 180 | day | NaN | FALSE | -0.309 | 23283 | 0.927 | | X |
| mas_m | 180 | variance | NaN | FALSE | 6.375 | 18085 | 0.927 | | |
| mas_m | 365 | maximum | NaN | FALSE | 7.893 | 18 | 0.904 | | X |
| mas_m | 365 | mean | NaN | FALSE | 4.052 | 9308 | 0.904 | | X |
| mas_m | 365 | variance | NaN | FALSE | 6.575 | 18856 | 0.917 | | |
| mas_m | 365 | day | NaN | FALSE | -0.301 | 23512 | 0.917 | | X |
| mas_m | 365 | minimum | NaN | FALSE | 1.544 | 18 | 0.904 | | X |
| mas_m | 730 | maximum | NaN | FALSE | 8.065 | 18 | 0.89 | X | X |
| mas_m | 730 | mean | NaN | FALSE | 3.906 | 10679 | 0.89 | X | X |
| mas_m | 730 | minimum | NaN | FALSE | 1.361 | 17 | 0.89 | X | X |
| mas_m | 730 | day | NaN | FALSE | -0.245 | 23744 | 0.904 | X | X |

| | | | | | | | | | |
|---|---|---|---|---|---|---|---|---|---|
| mas_m | 730 | variance | NaN | FALSE | 6.754 | 19854 | 0.904 | X | X |
| medicinering | 10 | count | 0 | FALSE | 0.004 | 3 | 0.996 | | |
| medicinering | 10 | boolean | 0 | FALSE | 0.004 | 2 | 0.996 | | |
| medicinering | 10 | summed | 0 | FALSE | 3.578 | 117 | 0.996 | | |
| medicinering | 30 | boolean | 0 | FALSE | 0.008 | 2 | 0.992 | | |
| medicinering | 30 | count | 0 | FALSE | 0.009 | 4 | 0.992 | | |
| medicinering | 30 | summed | 0 | FALSE | 8.562 | 122 | 0.992 | | |
| medicinering | 180 | boolean | 0 | FALSE | 0.021 | 2 | 0.979 | | |
| medicinering | 180 | count | 0 | FALSE | 0.026 | 7 | 0.979 | | |
| medicinering | 180 | summed | 0 | FALSE | 30.233 | 152 | 0.979 | | |
| medicinering | 365 | count | 0 | FALSE | 0.039 | 8 | 0.973 | | |
| medicinering | 365 | boolean | 0 | FALSE | 0.027 | 2 | 0.973 | | |
| medicinering | 365 | summed | 0 | FALSE | 47.008 | 184 | 0.973 | | |
| medicinering | 730 | boolean | 0 | FALSE | 0.037 | 2 | 0.963 | X | |
| medicinering | 730 | summed | 0 | FALSE | 78.639 | 221 | 0.963 | X | |
| medicinering | 730 | count | 0 | FALSE | 0.063 | 8 | 0.963 | | |
| medicinering | 1 | boolean | 0 | FALSE | 0.025 | 2 | 0.975 | | X |
| medicinering | 3 | boolean | 0 | FALSE | 0.026 | 2 | 0.974 | | X |
| medicinering | 7 | boolean | 0 | FALSE | 0.003 | 2 | 0.997 | X | |
| medicinering | 7 | summed | 0 | FALSE | 2.667 | 117 | 0.997 | X | |
| medicinering | 7 | count | 0 | FALSE | 0.003 | 3 | 0.997 | X | |
| mood_stabilisers | 1 | boolean | 0 | FALSE | 0.157 | 2 | 0.843 | X | |
| mood_stabilisers | 1 | count | 0 | FALSE | 0.226 | 8 | 0.843 | X | |
| mood_stabilisers | 3 | count | 0 | FALSE | 0.64 | 18 | 0.837 | | |
| mood_stabilisers | 3 | boolean | 0 | FALSE | 0.163 | 2 | 0.837 | | |
| mood_stabilisers | 7 | count | 0 | FALSE | 1.355 | 37 | 0.833 | X | |
| mood_stabilisers | 7 | boolean | 0 | FALSE | 0.167 | 2 | 0.833 | X | |
| mood_stabilisers | 10 | count | 0 | FALSE | 1.815 | 52 | 0.831 | | |
| mood_stabilisers | 10 | boolean | 0 | FALSE | 0.169 | 2 | 0.831 | | |
| mood_stabilisers | 30 | boolean | 0 | FALSE | 0.179 | 2 | 0.821 | X | |
| mood_stabilisers | 30 | count | 0 | FALSE | 3.921 | 146 | 0.821 | X | X |
| mood_stabilisers | 180 | boolean | 0 | FALSE | 0.2 | 2 | 0.8 | | |
| mood_stabilisers | 180 | count | 0 | FALSE | 9.945 | 456 | 0.8 | | X |
| mood_stabilisers | 365 | count | 0 | FALSE | 14.849 | 709 | 0.784 | | |
| mood_stabilisers | 365 | boolean | 0 | FALSE | 0.216 | 2 | 0.784 | | |
| mood_stabilisers | 730 | count | 0 | FALSE | 23.201 | 1040 | 0.759 | X | |
| mood_stabilisers | 730 | boolean | 0 | FALSE | 0.241 | 2 | 0.759 | X | |
| nervous_system_stimulants | 1 | boolean | 0 | FALSE | 0.039 | 2 | 0.961 | | |
| nervous_system_stimulants | 1 | count | 0 | FALSE | 0.062 | 8 | 0.961 | | |
| nervous_system_stimulants | 3 | boolean | 0 | FALSE | 0.04 | 2 | 0.96 | | |
| nervous_system_stimulants | 3 | count | 0 | FALSE | 0.174 | 21 | 0.96 | | |
| nervous_system_stimulants | 7 | boolean | 0 | FALSE | 0.041 | 2 | 0.959 | | |
| nervous_system_stimulants | 7 | count | 0 | FALSE | 0.371 | 45 | 0.959 | X | |
| nervous_system_stimulants | 10 | boolean | 0 | FALSE | 0.042 | 2 | 0.958 | | |
| nervous_system_stimulants | 10 | count | 0 | FALSE | 0.5 | 63 | 0.958 | | |
| nervous_system_stimulants | 30 | boolean | 0 | FALSE | 0.045 | 2 | 0.955 | | |
| nervous_system_stimulants | 30 | count | 0 | FALSE | 1.152 | 172 | 0.955 | X | |
| nervous_system_stimulants | 180 | boolean | 0 | FALSE | 0.051 | 2 | 0.949 | | |
| nervous_system_stimulants | 180 | count | 0 | FALSE | 3.35 | 679 | 0.949 | | |
| nervous_system_stimulants | 365 | boolean | 0 | FALSE | 0.056 | 2 | 0.944 | | |
| nervous_system_stimulants | 365 | count | 0 | FALSE | 4.717 | 920 | 0.944 | | |
| nervous_system_stimulants | 730 | boolean | 0 | FALSE | 0.064 | 2 | 0.936 | | |
| nervous_system_stimulants | 730 | count | 0 | FALSE | 6.768 | 1182 | 0.936 | | |
| no_temporary_leave | 1 | count | 0 | FALSE | 0.051 | 5 | 0.951 | X | X |
| no_temporary_leave | 1 | boolean | 0 | FALSE | 0.049 | 2 | 0.951 | X | X |
| no_temporary_leave | 3 | count | 0 | FALSE | 0.14 | 6 | 0.883 | | X |
| no_temporary_leave | 3 | boolean | 0 | FALSE | 0.117 | 2 | 0.883 | | X |
| no_temporary_leave | 7 | boolean | 0 | FALSE | 0.215 | 2 | 0.785 | X | X |
| no_temporary_leave | 7 | count | 0 | FALSE | 0.281 | 7 | 0.785 | X | X |
| no_temporary_leave | 10 | count | 0 | FALSE | 0.366 | 8 | 0.73 | | X |
| no_temporary_leave | 10 | boolean | 0 | FALSE | 0.27 | 2 | 0.73 | | X |
| no_temporary_leave | 30 | boolean | 0 | FALSE | 0.475 | 2 | 0.525 | X | X |
| no_temporary_leave | 30 | count | 0 | FALSE | 0.744 | 13 | 0.525 | X | X |
| no_temporary_leave | 180 | boolean | 0 | FALSE | 0.711 | 2 | 0.289 | | X |
| no_temporary_leave | 180 | count | 0 | FALSE | 1.718 | 39 | 0.289 | | X |
| no_temporary_leave | 365 | count | 0 | FALSE | 2.385 | 42 | 0.24 | | |
| no_temporary_leave | 365 | boolean | 0 | FALSE | 0.76 | 2 | 0.24 | | X |
| no_temporary_leave | 730 | boolean | 0 | FALSE | 0.796 | 2 | 0.204 | X | X |
| no_temporary_leave | 730 | count | 0 | FALSE | 3.361 | 65 | 0.204 | X | |
| non_sedative_antipsychotics | 1 | boolean | 0 | FALSE | 0.401 | 2 | 0.599 | X | X |
| non_sedative_antipsychotics | 1 | count | 0 | FALSE | 0.633 | 11 | 0.599 | | |
| non_sedative_antipsychotics | 3 | count | 0 | FALSE | 1.814 | 27 | 0.576 | | X |
| non_sedative_antipsychotics | 3 | boolean | 0 | FALSE | 0.424 | 2 | 0.576 | | |
| non_sedative_antipsychotics | 7 | count | 0 | FALSE | 3.903 | 59 | 0.553 | X | X |
| non_sedative_antipsychotics | 7 | boolean | 0 | FALSE | 0.447 | 2 | 0.553 | X | |
| non_sedative_antipsychotics | 10 | boolean | 0 | FALSE | 0.459 | 2 | 0.541 | | |

| | | | | | | | | | |
|---|---|---|---|---|---|---|---|---|---|
| non_sedative_antipsychotics | 10 | count | 0 | FALSE | 5.286 | 83 | 0.541 | | X |
| non_sedative_antipsychotics | 30 | boolean | 0 | FALSE | 0.502 | 2 | 0.498 | X | |
| non_sedative_antipsychotics | 30 | count | 0 | FALSE | 12.069 | 212 | 0.498 | X | X |
| non_sedative_antipsychotics | 180 | boolean | 0 | FALSE | 0.557 | 2 | 0.443 | | |
| non_sedative_antipsychotics | 180 | count | 0 | FALSE | 34.962 | 848 | 0.443 | | X |
| non_sedative_antipsychotics | 365 | boolean | 0 | FALSE | 0.585 | 2 | 0.415 | | |
| non_sedative_antipsychotics | 365 | count | 0 | FALSE | 53.741 | 1321 | 0.415 | | X |
| non_sedative_antipsychotics | 730 | boolean | 0 | FALSE | 0.619 | 2 | 0.381 | X | |
| non_sedative_antipsychotics | 730 | count | 0 | FALSE | 84.728 | 1953 | 0.381 | X | X |
| non_sedative_antipsychotics_engangs | 1 | count | 0 | FALSE | 0.005 | 4 | 0.995 | X | X |
| non_sedative_antipsychotics_engangs | 1 | boolean | 0 | FALSE | 0.005 | 2 | 0.995 | X | X |
| non_sedative_antipsychotics_engangs | 3 | count | 0 | FALSE | 0.014 | 7 | 0.987 | | X |
| non_sedative_antipsychotics_engangs | 3 | boolean | 0 | FALSE | 0.013 | 2 | 0.987 | | X |
| non_sedative_antipsychotics_engangs | 7 | count | 0 | FALSE | 0.03 | 9 | 0.974 | X | X |
| non_sedative_antipsychotics_engangs | 7 | boolean | 0 | FALSE | 0.026 | 2 | 0.974 | X | X |
| non_sedative_antipsychotics_engangs | 10 | boolean | 0 | FALSE | 0.034 | 2 | 0.966 | | X |
| non_sedative_antipsychotics_engangs | 10 | count | 0 | FALSE | 0.039 | 9 | 0.966 | | X |
| non_sedative_antipsychotics_engangs | 30 | count | 0 | FALSE | 0.085 | 12 | 0.933 | X | X |
| non_sedative_antipsychotics_engangs | 30 | boolean | 0 | FALSE | 0.067 | 2 | 0.933 | X | X |
| non_sedative_antipsychotics_engangs | 180 | count | 0 | FALSE | 0.202 | 16 | 0.863 | | |
| non_sedative_antipsychotics_engangs | 180 | boolean | 0 | FALSE | 0.137 | 2 | 0.863 | | |
| non_sedative_antipsychotics_engangs | 365 | count | 0 | FALSE | 0.292 | 20 | 0.822 | | |
| non_sedative_antipsychotics_engangs | 365 | boolean | 0 | FALSE | 0.178 | 2 | 0.822 | | |
| non_sedative_antipsychotics_engangs | 730 | count | 0 | FALSE | 0.462 | 26 | 0.761 | X | |
| non_sedative_antipsychotics_engangs | 730 | boolean | 0 | FALSE | 0.239 | 2 | 0.761 | X | |
| non_sedative_antipsychotics_fast | 1 | boolean | 0 | FALSE | 0.383 | 2 | 0.617 | X | X |
| non_sedative_antipsychotics_fast | 1 | count | 0 | FALSE | 0.565 | 9 | 0.617 | X | X |
| non_sedative_antipsychotics_fast | 3 | count | 0 | FALSE | 1.619 | 22 | 0.599 | | X |
| non_sedative_antipsychotics_fast | 3 | boolean | 0 | FALSE | 0.401 | 2 | 0.599 | | X |
| non_sedative_antipsychotics_fast | 7 | count | 0 | FALSE | 3.489 | 48 | 0.581 | X | X |
| non_sedative_antipsychotics_fast | 7 | boolean | 0 | FALSE | 0.419 | 2 | 0.581 | X | X |
| non_sedative_antipsychotics_fast | 10 | boolean | 0 | FALSE | 0.43 | 2 | 0.57 | | X |
| non_sedative_antipsychotics_fast | 10 | count | 0 | FALSE | 4.728 | 65 | 0.57 | | X |
| non_sedative_antipsychotics_fast | 30 | boolean | 0 | FALSE | 0.469 | 2 | 0.531 | X | X |
| non_sedative_antipsychotics_fast | 30 | count | 0 | FALSE | 10.83 | 172 | 0.531 | X | X |
| non_sedative_antipsychotics_fast | 180 | boolean | 0 | FALSE | 0.518 | 2 | 0.482 | | X |
| non_sedative_antipsychotics_fast | 180 | count | 0 | FALSE | 31.695 | 714 | 0.482 | | X |
| non_sedative_antipsychotics_fast | 365 | boolean | 0 | FALSE | 0.543 | 2 | 0.457 | | |
| non_sedative_antipsychotics_fast | 365 | count | 0 | FALSE | 48.775 | 1087 | 0.457 | | X |
| non_sedative_antipsychotics_fast | 730 | boolean | 0 | FALSE | 0.573 | 2 | 0.427 | X | |
| non_sedative_antipsychotics_fast | 730 | count | 0 | FALSE | 77.149 | 1789 | 0.427 | X | X |
| non_sedative_antipsychotics_im | 1 | count | 0 | FALSE | 0.006 | 4 | 0.994 | X | |
| non_sedative_antipsychotics_im | 1 | boolean | 0 | FALSE | 0.006 | 2 | 0.994 | X | |
| non_sedative_antipsychotics_im | 3 | boolean | 0 | FALSE | 0.016 | 2 | 0.984 | | |
| non_sedative_antipsychotics_im | 3 | count | 0 | FALSE | 0.018 | 9 | 0.984 | | |
| non_sedative_antipsychotics_im | 7 | boolean | 0 | FALSE | 0.034 | 2 | 0.966 | X | |
| non_sedative_antipsychotics_im | 7 | count | 0 | FALSE | 0.039 | 15 | 0.966 | X | |
| non_sedative_antipsychotics_im | 10 | count | 0 | FALSE | 0.053 | 21 | 0.956 | | |
| non_sedative_antipsychotics_im | 10 | boolean | 0 | FALSE | 0.044 | 2 | 0.956 | | |
| non_sedative_antipsychotics_im | 30 | count | 0 | FALSE | 0.128 | 52 | 0.922 | | |
| non_sedative_antipsychotics_im | 30 | boolean | 0 | FALSE | 0.078 | 2 | 0.922 | X | |
| non_sedative_antipsychotics_im | 180 | count | 0 | FALSE | 0.45 | 67 | 0.895 | | |
| non_sedative_antipsychotics_im | 180 | boolean | 0 | FALSE | 0.105 | 2 | 0.895 | | |
| non_sedative_antipsychotics_im | 365 | count | 0 | FALSE | 0.776 | 77 | 0.878 | | |
| non_sedative_antipsychotics_im | 365 | boolean | 0 | FALSE | 0.122 | 2 | 0.878 | | |
| non_sedative_antipsychotics_im | 730 | count | 0 | FALSE | 1.404 | 110 | 0.851 | X | |
| non_sedative_antipsychotics_im | 730 | boolean | 0 | FALSE | 0.149 | 2 | 0.851 | X | |
| non_sedative_antipsychotics_pn | 1 | boolean | 0 | FALSE | 0.039 | 2 | 0.961 | X | |
| non_sedative_antipsychotics_pn | 1 | count | 0 | FALSE | 0.062 | 8 | 0.961 | X | |
| non_sedative_antipsychotics_pn | 3 | boolean | 0 | FALSE | 0.056 | 2 | 0.944 | | |
| non_sedative_antipsychotics_pn | 3 | count | 0 | FALSE | 0.178 | 18 | 0.944 | | |
| non_sedative_antipsychotics_pn | 7 | boolean | 0 | FALSE | 0.071 | 2 | 0.929 | X | |
| non_sedative_antipsychotics_pn | 7 | count | 0 | FALSE | 0.378 | 38 | 0.929 | X | |
| non_sedative_antipsychotics_pn | 10 | boolean | 0 | FALSE | 0.077 | 2 | 0.923 | | |
| non_sedative_antipsychotics_pn | 10 | count | 0 | FALSE | 0.509 | 49 | 0.923 | | |
| non_sedative_antipsychotics_pn | 30 | boolean | 0 | FALSE | 0.101 | 2 | 0.899 | X | |
| non_sedative_antipsychotics_pn | 30 | count | 0 | FALSE | 1.135 | 132 | 0.899 | X | |
| non_sedative_antipsychotics_pn | 180 | boolean | 0 | FALSE | 0.148 | 2 | 0.852 | | |
| non_sedative_antipsychotics_pn | 180 | count | 0 | FALSE | 3.02 | 449 | 0.852 | | |
| non_sedative_antipsychotics_pn | 365 | boolean | 0 | FALSE | 0.178 | 2 | 0.822 | | |
| non_sedative_antipsychotics_pn | 365 | count | 0 | FALSE | 4.607 | 625 | 0.822 | | |
| non_sedative_antipsychotics_pn | 730 | boolean | 0 | FALSE | 0.221 | 2 | 0.779 | X | |
| non_sedative_antipsychotics_pn | 730 | count | 0 | FALSE | 7.017 | 793 | 0.779 | | |
| non_sedative_antipsychotics_po | 1 | boolean | 0 | FALSE | 0.001 | 2 | 0.999 | X | |
| non_sedative_antipsychotics_po | 1 | count | 0 | FALSE | 0.001 | 4 | 0.999 | X | |
| non_sedative_antipsychotics_po | 3 | count | 0 | FALSE | 0.004 | 10 | 0.999 | | |

| | | | | | | | | |
|---|---|---|---|---|---|---|---|---|
| non_sedative_antipsychotics_po | 3 | boolean | 0 | FALSE | 0.001 | 2 | 0.999 | |
| non_sedative_antipsychotics_po | 7 | boolean | 0 | FALSE | 0.001 | 2 | 0.999 | X |
| non_sedative_antipsychotics_po | 7 | count | 0 | FALSE | 0.008 | 22 | 0.999 | |
| non_sedative_antipsychotics_po | 10 | count | 0 | FALSE | 0.011 | 31 | 0.999 | |
| non_sedative_antipsychotics_po | 10 | boolean | 0 | FALSE | 0.001 | 2 | 0.999 | |
| non_sedative_antipsychotics_po | 30 | count | 0 | FALSE | 0.027 | 91 | 0.999 | |
| non_sedative_antipsychotics_po | 30 | boolean | 0 | FALSE | 0.001 | 2 | 0.999 | X |
| non_sedative_antipsychotics_po | 180 | count | 0 | FALSE | 0.049 | 174 | 0.999 | |
| non_sedative_antipsychotics_po | 180 | boolean | 0 | FALSE | 0.001 | 2 | 0.999 | |
| non_sedative_antipsychotics_po | 365 | count | 0 | FALSE | 0.06 | 181 | 0.998 | |
| non_sedative_antipsychotics_po | 365 | boolean | 0 | FALSE | 0.002 | 2 | 0.998 | |
| non_sedative_antipsychotics_po | 730 | boolean | 0 | FALSE | 0.002 | 2 | 0.998 | X |
| non_sedative_antipsychotics_po | 730 | count | 0 | FALSE | 0.108 | 231 | 0.998 | X |
| opioid_dependence | 1 | boolean | 0 | FALSE | 0.035 | 2 | 0.965 | |
| opioid_dependence | 1 | count | 0 | FALSE | 0.051 | 9 | 0.965 | |
| opioid_dependence | 3 | boolean | 0 | FALSE | 0.036 | 2 | 0.964 | |
| opioid_dependence | 3 | count | 0 | FALSE | 0.146 | 20 | 0.964 | |
| opioid_dependence | 7 | boolean | 0 | FALSE | 0.036 | 2 | 0.964 | |
| opioid_dependence | 7 | count | 0 | FALSE | 0.315 | 40 | 0.964 | X |
| opioid_dependence | 10 | boolean | 0 | FALSE | 0.036 | 2 | 0.964 | |
| opioid_dependence | 10 | count | 0 | FALSE | 0.431 | 54 | 0.964 | |
| opioid_dependence | 30 | boolean | 0 | FALSE | 0.038 | 2 | 0.962 | |
| opioid_dependence | 30 | count | 0 | FALSE | 1.063 | 137 | 0.962 | X |
| opioid_dependence | 180 | boolean | 0 | FALSE | 0.041 | 2 | 0.959 | |
| opioid_dependence | 180 | count | 0 | FALSE | 3.96 | 503 | 0.959 | |
| opioid_dependence | 365 | count | 0 | FALSE | 6.278 | 900 | 0.957 | |
| opioid_dependence | 365 | boolean | 0 | FALSE | 0.043 | 2 | 0.957 | |
| opioid_dependence | 730 | boolean | 0 | FALSE | 0.046 | 2 | 0.954 | X |
| opioid_dependence | 730 | count | 0 | FALSE | 9.571 | 1547 | 0.954 | X |
| outcome_all_restraint | 2 | boolean | 0 | FALSE | 0.004 | 2 | 0.996 | |
| outcome_chemical_restraint | 2 | boolean | 0 | FALSE | 0.003 | 2 | 0.997 | |
| outcome_manual_restraint | 2 | boolean | 0 | FALSE | 0.001 | 2 | 0.999 | |
| outcome_mechanical_restraint | 2 | boolean | 0 | FALSE | 0.001 | 2 | 0.999 | |
| p_aripiprazol | 10 | maximum | NaN | FALSE | 562.46 | 342 | 0.995 | |
| p_aripiprazol | 10 | latest | NaN | FALSE | 561.76 | 343 | 0.995 | |
| p_aripiprazol | 10 | minimum | NaN | FALSE | 560.69 | 342 | 0.995 | |
| p_aripiprazol | 10 | mean | NaN | FALSE | 561.58 | 352 | 0.995 | |
| p_aripiprazol | 30 | mean | NaN | FALSE | 553.18 | 427 | 0.99 | |
| p_aripiprazol | 30 | minimum | NaN | FALSE | 546.43 | 382 | 0.99 | |
| p_aripiprazol | 30 | maximum | NaN | FALSE | 560 | 386 | 0.99 | |
| p_aripiprazol | 30 | latest | NaN | FALSE | 554.58 | 389 | 0.99 | |
| p_aripiprazol | 180 | mean | NaN | FALSE | 567.07 | 594 | 0.974 | |
| p_aripiprazol | 180 | minimum | NaN | FALSE | 543.79 | 493 | 0.974 | |
| p_aripiprazol | 180 | maximum | NaN | FALSE | 591.99 | 497 | 0.974 | |
| p_aripiprazol | 180 | latest | NaN | FALSE | 573.25 | 523 | 0.974 | |
| p_aripiprazol | 365 | maximum | NaN | FALSE | 593.31 | 547 | 0.963 | |
| p_aripiprazol | 365 | minimum | NaN | FALSE | 516.29 | 525 | 0.963 | |
| p_aripiprazol | 365 | mean | NaN | FALSE | 553.06 | 690 | 0.963 | |
| p_aripiprazol | 365 | latest | NaN | FALSE | 557.13 | 578 | 0.963 | |
| p_aripiprazol | 730 | minimum | NaN | FALSE | 481.79 | 570 | 0.95 | |
| p_aripiprazol | 730 | maximum | NaN | FALSE | 593.81 | 593 | 0.95 | X |
| p_aripiprazol | 730 | mean | NaN | FALSE | 536.4 | 787 | 0.95 | |
| p_aripiprazol | 730 | latest | NaN | FALSE | 539.24 | 635 | 0.95 | |
| p_clomipramine | 10 | maximum | NaN | FALSE | 353.77 | 365 | 0.994 | |
| p_clomipramine | 10 | mean | NaN | FALSE | 347.59 | 402 | 0.994 | |
| p_clomipramine | 10 | minimum | NaN | FALSE | 341.41 | 365 | 0.994 | |
| p_clomipramine | 10 | latest | NaN | FALSE | 344.78 | 370 | 0.994 | |
| p_clomipramine | 30 | maximum | NaN | FALSE | 368.22 | 393 | 0.988 | |
| p_clomipramine | 30 | latest | NaN | FALSE | 342.94 | 403 | 0.988 | |
| p_clomipramine | 30 | minimum | NaN | FALSE | 317.39 | 371 | 0.988 | |
| p_clomipramine | 30 | mean | NaN | FALSE | 342.01 | 504 | 0.988 | |
| p_clomipramine | 180 | minimum | NaN | FALSE | 264.05 | 381 | 0.977 | |
| p_clomipramine | 180 | maximum | NaN | FALSE | 402.57 | 438 | 0.977 | |
| p_clomipramine | 180 | mean | NaN | FALSE | 329.64 | 674 | 0.977 | |
| p_clomipramine | 180 | latest | NaN | FALSE | 338.01 | 477 | 0.977 | |
| p_clomipramine | 365 | mean | NaN | FALSE | 325.98 | 748 | 0.971 | |
| p_clomipramine | 365 | latest | NaN | FALSE | 334.62 | 503 | 0.971 | |
| p_clomipramine | 365 | minimum | NaN | FALSE | 239.77 | 377 | 0.971 | |
| p_clomipramine | 365 | maximum | NaN | FALSE | 425.67 | 460 | 0.971 | |
| p_clomipramine | 730 | maximum | NaN | FALSE | 454.9 | 463 | 0.963 | |
| p_clomipramine | 730 | mean | NaN | FALSE | 324.64 | 813 | 0.963 | X |
| p_clomipramine | 730 | minimum | NaN | FALSE | 218.36 | 360 | 0.963 | X |
| p_clomipramine | 730 | latest | NaN | FALSE | 327.27 | 520 | 0.963 | X |
| p_clozapine | 10 | minimum | NaN | FALSE | 1225.2 | 1354 | 0.978 | |
| p_clozapine | 10 | mean | NaN | FALSE | 1246.3 | 1672 | 0.978 | |
| p_clozapine | 10 | maximum | NaN | FALSE | 1268 | 1363 | 0.978 | |

| | | | | | | | | |
|---|---|---|---|---|---|---|---|---|
| p_clozapine | 10 | latest | NaN | FALSE | 1245.4 | 1372 | 0.978 | |
| p_clozapine | 30 | maximum | NaN | FALSE | 1340.8 | 1411 | 0.959 | X |
| p_clozapine | 30 | latest | NaN | FALSE | 1243 | 1501 | 0.959 | X |
| p_clozapine | 30 | mean | NaN | FALSE | 1245.7 | 2209 | 0.959 | X |
| p_clozapine | 30 | minimum | NaN | FALSE | 1154.2 | 1354 | 0.959 | X |
| p_clozapine | 180 | minimum | NaN | FALSE | 947.43 | 1192 | 0.926 | |
| p_clozapine | 180 | mean | NaN | FALSE | 1220.1 | 3087 | 0.926 | |
| p_clozapine | 180 | maximum | NaN | FALSE | 1517.8 | 1414 | 0.926 | |
| p_clozapine | 180 | latest | NaN | FALSE | 1225.4 | 1711 | 0.926 | |
| p_clozapine | 365 | latest | NaN | FALSE | 1191.5 | 1771 | 0.915 | |
| p_clozapine | 365 | minimum | NaN | FALSE | 827.5 | 1079 | 0.915 | |
| p_clozapine | 365 | mean | NaN | FALSE | 1184.5 | 3448 | 0.915 | |
| p_clozapine | 365 | maximum | NaN | FALSE | 1592.9 | 1356 | 0.915 | |
| p_clozapine | 730 | maximum | NaN | FALSE | 1677.2 | 1242 | 0.906 | X |
| p_clozapine | 730 | minimum | NaN | FALSE | 674.51 | 942 | 0.906 | X |
| p_clozapine | 730 | latest | NaN | FALSE | 1149.7 | 1794 | 0.906 | X |
| p_clozapine | 730 | mean | NaN | FALSE | 1130.2 | 3726 | 0.906 | X |
| p_ethanol | 10 | maximum | NaN | FALSE | 34.898 | 143 | 0.992 | |
| p_ethanol | 10 | latest | NaN | FALSE | 33.529 | 139 | 0.992 | |
| p_ethanol | 10 | mean | NaN | FALSE | 33.945 | 213 | 0.992 | |
| p_ethanol | 10 | minimum | NaN | FALSE | 32.999 | 138 | 0.992 | |
| p_ethanol | 30 | minimum | NaN | FALSE | 32.472 | 139 | 0.984 | X |
| p_ethanol | 30 | mean | NaN | FALSE | 34.051 | 277 | 0.984 | X |
| p_ethanol | 30 | maximum | NaN | FALSE | 35.637 | 149 | 0.984 | X |
| p_ethanol | 30 | latest | NaN | FALSE | 33.502 | 140 | 0.984 | X |
| p_ethanol | 180 | latest | NaN | FALSE | 32.374 | 151 | 0.964 | |
| p_ethanol | 180 | mean | NaN | FALSE | 32.609 | 522 | 0.964 | |
| p_ethanol | 180 | minimum | NaN | FALSE | 29.032 | 146 | 0.964 | |
| p_ethanol | 180 | maximum | NaN | FALSE | 35.991 | 159 | 0.964 | |
| p_ethanol | 365 | minimum | NaN | FALSE | 27.175 | 149 | 0.95 | |
| p_ethanol | 365 | maximum | NaN | FALSE | 36.393 | 166 | 0.95 | |
| p_ethanol | 365 | mean | NaN | FALSE | 31.947 | 656 | 0.95 | |
| p_ethanol | 365 | latest | NaN | FALSE | 32.035 | 158 | 0.95 | |
| p_ethanol | 730 | minimum | NaN | FALSE | 25.081 | 154 | 0.93 | X |
| p_ethanol | 730 | maximum | NaN | FALSE | 35.59 | 170 | 0.93 | |
| p_ethanol | 730 | latest | NaN | FALSE | 30.718 | 165 | 0.93 | X |
| p_ethanol | 730 | mean | NaN | FALSE | 30.544 | 822 | 0.93 | |
| p_haloperidol | 10 | mean | NaN | FALSE | 21.652 | 30 | 1 | |
| p_haloperidol | 10 | latest | NaN | FALSE | 21.703 | 29 | 1 | |
| p_haloperidol | 10 | maximum | NaN | FALSE | 21.703 | 29 | 1 | |
| p_haloperidol | 10 | minimum | NaN | FALSE | 21.6 | 29 | 1 | |
| p_haloperidol | 30 | maximum | NaN | FALSE | 20.393 | 30 | 0.999 | |
| p_haloperidol | 30 | minimum | NaN | FALSE | 19.089 | 29 | 0.999 | |
| p_haloperidol | 30 | latest | NaN | FALSE | 20.11 | 32 | 0.999 | |
| p_haloperidol | 30 | mean | NaN | FALSE | 19.741 | 35 | 0.999 | |
| p_haloperidol | 180 | minimum | NaN | FALSE | 13.942 | 32 | 0.998 | |
| p_haloperidol | 180 | maximum | NaN | FALSE | 19.277 | 32 | 0.998 | |
| p_haloperidol | 180 | mean | NaN | FALSE | 16.427 | 54 | 0.998 | |
| p_haloperidol | 180 | latest | NaN | FALSE | 17.628 | 37 | 0.998 | |
| p_haloperidol | 365 | mean | NaN | FALSE | 17.56 | 67 | 0.997 | |
| p_haloperidol | 365 | latest | NaN | FALSE | 18.486 | 41 | 0.997 | |
| p_haloperidol | 365 | minimum | NaN | FALSE | 15.017 | 32 | 0.997 | |
| p_haloperidol | 365 | maximum | NaN | FALSE | 20.429 | 34 | 0.997 | |
| p_haloperidol | 730 | maximum | NaN | FALSE | 18.876 | 40 | 0.994 | X |
| p_haloperidol | 730 | minimum | NaN | FALSE | 13.344 | 37 | 0.994 | X |
| p_haloperidol | 730 | mean | NaN | FALSE | 15.983 | 76 | 0.994 | X |
| p_haloperidol | 730 | latest | NaN | FALSE | 17.118 | 45 | 0.994 | X |
| p_lithium | 10 | minimum | NaN | FALSE | 0.6 | 144 | 0.956 | |
| p_lithium | 10 | maximum | NaN | FALSE | 0.67 | 180 | 0.956 | |
| p_lithium | 10 | mean | NaN | FALSE | 0.635 | 717 | 0.956 | |
| p_lithium | 10 | latest | NaN | FALSE | 0.631 | 151 | 0.956 | |
| p_lithium | 30 | mean | NaN | FALSE | 0.63 | 1253 | 0.936 | X |
| p_lithium | 30 | maximum | NaN | FALSE | 0.726 | 185 | 0.936 | X |
| p_lithium | 30 | latest | NaN | FALSE | 0.633 | 151 | 0.936 | X |
| p_lithium | 30 | minimum | NaN | FALSE | 0.534 | 137 | 0.936 | X |
| p_lithium | 180 | minimum | NaN | FALSE | 0.407 | 113 | 0.912 | |
| p_lithium | 180 | maximum | NaN | FALSE | 0.817 | 195 | 0.912 | |
| p_lithium | 180 | latest | NaN | FALSE | 0.613 | 152 | 0.912 | |
| p_lithium | 180 | mean | NaN | FALSE | 0.61 | 2395 | 0.912 | |
| p_lithium | 365 | minimum | NaN | FALSE | 0.357 | 104 | 0.901 | |
| p_lithium | 365 | latest | NaN | FALSE | 0.597 | 153 | 0.901 | |
| p_lithium | 365 | maximum | NaN | FALSE | 0.862 | 204 | 0.901 | |
| p_lithium | 365 | mean | NaN | FALSE | 0.602 | 3218 | 0.901 | |
| p_lithium | 730 | maximum | NaN | FALSE | 0.907 | 211 | 0.887 | |
| p_lithium | 730 | minimum | NaN | FALSE | 0.308 | 98 | 0.887 | X |
| p_lithium | 730 | latest | NaN | FALSE | 0.577 | 155 | 0.887 | X |

| | | | | | | | | |
|---|---|---|---|---|---|---|---|---|
| p_lithium | 730 | mean | NaN | FALSE | 0.591 | 4027 | 0.887 | X |
| p_nortriptyline | 10 | minimum | NaN | FALSE | 414.49 | 609 | 0.987 | |
| p_nortriptyline | 10 | maximum | NaN | FALSE | 423.62 | 613 | 0.987 | |
| p_nortriptyline | 10 | latest | NaN | FALSE | 417.62 | 615 | 0.987 | |
| p_nortriptyline | 10 | mean | NaN | FALSE | 419.04 | 686 | 0.987 | |
| p_nortriptyline | 30 | latest | NaN | FALSE | 422.3 | 654 | 0.974 | |
| p_nortriptyline | 30 | maximum | NaN | FALSE | 450.45 | 647 | 0.974 | |
| p_nortriptyline | 30 | minimum | NaN | FALSE | 400.18 | 611 | 0.974 | |
| p_nortriptyline | 30 | mean | NaN | FALSE | 425.16 | 873 | 0.974 | |
| p_nortriptyline | 180 | maximum | NaN | FALSE | 499.05 | 680 | 0.952 | |
| p_nortriptyline | 180 | minimum | NaN | FALSE | 346.15 | 603 | 0.952 | |
| p_nortriptyline | 180 | latest | NaN | FALSE | 416.67 | 705 | 0.952 | |
| p_nortriptyline | 180 | mean | NaN | FALSE | 416.87 | 1198 | 0.952 | |
| p_nortriptyline | 365 | mean | NaN | FALSE | 411.16 | 1348 | 0.943 | |
| p_nortriptyline | 365 | maximum | NaN | FALSE | 517.44 | 696 | 0.943 | |
| p_nortriptyline | 365 | minimum | NaN | FALSE | 318.87 | 584 | 0.943 | |
| p_nortriptyline | 365 | latest | NaN | FALSE | 414.52 | 728 | 0.943 | |
| p_nortriptyline | 730 | maximum | NaN | FALSE | 547.71 | 720 | 0.931 | |
| p_nortriptyline | 730 | minimum | NaN | FALSE | 301.68 | 584 | 0.931 | |
| p_nortriptyline | 730 | latest | NaN | FALSE | 412.1 | 748 | 0.931 | |
| p_nortriptyline | 730 | mean | NaN | FALSE | 414.98 | 1558 | 0.931 | |
| p_olanzapine | 10 | maximum | NaN | FALSE | 184.44 | 248 | 0.995 | |
| p_olanzapine | 10 | latest | NaN | FALSE | 176.04 | 247 | 0.995 | |
| p_olanzapine | 10 | mean | NaN | FALSE | 180.63 | 268 | 0.995 | |
| p_olanzapine | 10 | minimum | NaN | FALSE | 175.67 | 246 | 0.995 | |
| p_olanzapine | 30 | maximum | NaN | FALSE | 176.79 | 258 | 0.989 | X |
| p_olanzapine | 30 | minimum | NaN | FALSE | 162.18 | 258 | 0.989 | X |
| p_olanzapine | 30 | latest | NaN | FALSE | 163.8 | 262 | 0.989 | X |
| p_olanzapine | 30 | mean | NaN | FALSE | 169.86 | 313 | 0.989 | X |
| p_olanzapine | 180 | mean | NaN | FALSE | 163.05 | 411 | 0.97 | |
| p_olanzapine | 180 | minimum | NaN | FALSE | 144.12 | 270 | 0.97 | |
| p_olanzapine | 180 | latest | NaN | FALSE | 152.6 | 288 | 0.97 | |
| p_olanzapine | 180 | maximum | NaN | FALSE | 182.39 | 279 | 0.97 | |
| p_olanzapine | 365 | maximum | NaN | FALSE | 175.63 | 299 | 0.957 | |
| p_olanzapine | 365 | mean | NaN | FALSE | 155.57 | 476 | 0.957 | |
| p_olanzapine | 365 | latest | NaN | FALSE | 145.4 | 299 | 0.957 | |
| p_olanzapine | 365 | minimum | NaN | FALSE | 135.79 | 274 | 0.957 | |
| p_olanzapine | 730 | maximum | NaN | FALSE | 174.8 | 319 | 0.935 | |
| p_olanzapine | 730 | mean | NaN | FALSE | 148.1 | 582 | 0.935 | |
| p_olanzapine | 730 | minimum | NaN | FALSE | 122.51 | 290 | 0.935 | |
| p_olanzapine | 730 | latest | NaN | FALSE | 137.14 | 318 | 0.935 | |
| p_paliperidone | 10 | maximum | NaN | FALSE | 54.255 | 147 | 0.993 | |
| p_paliperidone | 10 | minimum | NaN | FALSE | 53.735 | 146 | 0.993 | |
| p_paliperidone | 10 | mean | NaN | FALSE | 53.995 | 164 | 0.993 | |
| p_paliperidone | 10 | latest | NaN | FALSE | 54.073 | 147 | 0.993 | |
| p_paliperidone | 30 | maximum | NaN | FALSE | 55.391 | 153 | 0.984 | |
| p_paliperidone | 30 | latest | NaN | FALSE | 53.559 | 154 | 0.984 | |
| p_paliperidone | 30 | minimum | NaN | FALSE | 52.304 | 150 | 0.984 | |
| p_paliperidone | 30 | mean | NaN | FALSE | 53.83 | 212 | 0.984 | |
| p_paliperidone | 180 | minimum | NaN | FALSE | 50.74 | 170 | 0.959 | |
| p_paliperidone | 180 | maximum | NaN | FALSE | 61.6 | 177 | 0.959 | |
| p_paliperidone | 180 | mean | NaN | FALSE | 55.971 | 338 | 0.959 | |
| p_paliperidone | 180 | latest | NaN | FALSE | 56.421 | 179 | 0.959 | |
| p_paliperidone | 365 | maximum | NaN | FALSE | 64.562 | 192 | 0.942 | |
| p_paliperidone | 365 | mean | NaN | FALSE | 57.236 | 425 | 0.942 | |
| p_paliperidone | 365 | minimum | NaN | FALSE | 50.377 | 169 | 0.942 | |
| p_paliperidone | 365 | latest | NaN | FALSE | 57.917 | 187 | 0.942 | |
| p_paliperidone | 730 | maximum | NaN | FALSE | 65.989 | 197 | 0.916 | X |
| p_paliperidone | 730 | minimum | NaN | FALSE | 48.115 | 166 | 0.916 | |
| p_paliperidone | 730 | latest | NaN | FALSE | 58.22 | 190 | 0.916 | X |
| p_paliperidone | 730 | mean | NaN | FALSE | 56.747 | 509 | 0.916 | X |
| p_paracetamol | 10 | maximum | NaN | FALSE | 226.37 | 549 | 0.985 | |
| p_paracetamol | 10 | mean | NaN | FALSE | 122.26 | 832 | 0.985 | |
| p_paracetamol | 10 | latest | NaN | FALSE | 40.919 | 257 | 0.985 | |
| p_paracetamol | 10 | minimum | NaN | FALSE | 38.121 | 237 | 0.985 | |
| p_paracetamol | 30 | minimum | NaN | FALSE | 33.454 | 226 | 0.97 | X |
| p_paracetamol | 30 | maximum | NaN | FALSE | 245.55 | 574 | 0.97 | |
| p_paracetamol | 30 | latest | NaN | FALSE | 36.212 | 271 | 0.97 | X |
| p_paracetamol | 30 | mean | NaN | FALSE | 122.5 | 944 | 0.97 | |
| p_paracetamol | 180 | latest | NaN | FALSE | 35.883 | 293 | 0.936 | |
| p_paracetamol | 180 | maximum | NaN | FALSE | 271.88 | 612 | 0.936 | |
| p_paracetamol | 180 | minimum | NaN | FALSE | 31.729 | 212 | 0.936 | |
| p_paracetamol | 180 | mean | NaN | FALSE | 122.78 | 1195 | 0.936 | |
| p_paracetamol | 365 | maximum | NaN | FALSE | 253.91 | 655 | 0.914 | |
| p_paracetamol | 365 | latest | NaN | FALSE | 35.733 | 311 | 0.914 | |
| p_paracetamol | 365 | minimum | NaN | FALSE | 31.726 | 221 | 0.914 | |

| | | | | | | | | | |
|---|---|---|---|---|---|---|---|---|---|
| p_paracetamol | 365 | mean | NaN | FALSE | 111.96 | 1341 | 0.914 | | |
| p_paracetamol | 730 | maximum | NaN | FALSE | 271.09 | 715 | 0.886 | X | |
| p_paracetamol | 730 | minimum | NaN | FALSE | 28.975 | 225 | 0.886 | X | |
| p_paracetamol | 730 | mean | NaN | FALSE | 108.97 | 1606 | 0.886 | | |
| p_paracetamol | 730 | latest | NaN | FALSE | 36.275 | 331 | 0.886 | X | |
| p_risperidone | 10 | minimum | NaN | FALSE | 16.728 | 85 | 0.994 | | |
| p_risperidone | 10 | maximum | NaN | FALSE | 17.019 | 86 | 0.994 | | |
| p_risperidone | 10 | latest | NaN | FALSE | 16.834 | 86 | 0.994 | | |
| p_risperidone | 10 | mean | NaN | FALSE | 16.873 | 98 | 0.994 | | |
| p_risperidone | 30 | maximum | NaN | FALSE | 17.072 | 89 | 0.987 | X | |
| p_risperidone | 30 | latest | NaN | FALSE | 16.047 | 90 | 0.987 | X | |
| p_risperidone | 30 | minimum | NaN | FALSE | 15.642 | 89 | 0.987 | X | |
| p_risperidone | 30 | mean | NaN | FALSE | 16.337 | 123 | 0.987 | X | |
| p_risperidone | 180 | maximum | NaN | FALSE | 19.063 | 99 | 0.968 | | |
| p_risperidone | 180 | latest | NaN | FALSE | 16.641 | 102 | 0.968 | | |
| p_risperidone | 180 | mean | NaN | FALSE | 16.606 | 193 | 0.968 | | |
| p_risperidone | 180 | minimum | NaN | FALSE | 14.513 | 94 | 0.968 | | |
| p_risperidone | 365 | mean | NaN | FALSE | 16.756 | 254 | 0.954 | | |
| p_risperidone | 365 | minimum | NaN | FALSE | 14.121 | 105 | 0.954 | | |
| p_risperidone | 365 | maximum | NaN | FALSE | 19.867 | 108 | 0.954 | | |
| p_risperidone | 365 | latest | NaN | FALSE | 17.143 | 110 | 0.954 | | |
| p_risperidone | 730 | maximum | NaN | FALSE | 20.835 | 117 | 0.934 | | |
| p_risperidone | 730 | mean | NaN | FALSE | 16.715 | 337 | 0.934 | | |
| p_risperidone | 730 | latest | NaN | FALSE | 17.092 | 111 | 0.934 | X | |
| p_risperidone | 730 | minimum | NaN | FALSE | 13.524 | 100 | 0.934 | | |
| paa_grund_af_farlighed | 10 | summed | 0 | FALSE | 2.364 | 81 | 0.99 | | |
| paa_grund_af_farlighed | 10 | boolean | 0 | FALSE | 0.01 | 2 | 0.99 | | X |
| paa_grund_af_farlighed | 10 | count | 0 | FALSE | 0.01 | 4 | 0.99 | | X |
| paa_grund_af_farlighed | 30 | boolean | 0 | FALSE | 0.019 | 2 | 0.981 | X | X |
| paa_grund_af_farlighed | 30 | count | 0 | FALSE | 0.02 | 6 | 0.981 | X | X |
| paa_grund_af_farlighed | 30 | summed | 0 | FALSE | 4.654 | 82 | 0.981 | | |
| paa_grund_af_farlighed | 180 | summed | 0 | FALSE | 14.82 | 97 | 0.958 | | |
| paa_grund_af_farlighed | 180 | boolean | 0 | FALSE | 0.042 | 2 | 0.958 | | X |
| paa_grund_af_farlighed | 180 | count | 0 | FALSE | 0.05 | 10 | 0.958 | | X |
| paa_grund_af_farlighed | 365 | count | 0 | FALSE | 0.075 | 14 | 0.944 | | X |
| paa_grund_af_farlighed | 365 | boolean | 0 | FALSE | 0.056 | 2 | 0.944 | | X |
| paa_grund_af_farlighed | 365 | summed | 0 | FALSE | 24.663 | 116 | 0.944 | | |
| paa_grund_af_farlighed | 730 | count | 0 | FALSE | 0.129 | 20 | 0.916 | X | X |
| paa_grund_af_farlighed | 730 | summed | 0 | FALSE | 50.247 | 154 | 0.916 | X | |
| paa_grund_af_farlighed | 730 | boolean | 0 | FALSE | 0.084 | 2 | 0.916 | X | X |
| paa_grund_af_farlighed | 1 | boolean | 0 | FALSE | 0.023 | 2 | 0.977 | X | X |
| paa_grund_af_farlighed | 3 | boolean | 0 | FALSE | 0.025 | 2 | 0.975 | | X |
| paa_grund_af_farlighed | 7 | summed | 0 | FALSE | 1.902 | 81 | 0.992 | | |
| paa_grund_af_farlighed | 7 | boolean | 0 | FALSE | 0.008 | 2 | 0.992 | X | X |
| paa_grund_af_farlighed | 7 | count | 0 | FALSE | 0.008 | 4 | 0.992 | X | X |
| physical_visits | 365 | boolean | 0 | FALSE | 0.945 | 2 | 0.055 | | X |
| physical_visits | 365 | count | 0 | FALSE | 16.685 | 335 | 0.055 | | X |
| physical_visits | 365 | summed | 0 | FALSE | 16.685 | 335 | 0.055 | | X |
| physical_visits | 10 | count | 0 | FALSE | 0.66 | 23 | 0.64 | | X |
| physical_visits | 10 | boolean | 0 | FALSE | 0.36 | 2 | 0.64 | | X |
| physical_visits | 10 | summed | 0 | FALSE | 0.66 | 23 | 0.64 | | X |
| physical_visits | 730 | count | 0 | FALSE | 29.135 | 561 | 0.033 | X | |
| physical_visits | 730 | summed | 0 | FALSE | 29.135 | 561 | 0.033 | X | |
| physical_visits | 730 | boolean | 0 | FALSE | 0.967 | 2 | 0.033 | X | X |
| physical_visits | 30 | summed | 0 | FALSE | 1.899 | 58 | 0.359 | X | |
| physical_visits | 30 | boolean | 0 | FALSE | 0.641 | 2 | 0.359 | X | |
| physical_visits | 30 | count | 0 | FALSE | 1.899 | 58 | 0.359 | X | |
| physical_visits | 180 | boolean | 0 | FALSE | 0.909 | 2 | 0.091 | | X |
| physical_visits | 180 | count | 0 | FALSE | 9.212 | 189 | 0.091 | | X |
| physical_visits | 180 | summed | 0 | FALSE | 9.212 | 189 | 0.091 | | X |
| physical_visits_to_psychiatry | 365 | count | 0 | FALSE | 13.678 | 322 | 0.012 | | X |
| physical_visits_to_psychiatry | 365 | summed | 0 | FALSE | 96.666 | 57924 | 0.012 | | X |
| physical_visits_to_psychiatry | 365 | boolean | 0 | FALSE | 0.988 | 2 | 0.012 | | |
| physical_visits_to_psychiatry | 10 | boolean | 0 | FALSE | 0.429 | 2 | 0.571 | | X |
| physical_visits_to_psychiatry | 730 | count | 0 | FALSE | 23.348 | 525 | 0.003 | X | |
| physical_visits_to_psychiatry | 730 | boolean | 0 | FALSE | 0.997 | 2 | 0.003 | X | |
| physical_visits_to_psychiatry | 730 | summed | 0 | FALSE | 134.05 | 57764 | 0.003 | X | X |
| physical_visits_to_psychiatry | 10 | summed | 0 | FALSE | 9.905 | 38310 | 0.571 | | X |
| physical_visits_to_psychiatry | 10 | count | 0 | FALSE | 0.8 | 24 | 0.571 | | X |
| physical_visits_to_psychiatry | 30 | summed | 0 | FALSE | 25.21 | 47283 | 0.272 | X | X |
| physical_visits_to_psychiatry | 30 | boolean | 0 | FALSE | 0.728 | 2 | 0.272 | X | X |
| physical_visits_to_psychiatry | 30 | count | 0 | FALSE | 1.973 | 58 | 0.272 | X | |
| physical_visits_to_psychiatry | 180 | summed | 0 | FALSE | 70.287 | 56635 | 0.031 | | |
| physical_visits_to_psychiatry | 180 | count | 0 | FALSE | 7.848 | 181 | 0.031 | | X |
| physical_visits_to_psychiatry | 180 | boolean | 0 | FALSE | 0.969 | 2 | 0.031 | | X |
| physical_visits_to_somatic | 365 | summed | 0 | FALSE | 3.908 | 183 | 0.335 | | |

| | | | | | | | | | |
|---|---|---|---|---|---|---|---|---|---|
| physical_visits_to_somatic | 365 | count | 0 | FALSE | 3.908 | 183 | 0.335 | | |
| physical_visits_to_somatic | 365 | boolean | 0 | FALSE | 0.665 | 2 | 0.335 | | |
| physical_visits_to_somatic | 730 | summed | 0 | FALSE | 6.724 | 267 | 0.236 | X | |
| physical_visits_to_somatic | 730 | count | 0 | FALSE | 6.724 | 267 | 0.236 | X | |
| physical_visits_to_somatic | 730 | boolean | 0 | FALSE | 0.764 | 2 | 0.236 | X | |
| physical_visits_to_somatic | 10 | count | 0 | FALSE | 0.217 | 17 | 0.851 | | X |
| physical_visits_to_somatic | 10 | summed | 0 | FALSE | 0.217 | 17 | 0.851 | | X |
| physical_visits_to_somatic | 10 | boolean | 0 | FALSE | 0.149 | 2 | 0.851 | | X |
| physical_visits_to_somatic | 30 | count | 0 | FALSE | 0.551 | 36 | 0.713 | | |
| physical_visits_to_somatic | 30 | boolean | 0 | FALSE | 0.287 | 2 | 0.713 | X | |
| physical_visits_to_somatic | 30 | summed | 0 | FALSE | 0.551 | 36 | 0.713 | | |
| physical_visits_to_somatic | 180 | boolean | 0 | FALSE | 0.557 | 2 | 0.443 | | |
| physical_visits_to_somatic | 180 | count | 0 | FALSE | 2.227 | 102 | 0.443 | | |
| physical_visits_to_somatic | 180 | summed | 0 | FALSE | 2.227 | 102 | 0.443 | | |
| tfidf_10 | 180 | mean | NaN | FALSE | 0.004 | 478235 | 0.003 | | X |
| tfidf_10 mg | 180 | mean | NaN | FALSE | 0.001 | 147179 | 0.003 | | X |
| tfidf_100 | 180 | mean | NaN | FALSE | 0.001 | 200035 | 0.003 | | X |
| tfidf_100 mg | 180 | mean | NaN | FALSE | 0.001 | 113470 | 0.003 | | X |
| tfidf_1000 | 180 | mean | NaN | FALSE | 0.002 | 220004 | 0.003 | | |
| tfidf_11 | 180 | mean | NaN | FALSE | 0.002 | 313799 | 0.003 | | |
| tfidf_12 | 180 | mean | NaN | FALSE | 0.003 | 383408 | 0.003 | | |
| tfidf_13 | 180 | mean | NaN | FALSE | 0.002 | 298183 | 0.003 | | |
| tfidf_14 | 180 | mean | NaN | FALSE | 0.003 | 417054 | 0.003 | | |
| tfidf_14 dage | 180 | mean | NaN | FALSE | 0.001 | 225365 | 0.003 | | |
| tfidf_15 | 180 | mean | NaN | FALSE | 0.003 | 417140 | 0.003 | | X |
| tfidf_15 mg | 180 | mean | NaN | FALSE | 0.001 | 163433 | 0.003 | | X |
| tfidf_18 | 180 | mean | NaN | FALSE | 0.001 | 229258 | 0.003 | | |
| tfidf_20 | 180 | mean | NaN | FALSE | 0.003 | 382599 | 0.003 | | |
| tfidf_20 mg | 180 | mean | NaN | FALSE | 0 | 62251 | 0.003 | | |
| tfidf_200 | 180 | mean | NaN | FALSE | 0 | 108987 | 0.003 | | |
| tfidf_23 | 180 | mean | NaN | FALSE | 0.001 | 273502 | 0.003 | | |
| tfidf_25 | 180 | mean | NaN | FALSE | 0.001 | 251882 | 0.003 | | |
| tfidf_25 mg | 180 | mean | NaN | FALSE | 0.001 | 149426 | 0.003 | | |
| tfidf_30 | 180 | mean | NaN | FALSE | 0.002 | 297204 | 0.003 | | |
| tfidf_40 | 180 | mean | NaN | FALSE | 0.001 | 123187 | 0.003 | | |
| tfidf_50 | 180 | mean | NaN | FALSE | 0.001 | 216062 | 0.003 | | |
| tfidf_50 mg | 180 | mean | NaN | FALSE | 0.001 | 153421 | 0.003 | | |
| tfidf_75 | 180 | mean | NaN | FALSE | 0.001 | 133294 | 0.003 | | |
| tfidf_75 mg | 180 | mean | NaN | FALSE | 0.001 | 108127 | 0.003 | | |
| tfidf_abilify | 180 | mean | NaN | FALSE | 0.001 | 115601 | 0.003 | | |
| tfidf_accepterer | 180 | mean | NaN | FALSE | 0.003 | 382631 | 0.003 | | X |
| tfidf_adfærd | 180 | mean | NaN | FALSE | 0.005 | 408741 | 0.003 | | X |
| tfidf_adhd | 180 | mean | NaN | FALSE | 0.001 | 79744 | 0.003 | | |
| tfidf_adspurgt | 180 | mean | NaN | FALSE | 0.008 | 528830 | 0.003 | | X |
| tfidf_afd | 180 | mean | NaN | FALSE | 0.012 | 544708 | 0.003 | | X |
| tfidf_afdelingen | 180 | mean | NaN | FALSE | 0.012 | 556496 | 0.003 | | |
| tfidf_afsluttes | 180 | mean | NaN | FALSE | 0.001 | 126445 | 0.003 | | |
| tfidf_afstand | 180 | mean | NaN | FALSE | 0.002 | 330144 | 0.003 | | |
| tfidf_afsted | 180 | mean | NaN | FALSE | 0.002 | 282592 | 0.003 | | X |
| tfidf_aftale | 180 | mean | NaN | FALSE | 0.008 | 533210 | 0.003 | | |
| tfidf_aftaler | 180 | mean | NaN | FALSE | 0.007 | 508543 | 0.003 | | |
| tfidf_aftales | 180 | mean | NaN | FALSE | 0.004 | 429497 | 0.003 | | |
| tfidf_aftalt | 180 | mean | NaN | FALSE | 0.009 | 517092 | 0.003 | | X |
| tfidf_aften | 180 | mean | NaN | FALSE | 0.015 | 566398 | 0.003 | | X |
| tfidf_aftenen | 180 | mean | NaN | FALSE | 0.007 | 492490 | 0.003 | | X |
| tfidf_aftensmad | 180 | mean | NaN | FALSE | 0.005 | 418076 | 0.003 | | X |
| tfidf_afventer | 180 | mean | NaN | FALSE | 0.002 | 243874 | 0.003 | | |
| tfidf_afvisende | 180 | mean | NaN | FALSE | 0.003 | 305526 | 0.003 | | X |
| tfidf_afviser | 180 | mean | NaN | FALSE | 0.005 | 448246 | 0.003 | | X |
| tfidf_aktivitet | 180 | mean | NaN | FALSE | 0.001 | 233345 | 0.003 | | X |
| tfidf_aktiviteter | 180 | mean | NaN | FALSE | 0.003 | 348188 | 0.003 | | X |
| tfidf_aktuelle | 180 | mean | NaN | FALSE | 0.002 | 380336 | 0.003 | | |
| tfidf_aktuelt | 180 | mean | NaN | FALSE | 0.005 | 514243 | 0.003 | | |
| tfidf_akut | 180 | mean | NaN | FALSE | 0.003 | 334410 | 0.003 | | X |
| tfidf_alkohol | 180 | mean | NaN | FALSE | 0.002 | 220670 | 0.003 | | |
| tfidf_amb | 180 | mean | NaN | FALSE | 0.002 | 196085 | 0.003 | | |
| tfidf_ambulant | 180 | mean | NaN | FALSE | 0.002 | 264823 | 0.003 | | |
| tfidf_ang | 180 | mean | NaN | FALSE | 0.003 | 355211 | 0.003 | | |
| tfidf_angiveligt | 180 | mean | NaN | FALSE | 0.003 | 380581 | 0.003 | | X |
| tfidf_angiver | 180 | mean | NaN | FALSE | 0.016 | 589035 | 0.003 | | |
| tfidf_angst | 180 | mean | NaN | FALSE | 0.007 | 413078 | 0.003 | | X |
| tfidf_angsten | 180 | mean | NaN | FALSE | 0.001 | 145482 | 0.003 | | X |
| tfidf_fornavn* | 180 | mean | NaN | FALSE | 0.002 | 128641 | 0.003 | | |
| tfidf_appetit | 180 | mean | NaN | FALSE | 0.002 | 285537 | 0.003 | | X |
| tfidf_arbejde | 180 | mean | NaN | FALSE | 0.004 | 410077 | 0.003 | | |
| tfidf_arbejder | 180 | mean | NaN | FALSE | 0.002 | 244405 | 0.003 | | |

| | | | | | | | | | |
|---|---|---|---|---|---|---|---|---|---|
| tfidf_arbejdet | 180 | mean | NaN | FALSE | 0.001 | 216730 | 0.003 | | |
| tfidf_bad | 180 | mean | NaN | FALSE | 0.007 | 447699 | 0.003 | | |
| tfidf_baggrund | 180 | mean | NaN | FALSE | 0.001 | 293703 | 0.003 | | X |
| tfidf_bange | 180 | mean | NaN | FALSE | 0.006 | 489784 | 0.003 | | |
| tfidf_barn | 180 | mean | NaN | FALSE | 0.001 | 176883 | 0.003 | | |
| tfidf_beder | 180 | mean | NaN | FALSE | 0.004 | 430726 | 0.003 | | X |
| tfidf_bedre | 180 | mean | NaN | FALSE | 0.01 | 561238 | 0.003 | | X |
| tfidf_bedring | 180 | mean | NaN | FALSE | 0.004 | 373327 | 0.003 | | X |
| tfidf_bedst | 180 | mean | NaN | FALSE | 0.001 | 300833 | 0.003 | | |
| tfidf_bedt | 180 | mean | NaN | FALSE | 0.002 | 314200 | 0.003 | | X |
| tfidf_begynder | 180 | mean | NaN | FALSE | 0.002 | 354883 | 0.003 | | X |
| tfidf_begyndt | 180 | mean | NaN | FALSE | 0.002 | 317472 | 0.003 | | |
| tfidf_beh | 180 | mean | NaN | FALSE | 0.002 | 239917 | 0.003 | | |
| tfidf_behandler | 180 | mean | NaN | FALSE | 0.002 | 271785 | 0.003 | | |
| tfidf_behandling | 180 | mean | NaN | FALSE | 0.008 | 539922 | 0.003 | | |
| tfidf_behandlingen | 180 | mean | NaN | FALSE | 0.003 | 290327 | 0.003 | | X |
| tfidf_behov | 180 | mean | NaN | FALSE | 0.007 | 532844 | 0.003 | | |
| tfidf_bekræfter | 180 | mean | NaN | FALSE | 0.001 | 293848 | 0.003 | | X |
| tfidf_bekymret | 180 | mean | NaN | FALSE | 0.004 | 438693 | 0.003 | | X |
| tfidf_bekymring | 180 | mean | NaN | FALSE | 0.001 | 280102 | 0.003 | | |
| tfidf_benægter | 180 | mean | NaN | FALSE | 0.003 | 350514 | 0.003 | | X |
| tfidf_besked | 180 | mean | NaN | FALSE | 0.002 | 337093 | 0.003 | | |
| tfidf_beskrevet | 180 | mean | NaN | FALSE | 0.002 | 329740 | 0.003 | | X |
| tfidf_beskriver | 180 | mean | NaN | FALSE | 0.008 | 531077 | 0.003 | | X |
| tfidf_beskrives | 180 | mean | NaN | FALSE | 0.001 | 196146 | 0.003 | | X |
| tfidf_bestilt | 180 | mean | NaN | FALSE | 0.002 | 266684 | 0.003 | | |
| tfidf_besøg | 180 | mean | NaN | FALSE | 0.018 | 560708 | 0.003 | | X |
| tfidf_bivirkninger | 180 | mean | NaN | FALSE | 0.002 | 282162 | 0.003 | | |
| tfidf_bla | 180 | mean | NaN | FALSE | 0.003 | 430629 | 0.003 | | |
| tfidf_blevet | 180 | mean | NaN | FALSE | 0.009 | 577756 | 0.003 | | X |
| tfidf_blodprøver | 180 | mean | NaN | FALSE | 0.002 | 257435 | 0.003 | | X |
| tfidf_bo | 180 | mean | NaN | FALSE | 0.002 | 253568 | 0.003 | | |
| tfidf_bor | 180 | mean | NaN | FALSE | 0.009 | 496269 | 0.003 | | X |
| tfidf_bosted | 180 | mean | NaN | FALSE | 0.003 | 172926 | 0.003 | | |
| tfidf_bostedet | 180 | mean | NaN | FALSE | 0.002 | 105417 | 0.003 | | X |
| tfidf_bostøtte | 180 | mean | NaN | FALSE | 0.005 | 256495 | 0.003 | | |
| tfidf_bruge | 180 | mean | NaN | FALSE | 0.003 | 398957 | 0.003 | | |
| tfidf_bruger | 180 | mean | NaN | FALSE | 0.002 | 384873 | 0.003 | | |
| tfidf_brugt | 180 | mean | NaN | FALSE | 0.002 | 341768 | 0.003 | | |
| tfidf_børn | 180 | mean | NaN | FALSE | 0.004 | 283021 | 0.003 | | |
| tfidf_børnene | 180 | mean | NaN | FALSE | 0.001 | 102934 | 0.003 | | |
| tfidf_concerta | 180 | mean | NaN | FALSE | 0 | 11078 | 0.003 | | |
| tfidf_dage | 180 | mean | NaN | FALSE | 0.007 | 558204 | 0.003 | | |
| tfidf_dagen | 180 | mean | NaN | FALSE | 0.007 | 538809 | 0.003 | | X |
| tfidf_dagens | 180 | mean | NaN | FALSE | 0.002 | 308428 | 0.003 | | |
| tfidf_dagens samtale | 180 | mean | NaN | FALSE | 0.001 | 170503 | 0.003 | | |
| tfidf_dagligt | 180 | mean | NaN | FALSE | 0.002 | 375160 | 0.003 | | |
| tfidf_datter | 180 | mean | NaN | FALSE | 0.005 | 178756 | 0.003 | | |
| tfidf_datteren | 180 | mean | NaN | FALSE | 0.002 | 105569 | 0.003 | | |
| tfidf_dd | 180 | mean | NaN | FALSE | 0.006 | 486081 | 0.003 | | |
| tfidf_dels | 180 | mean | NaN | FALSE | 0.001 | 205299 | 0.003 | | |
| tfidf_deltage | 180 | mean | NaN | FALSE | 0.003 | 389578 | 0.003 | | X |
| tfidf_deltagelse | 180 | mean | NaN | FALSE | 0.002 | 231087 | 0.003 | | |
| tfidf_deltager | 180 | mean | NaN | FALSE | 0.004 | 398072 | 0.003 | | X |
| tfidf_depot | 180 | mean | NaN | FALSE | 0.001 | 104567 | 0.003 | | |
| tfidf_depression | 180 | mean | NaN | FALSE | 0.004 | 299014 | 0.003 | | X |
| tfidf_depressiv | 180 | mean | NaN | FALSE | 0.003 | 295352 | 0.003 | | X |
| tfidf_depressive | 180 | mean | NaN | FALSE | 0.003 | 326491 | 0.003 | | X |
| tfidf_depressive symptomer | 180 | mean | NaN | FALSE | 0.002 | 245598 | 0.003 | | X |
| tfidf_derhjemme | 180 | mean | NaN | FALSE | 0.003 | 370506 | 0.003 | | X |
| tfidf_derudover | 180 | mean | NaN | FALSE | 0.002 | 333040 | 0.003 | | |
| tfidf_desuden | 180 | mean | NaN | FALSE | 0.004 | 463274 | 0.003 | | |
| tfidf_dgl | 180 | mean | NaN | FALSE | 0.001 | 192455 | 0.003 | | |
| tfidf_dosis | 180 | mean | NaN | FALSE | 0.001 | 263551 | 0.003 | | |
| tfidf_dreng | 180 | mean | NaN | FALSE | 0 | 43883 | 0.003 | | |
| tfidf_drikke | 180 | mean | NaN | FALSE | 0.003 | 319463 | 0.003 | | X |
| tfidf_drøfter | 180 | mean | NaN | FALSE | 0.001 | 169212 | 0.003 | | |
| tfidf_drøftes | 180 | mean | NaN | FALSE | 0.001 | 175102 | 0.003 | | |
| tfidf_dårlig | 180 | mean | NaN | FALSE | 0.005 | 494991 | 0.003 | | |
| tfidf_dårligt | 180 | mean | NaN | FALSE | 0.006 | 499914 | 0.003 | | |
| tfidf_døren | 180 | mean | NaN | FALSE | 0.003 | 334230 | 0.003 | | X |
| tfidf_ect | 180 | mean | NaN | FALSE | 0.009 | 174369 | 0.003 | | X |
| tfidf_effekt | 180 | mean | NaN | FALSE | 0.005 | 488337 | 0.003 | | |
| tfidf_efterfølgende | 180 | mean | NaN | FALSE | 0.008 | 542519 | 0.003 | | |
| tfidf_eftermiddag | 180 | mean | NaN | FALSE | 0.006 | 448058 | 0.003 | | X |
| tfidf_egentlig | 180 | mean | NaN | FALSE | 0.002 | 357304 | 0.003 | | |

| | | | | | | | | | |
|---|---|---|---|---|---|---|---|---|---|
| tfidf_eget | 180 | mean | NaN | FALSE | 0.004 | 468978 | 0.003 | | |
| tfidf_egne | 180 | mean | NaN | FALSE | 0.002 | 369936 | 0.003 | | X |
| tfidf_ekg | 180 | mean | NaN | FALSE | 0.002 | 210580 | 0.003 | | X |
| tfidf_ekstra | 180 | mean | NaN | FALSE | 0.001 | 251507 | 0.003 | | |
| tfidf_el | 180 | mean | NaN | FALSE | 0.003 | 283057 | 0.003 | | |
| tfidf_emotionel | 180 | mean | NaN | FALSE | 0.007 | 520576 | 0.003 | | |
| tfidf_emotionel kontakt | 180 | mean | NaN | FALSE | 0.006 | 491009 | 0.003 | | |
| tfidf_endvidere | 180 | mean | NaN | FALSE | 0.002 | 306682 | 0.003 | | |
| tfidf_energi | 180 | mean | NaN | FALSE | 0.003 | 372052 | 0.003 | | X |
| tfidf_enkelt | 180 | mean | NaN | FALSE | 0.002 | 308983 | 0.003 | | |
| tfidf_enkelte | 180 | mean | NaN | FALSE | 0.001 | 268002 | 0.003 | | |
| tfidf_evt | 180 | mean | NaN | FALSE | 0.005 | 497225 | 0.003 | | |
| tfidf_falde | 180 | mean | NaN | FALSE | 0.002 | 355312 | 0.003 | | |
| tfidf_falde søvn | 180 | mean | NaN | FALSE | 0.001 | 248969 | 0.003 | | |
| tfidf_falder | 180 | mean | NaN | FALSE | 0.003 | 417760 | 0.003 | | X |
| tfidf_familie | 180 | mean | NaN | FALSE | 0.004 | 400083 | 0.003 | | |
| tfidf_familien | 180 | mean | NaN | FALSE | 0.003 | 335357 | 0.003 | | |
| tfidf_fast | 180 | mean | NaN | FALSE | 0.003 | 423184 | 0.003 | | X |
| tfidf_faste | 180 | mean | NaN | FALSE | 0.002 | 278663 | 0.003 | | X |
| tfidf_feks | 180 | mean | NaN | FALSE | 0.001 | 292655 | 0.003 | | |
| tfidf_ferie | 180 | mean | NaN | FALSE | 0.001 | 199674 | 0.003 | | |
| tfidf_film | 180 | mean | NaN | FALSE | 0.002 | 240777 | 0.003 | | X |
| tfidf_fin | 180 | mean | NaN | FALSE | 0.003 | 319500 | 0.003 | | X |
| tfidf_finder | 180 | mean | NaN | FALSE | 0.003 | 405878 | 0.003 | | X |
| tfidf_findes | 180 | mean | NaN | FALSE | 0.002 | 288939 | 0.003 | | X |
| tfidf_fint | 180 | mean | NaN | FALSE | 0.005 | 468554 | 0.003 | | |
| tfidf_flytte | 180 | mean | NaN | FALSE | 0.002 | 304610 | 0.003 | | |
| tfidf_flyttet | 180 | mean | NaN | FALSE | 0.002 | 271911 | 0.003 | | |
| tfidf_fokus | 180 | mean | NaN | FALSE | 0.002 | 302201 | 0.003 | | |
| tfidf_folk | 180 | mean | NaN | FALSE | 0.001 | 268123 | 0.003 | | X |
| tfidf_forbindelse | 180 | mean | NaN | FALSE | 0.004 | 477214 | 0.003 | | |
| tfidf_forhold | 180 | mean | NaN | FALSE | 0.005 | 506842 | 0.003 | | |
| tfidf_forklare | 180 | mean | NaN | FALSE | 0.002 | 335507 | 0.003 | | X |
| tfidf_forklarer | 180 | mean | NaN | FALSE | 0.002 | 359194 | 0.003 | | X |
| tfidf_forløb | 180 | mean | NaN | FALSE | 0.003 | 365050 | 0.003 | | |
| tfidf_forløbet | 180 | mean | NaN | FALSE | 0.001 | 231894 | 0.003 | | |
| tfidf_form | 180 | mean | NaN | FALSE | 0.003 | 453852 | 0.003 | | |
| tfidf_formel | 180 | mean | NaN | FALSE | 0.008 | 538118 | 0.003 | | |
| tfidf_formel emotionel | 180 | mean | NaN | FALSE | 0.004 | 414142 | 0.003 | | |
| tfidf_formel kontakt | 180 | mean | NaN | FALSE | 0.003 | 367833 | 0.003 | | |
| tfidf_formentlig | 180 | mean | NaN | FALSE | 0.001 | 265638 | 0.003 | | X |
| tfidf_formiddag | 180 | mean | NaN | FALSE | 0.004 | 410565 | 0.003 | | X |
| tfidf_forpint | 180 | mean | NaN | FALSE | 0.008 | 443228 | 0.003 | | X |
| tfidf_forsat | 180 | mean | NaN | FALSE | 0.003 | 295080 | 0.003 | | |
| tfidf_forskellige | 180 | mean | NaN | FALSE | 0.003 | 422636 | 0.003 | | X |
| tfidf_forstå | 180 | mean | NaN | FALSE | 0.002 | 355015 | 0.003 | | X |
| tfidf_forstår | 180 | mean | NaN | FALSE | 0.002 | 295043 | 0.003 | | X |
| tfidf_forsænket | 180 | mean | NaN | FALSE | 0.004 | 312421 | 0.003 | | X |
| tfidf_forsøge | 180 | mean | NaN | FALSE | 0.003 | 394600 | 0.003 | | |
| tfidf_forsøger | 180 | mean | NaN | FALSE | 0.003 | 445838 | 0.003 | | X |
| tfidf_forsøgt | 180 | mean | NaN | FALSE | 0.005 | 485349 | 0.003 | | X |
| tfidf_fortalt | 180 | mean | NaN | FALSE | 0.003 | 367946 | 0.003 | | |
| tfidf_fortsætte | 180 | mean | NaN | FALSE | 0.001 | 266268 | 0.003 | | |
| tfidf_fortsætter | 180 | mean | NaN | FALSE | 0.001 | 231188 | 0.003 | | X |
| tfidf_fortælle | 180 | mean | NaN | FALSE | 0.003 | 442822 | 0.003 | | X |
| tfidf_fortæller | 180 | mean | NaN | FALSE | 0.032 | 632898 | 0.003 | | X |
| tfidf_fortæller pt | 180 | mean | NaN | FALSE | 0.003 | 410282 | 0.003 | | |
| tfidf_forværring | 180 | mean | NaN | FALSE | 0.003 | 370545 | 0.003 | | |
| tfidf_forældre | 180 | mean | NaN | FALSE | 0.004 | 254713 | 0.003 | | |
| tfidf_forældrene | 180 | mean | NaN | FALSE | 0.002 | 156073 | 0.003 | | |
| tfidf_fraset | 180 | mean | NaN | FALSE | 0.003 | 324901 | 0.003 | | |
| tfidf_fredag | 180 | mean | NaN | FALSE | 0.004 | 371708 | 0.003 | | X |
| tfidf_frem | 180 | mean | NaN | FALSE | 0.005 | 508753 | 0.003 | | |
| tfidf_fremstår | 180 | mean | NaN | FALSE | 0.022 | 603490 | 0.003 | | X |
| tfidf_fremtiden | 180 | mean | NaN | FALSE | 0.002 | 291538 | 0.003 | | X |
| tfidf_fremtræder | 180 | mean | NaN | FALSE | 0.004 | 352161 | 0.003 | | |
| tfidf_frokost | 180 | mean | NaN | FALSE | 0.006 | 405004 | 0.003 | | X |
| tfidf_fulgt | 180 | mean | NaN | FALSE | 0.003 | 326148 | 0.003 | | |
| tfidf_fundet | 180 | mean | NaN | FALSE | 0.002 | 329854 | 0.003 | | X |
| tfidf_fylder | 180 | mean | NaN | FALSE | 0.002 | 315078 | 0.003 | | X |
| tfidf_fysisk | 180 | mean | NaN | FALSE | 0.002 | 325501 | 0.003 | | X |
| tfidf_fået | 180 | mean | NaN | FALSE | 0.017 | 613789 | 0.003 | | |
| tfidf_fælles | 180 | mean | NaN | FALSE | 0.004 | 359216 | 0.003 | | X |
| tfidf_fællesmiljøet | 180 | mean | NaN | FALSE | 0.007 | 335338 | 0.003 | | X |
| tfidf_føle | 180 | mean | NaN | FALSE | 0.002 | 304361 | 0.003 | | X |
| tfidf_følelser | 180 | mean | NaN | FALSE | 0.001 | 233456 | 0.003 | | X |

| | | | | | | | | | |
|---|---|---|---|---|---|---|---|---|---|
| tfidf_føler | 180 | mean | NaN | FALSE | 0.014 | 588604 | 0.003 | | X |
| tfidf_følges | 180 | mean | NaN | FALSE | 0.002 | 318264 | 0.003 | | X |
| tfidf_følt | 180 | mean | NaN | FALSE | 0.002 | 311946 | 0.003 | | |
| tfidf_følte | 180 | mean | NaN | FALSE | 0.002 | 293024 | 0.003 | | X |
| tfidf_gange | 180 | mean | NaN | FALSE | 0.012 | 592748 | 0.003 | | X |
| tfidf_gangen | 180 | mean | NaN | FALSE | 0.006 | 430233 | 0.003 | | X |
| tfidf_generelt | 180 | mean | NaN | FALSE | 0.002 | 340156 | 0.003 | | |
| tfidf_gik | 180 | mean | NaN | FALSE | 0.005 | 477490 | 0.003 | | X |
| tfidf_give | 180 | mean | NaN | FALSE | 0.004 | 468530 | 0.003 | | X |
| tfidf_gives | 180 | mean | NaN | FALSE | 0.002 | 316971 | 0.003 | | X |
| tfidf_glad | 180 | mean | NaN | FALSE | 0.01 | 555048 | 0.003 | | X |
| tfidf_glæde | 180 | mean | NaN | FALSE | 0.001 | 268621 | 0.003 | | X |
| tfidf_glæder | 180 | mean | NaN | FALSE | 0.002 | 292716 | 0.003 | | X |
| tfidf_gode | 180 | mean | NaN | FALSE | 0.002 | 359086 | 0.003 | | X |
| tfidf_grad | 180 | mean | NaN | FALSE | 0.002 | 323198 | 0.003 | | |
| tfidf_grund | 180 | mean | NaN | FALSE | 0.002 | 309761 | 0.003 | | X |
| tfidf_grundet | 180 | mean | NaN | FALSE | 0.009 | 555254 | 0.003 | | X |
| tfidf_grådlabil | 180 | mean | NaN | FALSE | 0.005 | 311665 | 0.003 | | X |
| tfidf_grædende | 180 | mean | NaN | FALSE | 0.004 | 262500 | 0.003 | | |
| tfidf_gået | 180 | mean | NaN | FALSE | 0.009 | 542656 | 0.003 | | X |
| tfidf_gåtur | 180 | mean | NaN | FALSE | 0.005 | 339103 | 0.003 | | X |
| tfidf_hallucinationer | 180 | mean | NaN | FALSE | 0.002 | 319961 | 0.003 | | X |
| tfidf_handle | 180 | mean | NaN | FALSE | 0.002 | 315988 | 0.003 | | X |
| tfidf_hash | 180 | mean | NaN | FALSE | 0.002 | 139207 | 0.003 | | X |
| tfidf_haven | 180 | mean | NaN | FALSE | 0.004 | 277693 | 0.003 | | X |
| tfidf_henblik | 180 | mean | NaN | FALSE | 0.001 | 231066 | 0.003 | | |
| tfidf_hente | 180 | mean | NaN | FALSE | 0.003 | 374304 | 0.003 | | |
| tfidf_henvender | 180 | mean | NaN | FALSE | 0.006 | 456441 | 0.003 | | |
| tfidf_henvises | 180 | mean | NaN | FALSE | 0.002 | 265839 | 0.003 | | |
| tfidf_henvisning | 180 | mean | NaN | FALSE | 0.001 | 216558 | 0.003 | | |
| tfidf_henvist | 180 | mean | NaN | FALSE | 0.003 | 304744 | 0.003 | | |
| tfidf_herfra | 180 | mean | NaN | FALSE | 0.002 | 296166 | 0.003 | | |
| tfidf_hertil | 180 | mean | NaN | FALSE | 0.002 | 347628 | 0.003 | | X |
| tfidf_herunder | 180 | mean | NaN | FALSE | 0.001 | 260303 | 0.003 | | |
| tfidf_hjem | 180 | mean | NaN | FALSE | 0.011 | 538627 | 0.003 | | X |
| tfidf_hjemme | 180 | mean | NaN | FALSE | 0.005 | 447672 | 0.003 | | X |
| tfidf_hjemmebesøg | 180 | mean | NaN | FALSE | 0.004 | 271613 | 0.003 | | X |
| tfidf_hjemmet | 180 | mean | NaN | FALSE | 0.002 | 289461 | 0.003 | | |
| tfidf_hjulpet | 180 | mean | NaN | FALSE | 0.002 | 334152 | 0.003 | | X |
| tfidf_hjælp | 180 | mean | NaN | FALSE | 0.007 | 540375 | 0.003 | | |
| tfidf_hjælpe | 180 | mean | NaN | FALSE | 0.005 | 491590 | 0.003 | | |
| tfidf_hjælper | 180 | mean | NaN | FALSE | 0.003 | 429538 | 0.003 | | X |
| tfidf_holde | 180 | mean | NaN | FALSE | 0.005 | 514081 | 0.003 | | |
| tfidf_holdt | 180 | mean | NaN | FALSE | 0.002 | 317799 | 0.003 | | |
| tfidf_hovedet | 180 | mean | NaN | FALSE | 0.004 | 443117 | 0.003 | | X |
| tfidf_hovedpine | 180 | mean | NaN | FALSE | 0.003 | 250826 | 0.003 | | |
| tfidf_humør | 180 | mean | NaN | FALSE | 0.006 | 429084 | 0.003 | | X |
| tfidf_humøret | 180 | mean | NaN | FALSE | 0.002 | 290055 | 0.003 | | |
| tfidf_hurtigt | 180 | mean | NaN | FALSE | 0.004 | 455454 | 0.003 | | X |
| tfidf_huske | 180 | mean | NaN | FALSE | 0.003 | 380083 | 0.003 | | |
| tfidf_husker | 180 | mean | NaN | FALSE | 0.001 | 222445 | 0.003 | | |
| tfidf_hverdagen | 180 | mean | NaN | FALSE | 0.001 | 184309 | 0.003 | | X |
| tfidf_hverken | 180 | mean | NaN | FALSE | 0.002 | 310426 | 0.003 | | X |
| tfidf_hvilket | 180 | mean | NaN | FALSE | 0.012 | 596280 | 0.003 | | |
| tfidf_hvilket pt | 180 | mean | NaN | FALSE | 0.002 | 345546 | 0.003 | | |
| tfidf_hvorvidt | 180 | mean | NaN | FALSE | 0.001 | 277311 | 0.003 | | |
| tfidf_højt | 180 | mean | NaN | FALSE | 0.002 | 293631 | 0.003 | | X |
| tfidf_høre | 180 | mean | NaN | FALSE | 0.004 | 459557 | 0.003 | | X |
| tfidf_idag | 180 | mean | NaN | FALSE | 0.006 | 472576 | 0.003 | | X |
| tfidf_idet | 180 | mean | NaN | FALSE | 0.004 | 443092 | 0.003 | | X |
| tfidf_ifht | 180 | mean | NaN | FALSE | 0.001 | 199072 | 0.003 | | |
| tfidf_ifm | 180 | mean | NaN | FALSE | 0.002 | 293518 | 0.003 | | |
| tfidf_ift | 180 | mean | NaN | FALSE | 0.005 | 465764 | 0.003 | | |
| tfidf_ifølge | 180 | mean | NaN | FALSE | 0.001 | 278469 | 0.003 | | X |
| tfidf_igang | 180 | mean | NaN | FALSE | 0.002 | 283342 | 0.003 | | |
| tfidf_igår | 180 | mean | NaN | FALSE | 0.004 | 413568 | 0.003 | | |
| tfidf_imorgen | 180 | mean | NaN | FALSE | 0.005 | 421907 | 0.003 | | |
| tfidf_imødekommende | 180 | mean | NaN | FALSE | 0.006 | 437779 | 0.003 | | X |
| tfidf_inde | 180 | mean | NaN | FALSE | 0.004 | 432889 | 0.003 | | X |
| tfidf_inden | 180 | mean | NaN | FALSE | 0.007 | 529565 | 0.003 | | X |
| tfidf_indforst√•et | 180 | mean | NaN | FALSE | 0.001 | 247981 | 0.003 | | |
| tfidf_indgå | 180 | mean | NaN | FALSE | 0.002 | 293066 | 0.003 | | |
| tfidf_indimellem | 180 | mean | NaN | FALSE | 0.002 | 330929 | 0.003 | | X |
| tfidf_indlagt | 180 | mean | NaN | FALSE | 0.01 | 570313 | 0.003 | | X |
| tfidf_indlæggelse | 180 | mean | NaN | FALSE | 0.01 | 555547 | 0.003 | | |
| tfidf_indlæggelsen | 180 | mean | NaN | FALSE | 0.005 | 485234 | 0.003 | | |

| | | | | | | | | | |
|---|---|---|---|---|---|---|---|---|---|
| tfidf_indlægges | 180 | mean | NaN | FALSE | 0.005 | 417955 | 0.003 | | X |
| tfidf_indre | 180 | mean | NaN | FALSE | 0.002 | 261019 | 0.003 | | |
| tfidf_indstillet | 180 | mean | NaN | FALSE | 0.001 | 248160 | 0.003 | | |
| tfidf_indtryk | 180 | mean | NaN | FALSE | 0.001 | 243767 | 0.003 | | |
| tfidf_informeres | 180 | mean | NaN | FALSE | 0.002 | 356806 | 0.003 | | X |
| tfidf_informeret | 180 | mean | NaN | FALSE | 0.005 | 457129 | 0.003 | | |
| tfidf_initiativ | 180 | mean | NaN | FALSE | 0.002 | 311699 | 0.003 | | X |
| tfidf_interesse | 180 | mean | NaN | FALSE | 0.001 | 225957 | 0.003 | | |
| tfidf_interesseret | 180 | mean | NaN | FALSE | 0.002 | 312859 | 0.003 | | |
| tfidf_irritabel | 180 | mean | NaN | FALSE | 0.003 | 261501 | 0.003 | | X |
| tfidf_ked | 180 | mean | NaN | FALSE | 0.005 | 448606 | 0.003 | | X |
| tfidf_kender | 180 | mean | NaN | FALSE | 0.002 | 368642 | 0.003 | | |
| tfidf_kendt | 180 | mean | NaN | FALSE | 0.008 | 505432 | 0.003 | | X |
| tfidf_kg | 180 | mean | NaN | FALSE | 0.002 | 198300 | 0.003 | | |
| tfidf_kigger | 180 | mean | NaN | FALSE | 0.003 | 374316 | 0.003 | | X |
| tfidf_kl | 180 | mean | NaN | FALSE | 0.027 | 613849 | 0.003 | | |
| tfidf_klager | 180 | mean | NaN | FALSE | 0.01 | 444184 | 0.003 | | X |
| tfidf_klar | 180 | mean | NaN | FALSE | 0.009 | 569911 | 0.003 | | |
| tfidf_klar orienteret | 180 | mean | NaN | FALSE | 0.003 | 341069 | 0.003 | | |
| tfidf_klare | 180 | mean | NaN | FALSE | 0.002 | 361300 | 0.003 | | |
| tfidf_klart | 180 | mean | NaN | FALSE | 0.002 | 307766 | 0.003 | | X |
| tfidf_klasse | 180 | mean | NaN | FALSE | 0 | 56291 | 0.003 | | |
| tfidf_klinik | 180 | mean | NaN | FALSE | 0.002 | 193622 | 0.003 | | |
| tfidf_kollega | 180 | mean | NaN | FALSE | 0.003 | 326326 | 0.003 | | X |
| tfidf_kommende | 180 | mean | NaN | FALSE | 0.001 | 221560 | 0.003 | | X |
| tfidf_kommune | 180 | mean | NaN | FALSE | 0.001 | 159506 | 0.003 | | |
| tfidf_kommunen | 180 | mean | NaN | FALSE | 0.002 | 237141 | 0.003 | | |
| tfidf_koncentrere | 180 | mean | NaN | FALSE | 0.001 | 221391 | 0.003 | | |
| tfidf_konf | 180 | mean | NaN | FALSE | 0.001 | 176717 | 0.003 | | |
| tfidf_konstant | 180 | mean | NaN | FALSE | 0.002 | 313794 | 0.003 | | X |
| tfidf_kontakt | 180 | mean | NaN | FALSE | 0.021 | 621754 | 0.003 | | |
| tfidf_kontakt pt | 180 | mean | NaN | FALSE | 0.002 | 296422 | 0.003 | | |
| tfidf_kontakte | 180 | mean | NaN | FALSE | 0.003 | 399100 | 0.003 | | |
| tfidf_kontakten | 180 | mean | NaN | FALSE | 0.021 | 592490 | 0.003 | | X |
| tfidf_kontakter | 180 | mean | NaN | FALSE | 0.003 | 369074 | 0.003 | | |
| tfidf_kontaktes | 180 | mean | NaN | FALSE | 0.002 | 274579 | 0.003 | | X |
| tfidf_kontaktet | 180 | mean | NaN | FALSE | 0.003 | 348433 | 0.003 | | X |
| tfidf_kontaktperson | 180 | mean | NaN | FALSE | 0.004 | 368164 | 0.003 | | |
| tfidf_kontrol | 180 | mean | NaN | FALSE | 0.001 | 213612 | 0.003 | | |
| tfidf_kort | 180 | mean | NaN | FALSE | 0.008 | 545781 | 0.003 | | X |
| tfidf_kp | 180 | mean | NaN | FALSE | 0.006 | 346565 | 0.003 | | |
| tfidf_kroppen | 180 | mean | NaN | FALSE | 0.003 | 363831 | 0.003 | | |
| tfidf_kunnet | 180 | mean | NaN | FALSE | 0.002 | 338601 | 0.003 | | |
| tfidf_kvalme | 180 | mean | NaN | FALSE | 0.002 | 205621 | 0.003 | | |
| tfidf_kvinde | 180 | mean | NaN | FALSE | 0.004 | 310428 | 0.003 | | X |
| tfidf_kæreste | 180 | mean | NaN | FALSE | 0.005 | 247924 | 0.003 | | |
| tfidf_kæresten | 180 | mean | NaN | FALSE | 0.002 | 128128 | 0.003 | | |
| tfidf_køre | 180 | mean | NaN | FALSE | 0.002 | 298677 | 0.003 | | |
| tfidf_kører | 180 | mean | NaN | FALSE | 0.002 | 337754 | 0.003 | | X |
| tfidf_lade | 180 | mean | NaN | FALSE | 0.002 | 350320 | 0.003 | | X |
| tfidf_lagt | 180 | mean | NaN | FALSE | 0.003 | 394657 | 0.003 | | |
| tfidf_lang | 180 | mean | NaN | FALSE | 0.003 | 432830 | 0.003 | | |
| tfidf_langt | 180 | mean | NaN | FALSE | 0.001 | 273786 | 0.003 | | |
| tfidf_latenstid | 180 | mean | NaN | FALSE | 0.005 | 373160 | 0.003 | | |
| tfidf_laver | 180 | mean | NaN | FALSE | 0.002 | 374482 | 0.003 | | X |
| tfidf_laves | 180 | mean | NaN | FALSE | 0.002 | 285571 | 0.003 | | |
| tfidf_lejlighed | 180 | mean | NaN | FALSE | 0.005 | 366245 | 0.003 | | X |
| tfidf_let | 180 | mean | NaN | FALSE | 0.009 | 536429 | 0.003 | | |
| tfidf_lettere | 180 | mean | NaN | FALSE | 0.004 | 439502 | 0.003 | | X |
| tfidf_lide | 180 | mean | NaN | FALSE | 0.001 | 247982 | 0.003 | | X |
| tfidf_ligeledes | 180 | mean | NaN | FALSE | 0.003 | 459479 | 0.003 | | X |
| tfidf_liv | 180 | mean | NaN | FALSE | 0.004 | 428300 | 0.003 | | |
| tfidf_livet | 180 | mean | NaN | FALSE | 0.002 | 327880 | 0.003 | | X |
| tfidf_lov | 180 | mean | NaN | FALSE | 0.002 | 317428 | 0.003 | | X |
| tfidf_lyst | 180 | mean | NaN | FALSE | 0.004 | 470904 | 0.003 | | X |
| tfidf_læge | 180 | mean | NaN | FALSE | 0.01 | 559291 | 0.003 | | X |
| tfidf_lægesamtale | 180 | mean | NaN | FALSE | 0.005 | 411744 | 0.003 | | |
| tfidf_længe | 180 | mean | NaN | FALSE | 0.003 | 428207 | 0.003 | | |
| tfidf_læse | 180 | mean | NaN | FALSE | 0.002 | 283907 | 0.003 | | |
| tfidf_løbet | 180 | mean | NaN | FALSE | 0.006 | 513719 | 0.003 | | |
| tfidf_mad | 180 | mean | NaN | FALSE | 0.006 | 453874 | 0.003 | | X |
| tfidf_mail | 180 | mean | NaN | FALSE | 0.001 | 159244 | 0.003 | | |
| tfidf_mandag | 180 | mean | NaN | FALSE | 0.005 | 444021 | 0.003 | | X |
| tfidf_manglende | 180 | mean | NaN | FALSE | 0.003 | 389886 | 0.003 | | |
| tfidf_mangler | 180 | mean | NaN | FALSE | 0.002 | 315480 | 0.003 | | |
| tfidf_max | 180 | mean | NaN | FALSE | 0.001 | 164307 | 0.003 | | |

| | | | | | | | | | |
|---|---|---|---|---|---|---|---|---|---|
| tfidf_mdr | 180 | mean | NaN | FALSE | 0.002 | 308478 | 0.003 | | |
| tfidf_medicin | 180 | mean | NaN | FALSE | 0.013 | 580384 | 0.003 | | X |
| tfidf_medicinen | 180 | mean | NaN | FALSE | 0.003 | 381180 | 0.003 | | X |
| tfidf_medicinsk | 180 | mean | NaN | FALSE | 0.002 | 280889 | 0.003 | | |
| tfidf_medicinsk behandling | 180 | mean | NaN | FALSE | 0.001 | 186879 | 0.003 | | |
| tfidf_medicinske | 180 | mean | NaN | FALSE | 0.001 | 202507 | 0.003 | | |
| tfidf_medicinske behandling | 180 | mean | NaN | FALSE | 0.001 | 163404 | 0.003 | | |
| tfidf_medikinet | 180 | mean | NaN | FALSE | 0 | 5850 | 0.003 | | |
| tfidf_medpt | 180 | mean | NaN | FALSE | 0.008 | 424312 | 0.003 | | X |
| tfidf_mener | 180 | mean | NaN | FALSE | 0.011 | 575299 | 0.003 | | X |
| tfidf_mennesker | 180 | mean | NaN | FALSE | 0.002 | 368026 | 0.003 | | |
| tfidf_meste | 180 | mean | NaN | FALSE | 0.007 | 492669 | 0.003 | | X |
| tfidf_fornavn* | 180 | mean | NaN | FALSE | 0.002 | 119400 | 0.003 | | |
| tfidf_mg | 180 | mean | NaN | FALSE | 0.005 | 481813 | 0.003 | | X |
| tfidf_mg pn | 180 | mean | NaN | FALSE | 0 | 76350 | 0.003 | | |
| tfidf_mhp | 180 | mean | NaN | FALSE | 0.005 | 469633 | 0.003 | | |
| tfidf_middag | 180 | mean | NaN | FALSE | 0.004 | 404989 | 0.003 | | X |
| tfidf_miljøet | 180 | mean | NaN | FALSE | 0.013 | 478611 | 0.003 | | X |
| tfidf_mimik | 180 | mean | NaN | FALSE | 0.005 | 368235 | 0.003 | | X |
| tfidf_misbrug | 180 | mean | NaN | FALSE | 0.002 | 221770 | 0.003 | | |
| tfidf_mistanke | 180 | mean | NaN | FALSE | 0.002 | 282279 | 0.003 | | X |
| tfidf_mm | 180 | mean | NaN | FALSE | 0.002 | 313704 | 0.003 | | X |
| tfidf_moderen | 180 | mean | NaN | FALSE | 0.002 | 199248 | 0.003 | | |
| tfidf_modtager | 180 | mean | NaN | FALSE | 0.002 | 280804 | 0.003 | | X |
| tfidf_modtaget | 180 | mean | NaN | FALSE | 0.002 | 291530 | 0.003 | | |
| tfidf_mor | 180 | mean | NaN | FALSE | 0.008 | 423549 | 0.003 | | |
| tfidf_morgen | 180 | mean | NaN | FALSE | 0.016 | 586149 | 0.003 | | X |
| tfidf_morgenen | 180 | mean | NaN | FALSE | 0.002 | 333039 | 0.003 | | X |
| tfidf_morgenmad | 180 | mean | NaN | FALSE | 0.004 | 378525 | 0.003 | | X |
| tfidf_motiveret | 180 | mean | NaN | FALSE | 0.002 | 309553 | 0.003 | | |
| tfidf_motorisk | 180 | mean | NaN | FALSE | 0.002 | 254239 | 0.003 | | X |
| tfidf_mulighed | 180 | mean | NaN | FALSE | 0.003 | 428837 | 0.003 | | |
| tfidf_muligheden | 180 | mean | NaN | FALSE | 0.001 | 264523 | 0.003 | | |
| tfidf_muligt | 180 | mean | NaN | FALSE | 0.004 | 461152 | 0.003 | | X |
| tfidf_muligvis | 180 | mean | NaN | FALSE | 0.002 | 322129 | 0.003 | | X |
| tfidf_musik | 180 | mean | NaN | FALSE | 0.003 | 282872 | 0.003 | | |
| tfidf_måde | 180 | mean | NaN | FALSE | 0.003 | 435584 | 0.003 | | X |
| tfidf_måned | 180 | mean | NaN | FALSE | 0.002 | 307509 | 0.003 | | |
| tfidf_måneder | 180 | mean | NaN | FALSE | 0.002 | 317147 | 0.003 | | |
| tfidf_mærke | 180 | mean | NaN | FALSE | 0.003 | 402478 | 0.003 | | X |
| tfidf_mærker | 180 | mean | NaN | FALSE | 0.001 | 262051 | 0.003 | | |
| tfidf_møde | 180 | mean | NaN | FALSE | 0.003 | 357859 | 0.003 | | |
| tfidf_møder | 180 | mean | NaN | FALSE | 0.002 | 268539 | 0.003 | | |
| tfidf_nat | 180 | mean | NaN | FALSE | 0.007 | 506344 | 0.003 | | |
| tfidf_natten | 180 | mean | NaN | FALSE | 0.005 | 496222 | 0.003 | | X |
| tfidf_nedsat | 180 | mean | NaN | FALSE | 0.009 | 459800 | 0.003 | | X |
| tfidf_negative | 180 | mean | NaN | FALSE | 0.002 | 270729 | 0.003 | | X |
| tfidf_nervøs | 180 | mean | NaN | FALSE | 0.002 | 304757 | 0.003 | | X |
| tfidf_neutral | 180 | mean | NaN | FALSE | 0.003 | 276113 | 0.003 | | |
| tfidf_neutralt | 180 | mean | NaN | FALSE | 0.006 | 463538 | 0.003 | | |
| tfidf_neutralt stemningsleje | 180 | mean | NaN | FALSE | 0.004 | 345419 | 0.003 | | |
| tfidf_normal | 180 | mean | NaN | FALSE | 0.003 | 382357 | 0.003 | | |
| tfidf_normalt | 180 | mean | NaN | FALSE | 0.003 | 412111 | 0.003 | | |
| tfidf_notat | 180 | mean | NaN | FALSE | 0.004 | 382243 | 0.003 | | |
| tfidf_nuværende | 180 | mean | NaN | FALSE | 0.003 | 440151 | 0.003 | | |
| tfidf_nærmere | 180 | mean | NaN | FALSE | 0.002 | 371627 | 0.003 | | X |
| tfidf_nævner | 180 | mean | NaN | FALSE | 0.002 | 356568 | 0.003 | | X |
| tfidf_obs | 180 | mean | NaN | FALSE | 0.004 | 405980 | 0.003 | | X |
| tfidf_observeres | 180 | mean | NaN | FALSE | 0.005 | 365763 | 0.003 | | |
| tfidf_ocd | 180 | mean | NaN | FALSE | 0.001 | 54480 | 0.003 | | |
| tfidf_ofte | 180 | mean | NaN | FALSE | 0.003 | 452503 | 0.003 | | X |
| tfidf_ok | 180 | mean | NaN | FALSE | 0.004 | 427465 | 0.003 | | X |
| tfidf_olanzapin | 180 | mean | NaN | FALSE | 0.002 | 162278 | 0.003 | | X |
| tfidf_ondt | 180 | mean | NaN | FALSE | 0.003 | 352311 | 0.003 | | |
| tfidf_onsdag | 180 | mean | NaN | FALSE | 0.003 | 317425 | 0.003 | | X |
| tfidf_opfordres | 180 | mean | NaN | FALSE | 0.002 | 352518 | 0.003 | | |
| tfidf_opfordret | 180 | mean | NaN | FALSE | 0.003 | 391869 | 0.003 | | X |
| tfidf_opfølgning | 180 | mean | NaN | FALSE | 0.002 | 241378 | 0.003 | | |
| tfidf_opgaver | 180 | mean | NaN | FALSE | 0.001 | 176092 | 0.003 | | X |
| tfidf_opholder | 180 | mean | NaN | FALSE | 0.009 | 472832 | 0.003 | | X |
| tfidf_opholdt | 180 | mean | NaN | FALSE | 0.016 | 540254 | 0.003 | | X |
| tfidf_oplevede | 180 | mean | NaN | FALSE | 0.001 | 279163 | 0.003 | | |
| tfidf_oplevelse | 180 | mean | NaN | FALSE | 0.002 | 310362 | 0.003 | | X |
| tfidf_oplevelser | 180 | mean | NaN | FALSE | 0.002 | 274071 | 0.003 | | |
| tfidf_oplever | 180 | mean | NaN | FALSE | 0.008 | 546836 | 0.003 | | X |
| tfidf_opleves | 180 | mean | NaN | FALSE | 0.004 | 400995 | 0.003 | | |

| | | | | | | | | | |
|---|---|---|---|---|---|---|---|---|---|
| tfidf_oplevet | 180 | mean | NaN | FALSE | 0.003 | 428410 | 0.003 | | X |
| tfidf_oplyser | 180 | mean | NaN | FALSE | 0.004 | 434851 | 0.003 | | |
| tfidf_opmærksom | 180 | mean | NaN | FALSE | 0.002 | 332389 | 0.003 | | |
| tfidf_opmærksomhed | 180 | mean | NaN | FALSE | 0 | 128752 | 0.003 | | X |
| tfidf_oppe | 180 | mean | NaN | FALSE | 0.005 | 416859 | 0.003 | | X |
| tfidf_opringning | 180 | mean | NaN | FALSE | 0.001 | 155142 | 0.003 | | |
| tfidf_opstart | 180 | mean | NaN | FALSE | 0.002 | 295126 | 0.003 | | X |
| tfidf_optaget | 180 | mean | NaN | FALSE | 0.002 | 268004 | 0.003 | | X |
| tfidf_ord | 180 | mean | NaN | FALSE | 0.004 | 371242 | 0.003 | | |
| tfidf_ordineret | 180 | mean | NaN | FALSE | 0.001 | 229617 | 0.003 | | |
| tfidf_orienteres | 180 | mean | NaN | FALSE | 0.001 | 180443 | 0.003 | | |
| tfidf_orienteret | 180 | mean | NaN | FALSE | 0.006 | 505682 | 0.003 | | X |
| tfidf_ovenstående | 180 | mean | NaN | FALSE | 0.001 | 227883 | 0.003 | | X |
| tfidf_overfor | 180 | mean | NaN | FALSE | 0.004 | 484001 | 0.003 | | X |
| tfidf_overlæge | 180 | mean | NaN | FALSE | 0.002 | 294299 | 0.003 | | |
| tfidf_overskud | 180 | mean | NaN | FALSE | 0.002 | 308874 | 0.003 | | X |
| tfidf_overskue | 180 | mean | NaN | FALSE | 0.003 | 320503 | 0.003 | | X |
| tfidf_ovl | 180 | mean | NaN | FALSE | 0.001 | 181809 | 0.003 | | |
| tfidf_oxapax | 180 | mean | NaN | FALSE | 0.004 | 272827 | 0.003 | | |
| tfidf_par | 180 | mean | NaN | FALSE | 0.005 | 518335 | 0.003 | | |
| tfidf_patient | 180 | mean | NaN | FALSE | 0.003 | 286382 | 0.003 | | |
| tfidf_patienten | 180 | mean | NaN | FALSE | 0.011 | 439542 | 0.003 | | X |
| tfidf_patientens | 180 | mean | NaN | FALSE | 0.001 | 207558 | 0.003 | | X |
| tfidf_penge | 180 | mean | NaN | FALSE | 0.003 | 261677 | 0.003 | | X |
| tfidf_periode | 180 | mean | NaN | FALSE | 0.002 | 365288 | 0.003 | | |
| tfidf_perioder | 180 | mean | NaN | FALSE | 0.002 | 349618 | 0.003 | | |
| tfidf_personale | 180 | mean | NaN | FALSE | 0.009 | 499245 | 0.003 | | X |
| tfidf_personalet | 180 | mean | NaN | FALSE | 0.008 | 505656 | 0.003 | | X |
| tfidf_pga | 180 | mean | NaN | FALSE | 0.004 | 468524 | 0.003 | | |
| tfidf_pige | 180 | mean | NaN | FALSE | 0 | 95972 | 0.003 | | X |
| tfidf_plads | 180 | mean | NaN | FALSE | 0.002 | 264817 | 0.003 | | |
| tfidf_plaget | 180 | mean | NaN | FALSE | 0.003 | 349662 | 0.003 | | X |
| tfidf_plan | 180 | mean | NaN | FALSE | 0.004 | 434046 | 0.003 | | X |
| tfidf_planen | 180 | mean | NaN | FALSE | 0.002 | 302895 | 0.003 | | |
| tfidf_planer | 180 | mean | NaN | FALSE | 0.005 | 459833 | 0.003 | | X |
| tfidf_planlagt | 180 | mean | NaN | FALSE | 0.003 | 339478 | 0.003 | | X |
| tfidf_plejer | 180 | mean | NaN | FALSE | 0.002 | 377978 | 0.003 | | |
| tfidf_pn | 180 | mean | NaN | FALSE | 0.011 | 498512 | 0.003 | | X |
| tfidf_politiet | 180 | mean | NaN | FALSE | 0.003 | 217443 | 0.003 | | X |
| tfidf_positiv | 180 | mean | NaN | FALSE | 0.002 | 258710 | 0.003 | | X |
| tfidf_pr | 180 | mean | NaN | FALSE | 0.001 | 252946 | 0.003 | | |
| tfidf_presset | 180 | mean | NaN | FALSE | 0.002 | 295751 | 0.003 | | |
| tfidf_primært | 180 | mean | NaN | FALSE | 0.004 | 392646 | 0.003 | | X |
| tfidf_problem | 180 | mean | NaN | FALSE | 0.001 | 250400 | 0.003 | | |
| tfidf_problemer | 180 | mean | NaN | FALSE | 0.004 | 472464 | 0.003 | | |
| tfidf_præget | 180 | mean | NaN | FALSE | 0.004 | 407257 | 0.003 | | X |
| tfidf_prøve | 180 | mean | NaN | FALSE | 0.003 | 375092 | 0.003 | | X |
| tfidf_prøver | 180 | mean | NaN | FALSE | 0.002 | 344755 | 0.003 | | X |
| tfidf_psykiatrisk | 180 | mean | NaN | FALSE | 0.003 | 313491 | 0.003 | | X |
| tfidf_psykisk | 180 | mean | NaN | FALSE | 0.005 | 465152 | 0.003 | | |
| tfidf_psykiske | 180 | mean | NaN | FALSE | 0.002 | 330805 | 0.003 | | |
| tfidf_psykolog | 180 | mean | NaN | FALSE | 0.002 | 247565 | 0.003 | | X |
| tfidf_psykomotorisk | 180 | mean | NaN | FALSE | 0.006 | 493267 | 0.003 | | |
| tfidf_psykomotorisk tempo | 180 | mean | NaN | FALSE | 0.004 | 393446 | 0.003 | | |
| tfidf_psykotisk | 180 | mean | NaN | FALSE | 0.011 | 534209 | 0.003 | | X |
| tfidf_psykotiske | 180 | mean | NaN | FALSE | 0.006 | 453671 | 0.003 | | X |
| tfidf_psykotiske symptomer | 180 | mean | NaN | FALSE | 0.003 | 386714 | 0.003 | | X |
| tfidf_pt | 180 | mean | NaN | FALSE | 0.108 | 645471 | 0.003 | | |
| tfidf_pt angiver | 180 | mean | NaN | FALSE | 0.004 | 422730 | 0.003 | | |
| tfidf_pt beskriver | 180 | mean | NaN | FALSE | 0.002 | 302928 | 0.003 | | X |
| tfidf_pt fortæller | 180 | mean | NaN | FALSE | 0.01 | 527637 | 0.003 | | X |
| tfidf_pt fremstår | 180 | mean | NaN | FALSE | 0.006 | 424433 | 0.003 | | |
| tfidf_pt fået | 180 | mean | NaN | FALSE | 0.003 | 377425 | 0.003 | | |
| tfidf_pt føler | 180 | mean | NaN | FALSE | 0.002 | 318462 | 0.003 | | X |
| tfidf_pt oplever | 180 | mean | NaN | FALSE | 0.002 | 279989 | 0.003 | | X |
| tfidf_pt pt | 180 | mean | NaN | FALSE | 0.003 | 377920 | 0.003 | | X |
| tfidf_pt svært | 180 | mean | NaN | FALSE | 0.002 | 340717 | 0.003 | | |
| tfidf_pt tager | 180 | mean | NaN | FALSE | 0.002 | 269980 | 0.003 | | |
| tfidf_pt ut | 180 | mean | NaN | FALSE | 0.004 | 326260 | 0.003 | | |
| tfidf_pt ønsker | 180 | mean | NaN | FALSE | 0.004 | 398060 | 0.003 | | X |
| tfidf_pts | 180 | mean | NaN | FALSE | 0.009 | 541475 | 0.003 | | X |
| tfidf_ptt | 180 | mean | NaN | FALSE | 0.002 | 188921 | 0.003 | | |
| tfidf_påvirket | 180 | mean | NaN | FALSE | 0.004 | 420373 | 0.003 | | |
| tfidf_quetiapin | 180 | mean | NaN | FALSE | 0.003 | 273035 | 0.003 | | |
| tfidf_recept | 180 | mean | NaN | FALSE | 0 | 57745 | 0.003 | | |
| tfidf_relevant | 180 | mean | NaN | FALSE | 0.01 | 550586 | 0.003 | | |

| | | | | | | | | | |
|---|---|---|---|---|---|---|---|---|---|
| tfidf_retur | 180 | mean | NaN | FALSE | 0.009 | 409023 | 0.003 | | X |
| tfidf_rigtig | 180 | mean | NaN | FALSE | 0.006 | 514296 | 0.003 | | |
| tfidf_ringe | 180 | mean | NaN | FALSE | 0.004 | 434077 | 0.003 | | X |
| tfidf_ringer | 180 | mean | NaN | FALSE | 0.004 | 399047 | 0.003 | | X |
| tfidf_ringet | 180 | mean | NaN | FALSE | 0.003 | 356734 | 0.003 | | X |
| tfidf_rolig | 180 | mean | NaN | FALSE | 0.013 | 537358 | 0.003 | | X |
| tfidf_roligt | 180 | mean | NaN | FALSE | 0.003 | 348212 | 0.003 | | X |
| tfidf_rp | 180 | mean | NaN | FALSE | 0 | 58726 | 0.003 | | |
| tfidf_ryge | 180 | mean | NaN | FALSE | 0.005 | 292101 | 0.003 | | X |
| tfidf_sagsbehandler | 180 | mean | NaN | FALSE | 0.001 | 172189 | 0.003 | | |
| tfidf_samarbejde | 180 | mean | NaN | FALSE | 0.002 | 320206 | 0.003 | | X |
| tfidf_samlet | 180 | mean | NaN | FALSE | 0.006 | 462912 | 0.003 | | |
| tfidf_samt | 180 | mean | NaN | FALSE | 0.015 | 602047 | 0.003 | | |
| tfidf_samtale | 180 | mean | NaN | FALSE | 0.024 | 614526 | 0.003 | | X |
| tfidf_samtale pt | 180 | mean | NaN | FALSE | 0.004 | 398963 | 0.003 | | |
| tfidf_samtalen | 180 | mean | NaN | FALSE | 0.014 | 591096 | 0.003 | | X |
| tfidf_samtaler | 180 | mean | NaN | FALSE | 0.004 | 424786 | 0.003 | | X |
| tfidf_samtidig | 180 | mean | NaN | FALSE | 0.002 | 401950 | 0.003 | | |
| tfidf_sat | 180 | mean | NaN | FALSE | 0.003 | 375815 | 0.003 | | |
| tfidf_selvmord | 180 | mean | NaN | FALSE | 0.002 | 291869 | 0.003 | | |
| tfidf_selvmordstanker | 180 | mean | NaN | FALSE | 0.007 | 457139 | 0.003 | | X |
| tfidf_selvskade | 180 | mean | NaN | FALSE | 0.002 | 173752 | 0.003 | | |
| tfidf_sendes | 180 | mean | NaN | FALSE | 0.001 | 146278 | 0.003 | | |
| tfidf_sendt | 180 | mean | NaN | FALSE | 0.002 | 286793 | 0.003 | | |
| tfidf_seneste | 180 | mean | NaN | FALSE | 0.004 | 448715 | 0.003 | | |
| tfidf_seng | 180 | mean | NaN | FALSE | 0.008 | 495156 | 0.003 | | |
| tfidf_sengen | 180 | mean | NaN | FALSE | 0.009 | 483488 | 0.003 | | X |
| tfidf_seroquel | 180 | mean | NaN | FALSE | 0.001 | 90412 | 0.003 | | |
| tfidf_sertralin | 180 | mean | NaN | FALSE | 0.001 | 95784 | 0.003 | | |
| tfidf_sidde | 180 | mean | NaN | FALSE | 0.003 | 374847 | 0.003 | | |
| tfidf_sidder | 180 | mean | NaN | FALSE | 0.008 | 525800 | 0.003 | | X |
| tfidf_siddet | 180 | mean | NaN | FALSE | 0.007 | 429293 | 0.003 | | X |
| tfidf_side | 180 | mean | NaN | FALSE | 0.002 | 347594 | 0.003 | | |
| tfidf_sidst | 180 | mean | NaN | FALSE | 0.005 | 491807 | 0.003 | | X |
| tfidf_sidste | 180 | mean | NaN | FALSE | 0.009 | 583984 | 0.003 | | |
| tfidf_sidste uge | 180 | mean | NaN | FALSE | 0.001 | 271783 | 0.003 | | |
| tfidf_situation | 180 | mean | NaN | FALSE | 0.003 | 394712 | 0.003 | | |
| tfidf_situationen | 180 | mean | NaN | FALSE | 0.002 | 324890 | 0.003 | | |
| tfidf_situationer | 180 | mean | NaN | FALSE | 0.001 | 186640 | 0.003 | | |
| tfidf_sket | 180 | mean | NaN | FALSE | 0.001 | 303398 | 0.003 | | X |
| tfidf_skidt | 180 | mean | NaN | FALSE | 0.003 | 355920 | 0.003 | | X |
| tfidf_skole | 180 | mean | NaN | FALSE | 0.001 | 106394 | 0.003 | | |
| tfidf_skolen | 180 | mean | NaN | FALSE | 0.001 | 94971 | 0.003 | | |
| tfidf_skrevet | 180 | mean | NaN | FALSE | 0.003 | 357039 | 0.003 | | |
| tfidf_skyldes | 180 | mean | NaN | FALSE | 0.002 | 329926 | 0.003 | | |
| tfidf_slet | 180 | mean | NaN | FALSE | 0.002 | 333417 | 0.003 | | |
| tfidf_smerter | 180 | mean | NaN | FALSE | 0.005 | 321949 | 0.003 | | |
| tfidf_smil | 180 | mean | NaN | FALSE | 0.005 | 374074 | 0.003 | | X |
| tfidf_smilende | 180 | mean | NaN | FALSE | 0.01 | 462513 | 0.003 | | X |
| tfidf_smule | 180 | mean | NaN | FALSE | 0.003 | 364328 | 0.003 | | |
| tfidf_snak | 180 | mean | NaN | FALSE | 0.004 | 399693 | 0.003 | | X |
| tfidf_snakke | 180 | mean | NaN | FALSE | 0.005 | 462680 | 0.003 | | X |
| tfidf_snakker | 180 | mean | NaN | FALSE | 0.005 | 457875 | 0.003 | | |
| tfidf_snakket | 180 | mean | NaN | FALSE | 0.004 | 397495 | 0.003 | | X |
| tfidf_social | 180 | mean | NaN | FALSE | 0.005 | 381489 | 0.003 | | X |
| tfidf_sociale | 180 | mean | NaN | FALSE | 0.002 | 274673 | 0.003 | | X |
| tfidf_socialt | 180 | mean | NaN | FALSE | 0.001 | 192537 | 0.003 | | |
| tfidf_somatisk | 180 | mean | NaN | FALSE | 0.003 | 264012 | 0.003 | | X |
| tfidf_sove | 180 | mean | NaN | FALSE | 0.006 | 496858 | 0.003 | | |
| tfidf_sover | 180 | mean | NaN | FALSE | 0.009 | 524772 | 0.003 | | |
| tfidf_sovet | 180 | mean | NaN | FALSE | 0.014 | 551216 | 0.003 | | |
| tfidf_spille | 180 | mean | NaN | FALSE | 0.002 | 231423 | 0.003 | | |
| tfidf_spiller | 180 | mean | NaN | FALSE | 0.001 | 204243 | 0.003 | | |
| tfidf_spillet | 180 | mean | NaN | FALSE | 0.004 | 240272 | 0.003 | | X |
| tfidf_spise | 180 | mean | NaN | FALSE | 0.006 | 470078 | 0.003 | | |
| tfidf_spiser | 180 | mean | NaN | FALSE | 0.005 | 446238 | 0.003 | | |
| tfidf_spist | 180 | mean | NaN | FALSE | 0.007 | 456314 | 0.003 | | |
| tfidf_spurgt | 180 | mean | NaN | FALSE | 0.002 | 349578 | 0.003 | | |
| tfidf_spørger | 180 | mean | NaN | FALSE | 0.008 | 533981 | 0.003 | | X |
| tfidf_spørgsmål | 180 | mean | NaN | FALSE | 0.005 | 466065 | 0.003 | | X |
| tfidf_stabil | 180 | mean | NaN | FALSE | 0.001 | 201511 | 0.003 | | |
| tfidf_stand | 180 | mean | NaN | FALSE | 0.001 | 286382 | 0.003 | | |
| tfidf_start | 180 | mean | NaN | FALSE | 0.002 | 321554 | 0.003 | | X |
| tfidf_starte | 180 | mean | NaN | FALSE | 0.001 | 270475 | 0.003 | | |
| tfidf_starten | 180 | mean | NaN | FALSE | 0.003 | 373395 | 0.003 | | |
| tfidf_starter | 180 | mean | NaN | FALSE | 0.001 | 245910 | 0.003 | | |

| | | | | | | | | | |
|---|---|---|---|---|---|---|---|---|---|
| tfidf_startet | 180 | mean | NaN | FALSE | 0.001 | 193523 | 0.003 | | |
| tfidf_sted | 180 | mean | NaN | FALSE | 0.003 | 402804 | 0.003 | | X |
| tfidf_stede | 180 | mean | NaN | FALSE | 0.005 | 363997 | 0.003 | | |
| tfidf_stedet | 180 | mean | NaN | FALSE | 0.002 | 381265 | 0.003 | | |
| tfidf_stemmehøring | 180 | mean | NaN | FALSE | 0.003 | 228449 | 0.003 | | |
| tfidf_stemmer | 180 | mean | NaN | FALSE | 0.004 | 285068 | 0.003 | | |
| tfidf_stemmerne | 180 | mean | NaN | FALSE | 0.003 | 153557 | 0.003 | | |
| tfidf_stemningsleje | 180 | mean | NaN | FALSE | 0.013 | 550311 | 0.003 | | X |
| tfidf_stemningslejet | 180 | mean | NaN | FALSE | 0.003 | 347537 | 0.003 | | |
| tfidf_stille | 180 | mean | NaN | FALSE | 0.011 | 531438 | 0.003 | | |
| tfidf_stk | 180 | mean | NaN | FALSE | 0.001 | 188076 | 0.003 | | |
| tfidf_stoppe | 180 | mean | NaN | FALSE | 0.001 | 252821 | 0.003 | | X |
| tfidf_strattera | 180 | mean | NaN | FALSE | 0 | 20628 | 0.003 | | |
| tfidf_struktur | 180 | mean | NaN | FALSE | 0.001 | 197705 | 0.003 | | |
| tfidf_stue | 180 | mean | NaN | FALSE | 0.02 | 560985 | 0.003 | | |
| tfidf_stuen | 180 | mean | NaN | FALSE | 0.023 | 588977 | 0.003 | | X |
| tfidf_stå | 180 | mean | NaN | FALSE | 0.003 | 365098 | 0.003 | | |
| tfidf_står | 180 | mean | NaN | FALSE | 0.005 | 475302 | 0.003 | | X |
| tfidf_større | 180 | mean | NaN | FALSE | 0.001 | 237706 | 0.003 | | |
| tfidf_støtte | 180 | mean | NaN | FALSE | 0.004 | 405885 | 0.003 | | X |
| tfidf_svar | 180 | mean | NaN | FALSE | 0.003 | 378445 | 0.003 | | |
| tfidf_svare | 180 | mean | NaN | FALSE | 0.003 | 354796 | 0.003 | | X |
| tfidf_svarer | 180 | mean | NaN | FALSE | 0.009 | 539143 | 0.003 | | X |
| tfidf_svingende | 180 | mean | NaN | FALSE | 0.002 | 271817 | 0.003 | | X |
| tfidf_svær | 180 | mean | NaN | FALSE | 0.004 | 436604 | 0.003 | | X |
| tfidf_svære | 180 | mean | NaN | FALSE | 0.001 | 234338 | 0.003 | | X |
| tfidf_svært | 180 | mean | NaN | FALSE | 0.016 | 604916 | 0.003 | | |
| tfidf_syg | 180 | mean | NaN | FALSE | 0.002 | 319114 | 0.003 | | X |
| tfidf_sygdom | 180 | mean | NaN | FALSE | 0.002 | 290460 | 0.003 | | |
| tfidf_symptomer | 180 | mean | NaN | FALSE | 0.008 | 545821 | 0.003 | | |
| tfidf_såfremt | 180 | mean | NaN | FALSE | 0.002 | 337666 | 0.003 | | |
| tfidf_særligt | 180 | mean | NaN | FALSE | 0.001 | 301831 | 0.003 | | |
| tfidf_sætte | 180 | mean | NaN | FALSE | 0.002 | 357629 | 0.003 | | |
| tfidf_sætter | 180 | mean | NaN | FALSE | 0.002 | 319911 | 0.003 | | X |
| tfidf_søn | 180 | mean | NaN | FALSE | 0.004 | 172800 | 0.003 | | |
| tfidf_søster | 180 | mean | NaN | FALSE | 0.004 | 227982 | 0.003 | | |
| tfidf_søvn | 180 | mean | NaN | FALSE | 0.005 | 480432 | 0.003 | | X |
| tfidf_tabl | 180 | mean | NaN | FALSE | 0.001 | 152468 | 0.003 | | X |
| tfidf_tager | 180 | mean | NaN | FALSE | 0.009 | 573952 | 0.003 | | X |
| tfidf_tages | 180 | mean | NaN | FALSE | 0.002 | 356710 | 0.003 | | |
| tfidf_taget | 180 | mean | NaN | FALSE | 0.01 | 572455 | 0.003 | | |
| tfidf_tale | 180 | mean | NaN | FALSE | 0.011 | 574415 | 0.003 | | X |
| tfidf_taler | 180 | mean | NaN | FALSE | 0.011 | 579352 | 0.003 | | X |
| tfidf_tales | 180 | mean | NaN | FALSE | 0.002 | 299006 | 0.003 | | X |
| tfidf_talt | 180 | mean | NaN | FALSE | 0.006 | 507754 | 0.003 | | X |
| tfidf_tankemylder | 180 | mean | NaN | FALSE | 0.002 | 270047 | 0.003 | | X |
| tfidf_tanker | 180 | mean | NaN | FALSE | 0.012 | 558620 | 0.003 | | X |
| tfidf_tankerne | 180 | mean | NaN | FALSE | 0.003 | 360688 | 0.003 | | X |
| tfidf_tbl | 180 | mean | NaN | FALSE | 0.002 | 238150 | 0.003 | | |
| tfidf_tegn | 180 | mean | NaN | FALSE | 0.002 | 341994 | 0.003 | | |
| tfidf_telefon | 180 | mean | NaN | FALSE | 0.003 | 328561 | 0.003 | | X |
| tfidf_telefonisk | 180 | mean | NaN | FALSE | 0.001 | 224848 | 0.003 | | |
| tfidf_tempo | 180 | mean | NaN | FALSE | 0.005 | 455409 | 0.003 | | |
| tfidf_tendens | 180 | mean | NaN | FALSE | 0.001 | 264007 | 0.003 | | |
| tfidf_tid | 180 | mean | NaN | FALSE | 0.014 | 604152 | 0.003 | | |
| tfidf_tiden | 180 | mean | NaN | FALSE | 0.007 | 550956 | 0.003 | | |
| tfidf_tider | 180 | mean | NaN | FALSE | 0.002 | 320410 | 0.003 | | X |
| tfidf_tidl | 180 | mean | NaN | FALSE | 0.003 | 360742 | 0.003 | | |
| tfidf_tidspunkt | 180 | mean | NaN | FALSE | 0.003 | 458930 | 0.003 | | X |
| tfidf_tilbagemelding | 180 | mean | NaN | FALSE | 0.001 | 86816 | 0.003 | | |
| tfidf_tilbud | 180 | mean | NaN | FALSE | 0.001 | 232642 | 0.003 | | |
| tfidf_tilbudt | 180 | mean | NaN | FALSE | 0.003 | 391008 | 0.003 | | X |
| tfidf_tilbydes | 180 | mean | NaN | FALSE | 0.003 | 379782 | 0.003 | | X |
| tfidf_tilstand | 180 | mean | NaN | FALSE | 0.007 | 523085 | 0.003 | | X |
| tfidf_tilstanden | 180 | mean | NaN | FALSE | 0.002 | 252672 | 0.003 | | |
| tfidf_tilstede | 180 | mean | NaN | FALSE | 0.004 | 344275 | 0.003 | | |
| tfidf_tilsyn | 180 | mean | NaN | FALSE | 0.013 | 444350 | 0.003 | | X |
| tfidf_tiltagende | 180 | mean | NaN | FALSE | 0.005 | 460569 | 0.003 | | X |
| tfidf_time | 180 | mean | NaN | FALSE | 0.004 | 434943 | 0.003 | | |
| tfidf_timer | 180 | mean | NaN | FALSE | 0.005 | 480957 | 0.003 | | |
| tfidf_ting | 180 | mean | NaN | FALSE | 0.008 | 553787 | 0.003 | | X |
| tfidf_tingene | 180 | mean | NaN | FALSE | 0.002 | 296629 | 0.003 | | |
| tfidf_tirsdag | 180 | mean | NaN | FALSE | 0.003 | 317885 | 0.003 | | X |
| tfidf_tlf | 180 | mean | NaN | FALSE | 0.002 | 303042 | 0.003 | | |
| tfidf_tog | 180 | mean | NaN | FALSE | 0.003 | 399252 | 0.003 | | |
| tfidf_torsdag | 180 | mean | NaN | FALSE | 0.003 | 328827 | 0.003 | | X |

| | | | | | | | | | |
|---|---|---|---|---|---|---|---|---|---|
| tfidf_trist | 180 | mean | NaN | FALSE | 0.006 | 423949 | 0.003 | | X |
| tfidf_trods | 180 | mean | NaN | FALSE | 0.003 | 429088 | 0.003 | | X |
| tfidf_tror | 180 | mean | NaN | FALSE | 0.003 | 405145 | 0.003 | | X |
| tfidf_truxal | 180 | mean | NaN | FALSE | 0.001 | 65916 | 0.003 | | |
| tfidf_træt | 180 | mean | NaN | FALSE | 0.009 | 528120 | 0.003 | | X |
| tfidf_træthed | 180 | mean | NaN | FALSE | 0.001 | 210513 | 0.003 | | X |
| tfidf_tur | 180 | mean | NaN | FALSE | 0.009 | 478314 | 0.003 | | X |
| tfidf_tv | 180 | mean | NaN | FALSE | 0.008 | 457407 | 0.003 | | X |
| tfidf_tvivl | 180 | mean | NaN | FALSE | 0.002 | 318433 | 0.003 | | |
| tfidf_tydeligt | 180 | mean | NaN | FALSE | 0.003 | 375564 | 0.003 | | X |
| tfidf_tænke | 180 | mean | NaN | FALSE | 0.002 | 361839 | 0.003 | | X |
| tfidf_tænker | 180 | mean | NaN | FALSE | 0.003 | 453819 | 0.003 | | |
| tfidf_tøj | 180 | mean | NaN | FALSE | 0.005 | 405891 | 0.003 | | X |
| tfidf_udenfor | 180 | mean | NaN | FALSE | 0.002 | 308367 | 0.003 | | X |
| tfidf_udgang | 180 | mean | NaN | FALSE | 0.008 | 397079 | 0.003 | | X |
| tfidf_udleveret | 180 | mean | NaN | FALSE | 0.004 | 387111 | 0.003 | | |
| tfidf_udredning | 180 | mean | NaN | FALSE | 0.001 | 175225 | 0.003 | | |
| tfidf_udskrevet | 180 | mean | NaN | FALSE | 0.003 | 383693 | 0.003 | | X |
| tfidf_udskrivelse | 180 | mean | NaN | FALSE | 0.005 | 410055 | 0.003 | | X |
| tfidf_udskrives | 180 | mean | NaN | FALSE | 0.003 | 354642 | 0.003 | | |
| tfidf_udtryk | 180 | mean | NaN | FALSE | 0.008 | 537309 | 0.003 | | |
| tfidf_udtrykker | 180 | mean | NaN | FALSE | 0.003 | 377315 | 0.003 | | |
| tfidf_uge | 180 | mean | NaN | FALSE | 0.006 | 506663 | 0.003 | | X |
| tfidf_ugen | 180 | mean | NaN | FALSE | 0.002 | 296456 | 0.003 | | |
| tfidf_uger | 180 | mean | NaN | FALSE | 0.003 | 429205 | 0.003 | | |
| tfidf_umiddelbart | 180 | mean | NaN | FALSE | 0.003 | 427495 | 0.003 | | X |
| tfidf_undersøgelse | 180 | mean | NaN | FALSE | 0.001 | 185763 | 0.003 | | X |
| tfidf_undertegnede | 180 | mean | NaN | FALSE | 0.003 | 257403 | 0.003 | | |
| tfidf_upåfaldende | 180 | mean | NaN | FALSE | 0.003 | 283361 | 0.003 | | X |
| tfidf_uro | 180 | mean | NaN | FALSE | 0.006 | 454164 | 0.003 | | X |
| tfidf_urolig | 180 | mean | NaN | FALSE | 0.005 | 384679 | 0.003 | | X |
| tfidf_usikker | 180 | mean | NaN | FALSE | 0.001 | 249481 | 0.003 | | |
| tfidf_ut | 180 | mean | NaN | FALSE | 0.04 | 625835 | 0.003 | | X |
| tfidf_ut pt | 180 | mean | NaN | FALSE | 0.003 | 343768 | 0.003 | | |
| tfidf_ut spørger | 180 | mean | NaN | FALSE | 0.003 | 349701 | 0.003 | | X |
| tfidf_uændret | 180 | mean | NaN | FALSE | 0.004 | 302023 | 0.003 | | X |
| tfidf_vagten | 180 | mean | NaN | FALSE | 0.014 | 526651 | 0.003 | | |
| tfidf_vanlig | 180 | mean | NaN | FALSE | 0.004 | 261623 | 0.003 | | X |
| tfidf_vanligt | 180 | mean | NaN | FALSE | 0.004 | 360553 | 0.003 | | X |
| tfidf_vanskeligheder | 180 | mean | NaN | FALSE | 0.001 | 151438 | 0.003 | | |
| tfidf_vanskeligt | 180 | mean | NaN | FALSE | 0.001 | 204067 | 0.003 | | |
| tfidf_vedr | 180 | mean | NaN | FALSE | 0.002 | 341082 | 0.003 | | |
| tfidf_vej | 180 | mean | NaN | FALSE | 0.002 | 327570 | 0.003 | | X |
| tfidf_velbefindende | 180 | mean | NaN | FALSE | 0.003 | 276789 | 0.003 | | |
| tfidf_ven | 180 | mean | NaN | FALSE | 0.002 | 209167 | 0.003 | | |
| tfidf_veninde | 180 | mean | NaN | FALSE | 0.003 | 222115 | 0.003 | | |
| tfidf_venlafaxin | 180 | mean | NaN | FALSE | 0 | 48925 | 0.003 | | |
| tfidf_venlig | 180 | mean | NaN | FALSE | 0.023 | 576138 | 0.003 | | X |
| tfidf_venlig imødekommende | 180 | mean | NaN | FALSE | 0.005 | 368264 | 0.003 | | X |
| tfidf_venlig kontakten | 180 | mean | NaN | FALSE | 0.01 | 459341 | 0.003 | | X |
| tfidf_venner | 180 | mean | NaN | FALSE | 0.002 | 301277 | 0.003 | | |
| tfidf_vente | 180 | mean | NaN | FALSE | 0.002 | 306496 | 0.003 | | |
| tfidf_vide | 180 | mean | NaN | FALSE | 0.003 | 398247 | 0.003 | | X |
| tfidf_vigtigt | 180 | mean | NaN | FALSE | 0.002 | 313086 | 0.003 | | |
| tfidf_virker | 180 | mean | NaN | FALSE | 0.012 | 572614 | 0.003 | | X |
| tfidf_virkning | 180 | mean | NaN | FALSE | 0.002 | 279696 | 0.003 | | |
| tfidf_viser | 180 | mean | NaN | FALSE | 0.002 | 373354 | 0.003 | | X |
| tfidf_vko | 180 | mean | NaN | FALSE | 0.006 | 418707 | 0.003 | | |
| tfidf_vrangforestillinger | 180 | mean | NaN | FALSE | 0.004 | 369533 | 0.003 | | X |
| tfidf_vred | 180 | mean | NaN | FALSE | 0.004 | 344244 | 0.003 | | X |
| tfidf_vurdere | 180 | mean | NaN | FALSE | 0.001 | 268686 | 0.003 | | X |
| tfidf_vurderes | 180 | mean | NaN | FALSE | 0.006 | 507023 | 0.003 | | X |
| tfidf_vurdering | 180 | mean | NaN | FALSE | 0.003 | 335315 | 0.003 | | |
| tfidf_vågen | 180 | mean | NaN | FALSE | 0.008 | 541375 | 0.003 | | X |
| tfidf_vågen klar | 180 | mean | NaN | FALSE | 0.004 | 441572 | 0.003 | | |
| tfidf_vågner | 180 | mean | NaN | FALSE | 0.002 | 346668 | 0.003 | | |
| tfidf_vægt | 180 | mean | NaN | FALSE | 0.001 | 157166 | 0.003 | | |
| tfidf_væk | 180 | mean | NaN | FALSE | 0.002 | 396193 | 0.003 | | X |
| tfidf_værelset | 180 | mean | NaN | FALSE | 0.002 | 229577 | 0.003 | | |
| tfidf_weekenden | 180 | mean | NaN | FALSE | 0.004 | 405924 | 0.003 | | X |
| tfidf_yderligere | 180 | mean | NaN | FALSE | 0.003 | 441886 | 0.003 | | X |
| tfidf_åben | 180 | mean | NaN | FALSE | 0.003 | 327642 | 0.003 | | X |
| tfidf_ægtefælle | 180 | mean | NaN | FALSE | 0.003 | 114255 | 0.003 | | |
| tfidf_øges | 180 | mean | NaN | FALSE | 0 | 131629 | 0.003 | | |
| tfidf_øget | 180 | mean | NaN | FALSE | 0.003 | 416310 | 0.003 | | X |
| tfidf_øgning | 180 | mean | NaN | FALSE | 0.001 | 162608 | 0.003 | | |

| | | | | | | | | | |
|---|---|---|---|---|---|---|---|---|---|
| tfidf_øjenkontakt | 180 | mean | NaN | FALSE | 0.009 | 487915 | 0.003 | | X |
| tfidf_ønske | 180 | mean | NaN | FALSE | 0.005 | 502726 | 0.003 | | |
| tfidf_ønsker | 180 | mean | NaN | FALSE | 0.018 | 605521 | 0.003 | | X |
| tfidf_ønsket | 180 | mean | NaN | FALSE | 0.003 | 343257 | 0.003 | | |
| pregabaline | 1 | boolean | 0 | FALSE | 0.078 | 2 | 0.922 | X | |
| pregabaline | 1 | count | 0 | FALSE | 0.159 | 8 | 0.922 | X | |
| pregabaline | 3 | count | 0 | FALSE | 0.45 | 20 | 0.92 | | |
| pregabaline | 3 | boolean | 0 | FALSE | 0.08 | 2 | 0.92 | | |
| pregabaline | 7 | boolean | 0 | FALSE | 0.081 | 2 | 0.919 | X | |
| pregabaline | 7 | count | 0 | FALSE | 0.955 | 41 | 0.919 | X | |
| pregabaline | 10 | count | 0 | FALSE | 1.283 | 57 | 0.917 | | |
| pregabaline | 10 | boolean | 0 | FALSE | 0.083 | 2 | 0.917 | | |
| pregabaline | 30 | boolean | 0 | FALSE | 0.088 | 2 | 0.912 | X | |
| pregabaline | 30 | count | 0 | FALSE | 2.874 | 144 | 0.912 | X | X |
| pregabaline | 180 | boolean | 0 | FALSE | 0.102 | 2 | 0.898 | | X |
| pregabaline | 180 | count | 0 | FALSE | 8.161 | 666 | 0.898 | | |
| pregabaline | 365 | count | 0 | FALSE | 12.155 | 1177 | 0.888 | | |
| pregabaline | 365 | boolean | 0 | FALSE | 0.112 | 2 | 0.888 | | X |
| pregabaline | 730 | count | 0 | FALSE | 18.07 | 1447 | 0.87 | X | |
| pregabaline | 730 | boolean | 0 | FALSE | 0.13 | 2 | 0.87 | X | X |
| pregabaline_engangs | 1 | count | 0 | FALSE | 0 | 3 | 1 | | |
| pregabaline_engangs | 1 | boolean | 0 | FALSE | 0 | 2 | 1 | | |
| pregabaline_engangs | 3 | count | 0 | FALSE | 0.001 | 4 | 0.999 | | |
| pregabaline_engangs | 3 | boolean | 0 | FALSE | 0.001 | 2 | 0.999 | | |
| pregabaline_engangs | 7 | count | 0 | FALSE | 0.002 | 4 | 0.998 | X | |
| pregabaline_engangs | 7 | boolean | 0 | FALSE | 0.002 | 2 | 0.998 | X | |
| pregabaline_engangs | 10 | count | 0 | FALSE | 0.003 | 4 | 0.997 | | |
| pregabaline_engangs | 10 | boolean | 0 | FALSE | 0.003 | 2 | 0.997 | | |
| pregabaline_engangs | 30 | boolean | 0 | FALSE | 0.006 | 2 | 0.994 | X | |
| pregabaline_engangs | 30 | count | 0 | FALSE | 0.007 | 4 | 0.994 | | |
| pregabaline_engangs | 180 | count | 0 | FALSE | 0.016 | 6 | 0.988 | | |
| pregabaline_engangs | 180 | boolean | 0 | FALSE | 0.012 | 2 | 0.988 | | |
| pregabaline_engangs | 365 | count | 0 | FALSE | 0.021 | 6 | 0.984 | | |
| pregabaline_engangs | 365 | boolean | 0 | FALSE | 0.016 | 2 | 0.984 | | |
| pregabaline_engangs | 730 | count | 0 | FALSE | 0.029 | 7 | 0.978 | X | |
| pregabaline_engangs | 730 | boolean | 0 | FALSE | 0.022 | 2 | 0.978 | X | |
| pregabaline_fast | 1 | boolean | 0 | FALSE | 0.077 | 2 | 0.923 | X | |
| pregabaline_fast | 1 | count | 0 | FALSE | 0.157 | 7 | 0.923 | X | |
| pregabaline_fast | 3 | count | 0 | FALSE | 0.446 | 18 | 0.921 | | |
| pregabaline_fast | 3 | boolean | 0 | FALSE | 0.079 | 2 | 0.921 | | |
| pregabaline_fast | 7 | boolean | 0 | FALSE | 0.081 | 2 | 0.919 | X | |
| pregabaline_fast | 7 | count | 0 | FALSE | 0.947 | 38 | 0.919 | X | |
| pregabaline_fast | 10 | boolean | 0 | FALSE | 0.082 | 2 | 0.918 | | |
| pregabaline_fast | 10 | count | 0 | FALSE | 1.273 | 51 | 0.918 | | |
| pregabaline_fast | 30 | boolean | 0 | FALSE | 0.087 | 2 | 0.913 | X | |
| pregabaline_fast | 30 | count | 0 | FALSE | 2.851 | 136 | 0.913 | X | X |
| pregabaline_fast | 180 | count | 0 | FALSE | 8.092 | 662 | 0.9 | | X |
| pregabaline_fast | 180 | boolean | 0 | FALSE | 0.1 | 2 | 0.9 | | |
| pregabaline_fast | 365 | boolean | 0 | FALSE | 0.111 | 2 | 0.889 | | X |
| pregabaline_fast | 365 | count | 0 | FALSE | 12.042 | 1123 | 0.889 | | |
| pregabaline_fast | 730 | count | 0 | FALSE | 17.906 | 1413 | 0.872 | X | |
| pregabaline_fast | 730 | boolean | 0 | FALSE | 0.128 | 2 | 0.872 | X | X |
| pregabaline_pn | 1 | boolean | 0 | FALSE | 0.001 | 2 | 0.999 | X | |
| pregabaline_pn | 1 | count | 0 | FALSE | 0.001 | 4 | 0.999 | X | |
| pregabaline_pn | 3 | boolean | 0 | FALSE | 0.001 | 2 | 0.999 | | |
| pregabaline_pn | 3 | count | 0 | FALSE | 0.003 | 9 | 0.999 | | |
| pregabaline_pn | 7 | boolean | 0 | FALSE | 0.002 | 2 | 0.998 | X | |
| pregabaline_pn | 7 | count | 0 | FALSE | 0.005 | 16 | 0.998 | X | |
| pregabaline_pn | 10 | boolean | 0 | FALSE | 0.002 | 2 | 0.998 | | |
| pregabaline_pn | 10 | count | 0 | FALSE | 0.007 | 19 | 0.998 | | |
| pregabaline_pn | 30 | count | 0 | FALSE | 0.015 | 32 | 0.998 | X | |
| pregabaline_pn | 30 | boolean | 0 | FALSE | 0.002 | 2 | 0.998 | X | |
| pregabaline_pn | 180 | count | 0 | FALSE | 0.052 | 96 | 0.996 | | |
| pregabaline_pn | 180 | boolean | 0 | FALSE | 0.004 | 2 | 0.996 | | |
| pregabaline_pn | 365 | count | 0 | FALSE | 0.091 | 127 | 0.995 | | |
| pregabaline_pn | 365 | boolean | 0 | FALSE | 0.005 | 2 | 0.995 | | |
| pregabaline_pn | 730 | boolean | 0 | FALSE | 0.007 | 2 | 0.993 | X | |
| pregabaline_pn | 730 | count | 0 | FALSE | 0.133 | 121 | 0.993 | X | |
| remme | 730 | boolean | 0 | FALSE | 0.02 | 2 | 0.98 | X | X |
| remme | 730 | summed | 0 | FALSE | 0.866 | 388 | 0.98 | X | X |
| remme | 730 | count | 0 | FALSE | 0.037 | 26 | 0.98 | X | X |
| sedative_antipsychotics | 1 | count | 0 | FALSE | 0.979 | 13 | 0.496 | X | X |
| sedative_antipsychotics | 1 | boolean | 0 | FALSE | 0.504 | 2 | 0.496 | X | X |
| sedative_antipsychotics | 3 | boolean | 0 | FALSE | 0.554 | 2 | 0.446 | | X |
| sedative_antipsychotics | 3 | count | 0 | FALSE | 2.798 | 30 | 0.446 | | |
| sedative_antipsychotics | 7 | boolean | 0 | FALSE | 0.59 | 2 | 0.41 | X | X |

| | | | | | | | | | |
|---|---|---|---|---|---|---|---|---|---|
| sedative_antipsychotics | 7 | count | 0 | FALSE | 5.999 | 65 | 0.41 | X | |
| sedative_antipsychotics | 10 | boolean | 0 | FALSE | 0.606 | 2 | 0.394 | | X |
| sedative_antipsychotics | 10 | count | 0 | FALSE | 8.105 | 89 | 0.394 | | |
| sedative_antipsychotics | 30 | boolean | 0 | FALSE | 0.653 | 2 | 0.347 | X | |
| sedative_antipsychotics | 30 | count | 0 | FALSE | 18.091 | 242 | 0.347 | X | X |
| sedative_antipsychotics | 180 | boolean | 0 | FALSE | 0.713 | 2 | 0.287 | | |
| sedative_antipsychotics | 180 | count | 0 | FALSE | 46.447 | 836 | 0.287 | | X |
| sedative_antipsychotics | 365 | count | 0 | FALSE | 67.507 | 1354 | 0.259 | | X |
| sedative_antipsychotics | 365 | boolean | 0 | FALSE | 0.741 | 2 | 0.259 | | |
| sedative_antipsychotics | 730 | count | 0 | FALSE | 101.64 | 2092 | 0.226 | X | X |
| sedative_antipsychotics | 730 | boolean | 0 | FALSE | 0.774 | 2 | 0.226 | X | |
| sedative_antipsychotics_engangs | 1 | count | 0 | FALSE | 0.012 | 6 | 0.989 | X | X |
| sedative_antipsychotics_engangs | 1 | boolean | 0 | FALSE | 0.011 | 2 | 0.989 | X | X |
| sedative_antipsychotics_engangs | 3 | count | 0 | FALSE | 0.033 | 9 | 0.973 | | X |
| sedative_antipsychotics_engangs | 3 | boolean | 0 | FALSE | 0.027 | 2 | 0.973 | | X |
| sedative_antipsychotics_engangs | 7 | count | 0 | FALSE | 0.069 | 11 | 0.948 | X | X |
| sedative_antipsychotics_engangs | 7 | boolean | 0 | FALSE | 0.052 | 2 | 0.948 | X | X |
| sedative_antipsychotics_engangs | 10 | boolean | 0 | FALSE | 0.067 | 2 | 0.933 | | X |
| sedative_antipsychotics_engangs | 10 | count | 0 | FALSE | 0.092 | 11 | 0.933 | | X |
| sedative_antipsychotics_engangs | 30 | boolean | 0 | FALSE | 0.13 | 2 | 0.87 | X | X |
| sedative_antipsychotics_engangs | 30 | count | 0 | FALSE | 0.202 | 16 | 0.87 | X | X |
| sedative_antipsychotics_engangs | 180 | count | 0 | FALSE | 0.455 | 17 | 0.757 | | X |
| sedative_antipsychotics_engangs | 180 | boolean | 0 | FALSE | 0.243 | 2 | 0.757 | | X |
| sedative_antipsychotics_engangs | 365 | count | 0 | FALSE | 0.628 | 20 | 0.703 | | X |
| sedative_antipsychotics_engangs | 365 | boolean | 0 | FALSE | 0.297 | 2 | 0.703 | | X |
| sedative_antipsychotics_engangs | 730 | boolean | 0 | FALSE | 0.369 | 2 | 0.631 | X | X |
| sedative_antipsychotics_engangs | 730 | count | 0 | FALSE | 0.999 | 43 | 0.631 | X | X |
| sedative_antipsychotics_fast | 1 | boolean | 0 | FALSE | 0.395 | 2 | 0.605 | X | |
| sedative_antipsychotics_fast | 1 | count | 0 | FALSE | 0.606 | 8 | 0.605 | X | |
| sedative_antipsychotics_fast | 3 | boolean | 0 | FALSE | 0.414 | 2 | 0.586 | | |
| sedative_antipsychotics_fast | 3 | count | 0 | FALSE | 1.732 | 22 | 0.586 | | |
| sedative_antipsychotics_fast | 7 | count | 0 | FALSE | 3.72 | 44 | 0.569 | X | X |
| sedative_antipsychotics_fast | 7 | boolean | 0 | FALSE | 0.431 | 2 | 0.569 | X | |
| sedative_antipsychotics_fast | 10 | count | 0 | FALSE | 5.028 | 61 | 0.558 | | X |
| sedative_antipsychotics_fast | 10 | boolean | 0 | FALSE | 0.442 | 2 | 0.558 | | |
| sedative_antipsychotics_fast | 30 | count | 0 | FALSE | 11.215 | 169 | 0.524 | X | X |
| sedative_antipsychotics_fast | 30 | boolean | 0 | FALSE | 0.476 | 2 | 0.524 | X | |
| sedative_antipsychotics_fast | 180 | boolean | 0 | FALSE | 0.535 | 2 | 0.465 | | |
| sedative_antipsychotics_fast | 180 | count | 0 | FALSE | 28.975 | 622 | 0.465 | | X |
| sedative_antipsychotics_fast | 365 | boolean | 0 | FALSE | 0.571 | 2 | 0.429 | | |
| sedative_antipsychotics_fast | 365 | count | 0 | FALSE | 42.258 | 987 | 0.429 | | X |
| sedative_antipsychotics_fast | 730 | count | 0 | FALSE | 64.102 | 1330 | 0.385 | X | X |
| sedative_antipsychotics_fast | 730 | boolean | 0 | FALSE | 0.615 | 2 | 0.385 | X | |
| sedative_antipsychotics_im | 1 | count | 0 | FALSE | 0.004 | 6 | 0.996 | X | X |
| sedative_antipsychotics_im | 1 | boolean | 0 | FALSE | 0.004 | 2 | 0.996 | X | X |
| sedative_antipsychotics_im | 3 | count | 0 | FALSE | 0.012 | 11 | 0.99 | | X |
| sedative_antipsychotics_im | 3 | boolean | 0 | FALSE | 0.01 | 2 | 0.99 | | X |
| sedative_antipsychotics_im | 7 | count | 0 | FALSE | 0.026 | 21 | 0.98 | X | |
| sedative_antipsychotics_im | 7 | boolean | 0 | FALSE | 0.02 | 2 | 0.98 | X | X |
| sedative_antipsychotics_im | 10 | count | 0 | FALSE | 0.035 | 25 | 0.973 | | X |
| sedative_antipsychotics_im | 10 | boolean | 0 | FALSE | 0.027 | 2 | 0.973 | | X |
| sedative_antipsychotics_im | 30 | count | 0 | FALSE | 0.089 | 41 | 0.959 | X | |
| sedative_antipsychotics_im | 30 | boolean | 0 | FALSE | 0.041 | 2 | 0.959 | X | X |
| sedative_antipsychotics_im | 180 | count | 0 | FALSE | 0.305 | 51 | 0.942 | | |
| sedative_antipsychotics_im | 180 | boolean | 0 | FALSE | 0.058 | 2 | 0.942 | | |
| sedative_antipsychotics_im | 365 | boolean | 0 | FALSE | 0.068 | 2 | 0.932 | | X |
| sedative_antipsychotics_im | 365 | count | 0 | FALSE | 0.508 | 61 | 0.932 | | |
| sedative_antipsychotics_im | 730 | boolean | 0 | FALSE | 0.095 | 2 | 0.905 | X | X |
| sedative_antipsychotics_im | 730 | count | 0 | FALSE | 0.926 | 74 | 0.905 | | |
| sedative_antipsychotics_pn | 1 | boolean | 0 | FALSE | 0.224 | 2 | 0.776 | X | X |
| sedative_antipsychotics_pn | 1 | count | 0 | FALSE | 0.363 | 11 | 0.776 | X | X |
| sedative_antipsychotics_pn | 3 | boolean | 0 | FALSE | 0.315 | 2 | 0.685 | | X |
| sedative_antipsychotics_pn | 3 | count | 0 | FALSE | 1.035 | 26 | 0.685 | | X |
| sedative_antipsychotics_pn | 7 | boolean | 0 | FALSE | 0.381 | 2 | 0.619 | X | X |
| sedative_antipsychotics_pn | 7 | count | 0 | FALSE | 2.216 | 51 | 0.619 | X | |
| sedative_antipsychotics_pn | 10 | boolean | 0 | FALSE | 0.408 | 2 | 0.592 | | X |
| sedative_antipsychotics_pn | 10 | count | 0 | FALSE | 2.993 | 68 | 0.592 | | |
| sedative_antipsychotics_pn | 30 | boolean | 0 | FALSE | 0.486 | 2 | 0.514 | X | |
| sedative_antipsychotics_pn | 30 | count | 0 | FALSE | 6.693 | 159 | 0.514 | X | X |
| sedative_antipsychotics_pn | 180 | boolean | 0 | FALSE | 0.579 | 2 | 0.421 | | |
| sedative_antipsychotics_pn | 180 | count | 0 | FALSE | 17.063 | 483 | 0.421 | | X |
| sedative_antipsychotics_pn | 365 | boolean | 0 | FALSE | 0.618 | 2 | 0.382 | | |
| sedative_antipsychotics_pn | 365 | count | 0 | FALSE | 24.689 | 878 | 0.382 | | X |
| sedative_antipsychotics_pn | 730 | boolean | 0 | FALSE | 0.663 | 2 | 0.337 | X | |
| sedative_antipsychotics_pn | 730 | count | 0 | FALSE | 36.648 | 1500 | 0.337 | X | X |
| sedative_antipsychotics_po | 1 | boolean | 0 | FALSE | 0 | 2 | 1 | X | X |

| | | | | | | | | | | |
|---|---|---|---|---|---|---|---|---|---|---|
| sedative_antipsychotics_po | 1 | count | 0 | FALSE | 0 | 3 | 1 | X | | |
| sedative_antipsychotics_po | 3 | count | 0 | FALSE | 0 | 7 | 1 | | | |
| sedative_antipsychotics_po | 3 | boolean | 0 | FALSE | 0 | 2 | 1 | | | |
| sedative_antipsychotics_po | 7 | count | 0 | FALSE | 0 | 12 | 1 | X | | |
| sedative_antipsychotics_po | 7 | boolean | 0 | FALSE | 0 | 2 | 1 | X | | |
| sedative_antipsychotics_po | 10 | boolean | 0 | FALSE | 0 | 2 | 1 | | | |
| sedative_antipsychotics_po | 10 | count | 0 | FALSE | 0 | 15 | 1 | | | |
| sedative_antipsychotics_po | 30 | count | 0 | FALSE | 0.001 | 20 | 1 | X | | |
| sedative_antipsychotics_po | 30 | boolean | 0 | FALSE | 0 | 2 | 1 | X | | |
| sedative_antipsychotics_po | 180 | count | 0 | FALSE | 0.001 | 20 | 1 | | | |
| sedative_antipsychotics_po | 180 | boolean | 0 | FALSE | 0 | 2 | 1 | | | |
| sedative_antipsychotics_po | 365 | boolean | 0 | FALSE | 0 | 2 | 1 | | | |
| sedative_antipsychotics_po | 365 | count | 0 | FALSE | 0.001 | 20 | 1 | | | |
| sedative_antipsychotics_po | 730 | boolean | 0 | FALSE | 0.001 | 2 | 0.999 | X | | |
| sedative_antipsychotics_po | 730 | count | 0 | FALSE | 0.005 | 22 | 0.999 | | X | |
| selvmordsrisiko | 1 | mean | NaN | FALSE | 1.364 | 25 | 0.837 | X | | |
| selvmordsrisiko | 1 | maximum | NaN | FALSE | 1.409 | 4 | 0.837 | X | | |
| selvmordsrisiko | 1 | day | NaN | FALSE | -1.26 | 6014 | 0.96 | | | |
| selvmordsrisiko | 1 | variance | NaN | FALSE | 0.162 | 31 | 0.96 | | | |
| selvmordsrisiko | 1 | minimum | NaN | FALSE | 1.321 | 4 | 0.837 | X | | |
| selvmordsrisiko | 3 | maximum | NaN | FALSE | 1.455 | 4 | 0.68 | | | |
| selvmordsrisiko | 3 | mean | NaN | FALSE | 1.358 | 74 | 0.68 | | | |
| selvmordsrisiko | 3 | day | NaN | FALSE | -0.502 | 26587 | 0.854 | | | |
| selvmordsrisiko | 3 | variance | NaN | FALSE | 0.162 | 167 | 0.854 | | | |
| selvmordsrisiko | 3 | minimum | NaN | FALSE | 1.266 | 4 | 0.68 | | | |
| selvmordsrisiko | 7 | maximum | NaN | FALSE | 1.505 | 4 | 0.508 | X | X | |
| selvmordsrisiko | 7 | mean | NaN | FALSE | 1.356 | 170 | 0.508 | | | |
| selvmordsrisiko | 7 | minimum | NaN | FALSE | 1.222 | 4 | 0.508 | X | | |
| selvmordsrisiko | 7 | variance | NaN | FALSE | 0.163 | 564 | 0.7 | X | | |
| selvmordsrisiko | 7 | day | NaN | FALSE | -0.229 | 44964 | 0.7 | | | |
| selvmordsrisiko | 10 | maximum | NaN | FALSE | 1.531 | 4 | 0.43 | | X | |
| selvmordsrisiko | 10 | mean | NaN | FALSE | 1.356 | 233 | 0.43 | | | |
| selvmordsrisiko | 10 | minimum | NaN | FALSE | 1.202 | 4 | 0.43 | | | |
| selvmordsrisiko | 10 | variance | NaN | FALSE | 0.164 | 888 | 0.62 | | | |
| selvmordsrisiko | 10 | day | NaN | FALSE | -0.196 | 52737 | 0.62 | | | |
| selvmordsrisiko | 30 | mean | NaN | FALSE | 1.349 | 666 | 0.201 | X | X | |
| selvmordsrisiko | 30 | maximum | NaN | FALSE | 1.613 | 4 | 0.201 | X | X | |
| selvmordsrisiko | 30 | minimum | NaN | FALSE | 1.14 | 4 | 0.201 | X | | |
| selvmordsrisiko | 30 | day | NaN | FALSE | -0.139 | 70405 | 0.358 | | | |
| selvmordsrisiko | 30 | variance | NaN | FALSE | 0.171 | 3008 | 0.358 | X | | |
| selvmordsrisiko | 180 | mean | NaN | FALSE | 1.302 | 2249 | 0.039 | | X | |
| selvmordsrisiko | 180 | maximum | NaN | FALSE | 1.709 | 4 | 0.039 | | X | |
| selvmordsrisiko | 180 | minimum | NaN | FALSE | 1.062 | 4 | 0.039 | | | |
| selvmordsrisiko | 180 | variance | NaN | FALSE | 0.166 | 11408 | 0.126 | | X | |
| selvmordsrisiko | 180 | day | NaN | FALSE | -0.024 | 81674 | 0.126 | | | |
| selvmordsrisiko | 365 | maximum | NaN | FALSE | 1.759 | 4 | 0.02 | | X | |
| selvmordsrisiko | 365 | mean | NaN | FALSE | 1.288 | 3712 | 0.02 | | X | |
| selvmordsrisiko | 365 | day | NaN | FALSE | -0.001 | 85275 | 0.083 | | | |
| selvmordsrisiko | 365 | variance | NaN | FALSE | 0.163 | 17158 | 0.083 | | X | |
| selvmordsrisiko | 365 | minimum | NaN | FALSE | 1.046 | 4 | 0.02 | | | |
| selvmordsrisiko | 730 | mean | NaN | FALSE | 1.278 | 5562 | 0.012 | X | X | |
| selvmordsrisiko | 730 | maximum | NaN | FALSE | 1.828 | 4 | 0.012 | X | X | |
| selvmordsrisiko | 730 | variance | NaN | FALSE | 0.164 | 23929 | 0.055 | X | X | |
| selvmordsrisiko | 730 | day | NaN | FALSE | 0.023 | 88549 | 0.055 | | | |
| selvmordsrisiko | 730 | minimum | NaN | FALSE | 1.035 | 4 | 0.012 | | | |
| sex_female | | nan | 0 | TRUE | 0.508 | 2 | 0.492 | X | | X |
| skema_1 | 10 | boolean | 0 | FALSE | 0.025 | 2 | 0.975 | | X | |
| skema_1 | 10 | count | 0 | FALSE | 0.025 | 4 | 0.975 | | X | |
| skema_1 | 10 | summed | 0 | FALSE | 8.528 | 143 | 0.975 | | | |
| skema_1 | 30 | boolean | 0 | FALSE | 0.047 | 2 | 0.953 | X | X | |
| skema_1 | 30 | summed | 0 | FALSE | 17.01 | 145 | 0.953 | X | | |
| skema_1 | 30 | count | 0 | FALSE | 0.049 | 6 | 0.953 | X | X | |
| skema_1 | 180 | count | 0 | FALSE | 0.119 | 12 | 0.903 | | X | |
| skema_1 | 180 | boolean | 0 | FALSE | 0.097 | 2 | 0.903 | | X | |
| skema_1 | 180 | summed | 0 | FALSE | 51.232 | 184 | 0.903 | | | |
| skema_1 | 365 | boolean | 0 | FALSE | 0.125 | 2 | 0.875 | | X | |
| skema_1 | 365 | count | 0 | FALSE | 0.174 | 16 | 0.875 | | X | |
| skema_1 | 365 | summed | 0 | FALSE | 82.235 | 224 | 0.875 | | X | |
| skema_1 | 730 | summed | 0 | FALSE | 160.89 | 291 | 0.826 | X | X | |
| skema_1 | 730 | boolean | 0 | FALSE | 0.174 | 2 | 0.826 | X | X | |
| skema_1 | 730 | count | 0 | FALSE | 0.292 | 21 | 0.826 | X | X | |
| skema_1 | 1 | boolean | 0 | FALSE | 0.087 | 2 | 0.913 | X | X | |
| skema_1 | 3 | boolean | 0 | FALSE | 0.092 | 2 | 0.908 | | X | |
| skema_1 | 7 | count | 0 | FALSE | 0.02 | 4 | 0.98 | X | X | |
| skema_1 | 7 | summed | 0 | FALSE | 6.78 | 143 | 0.98 | X | | |
| skema_1 | 7 | boolean | 0 | FALSE | 0.02 | 2 | 0.98 | X | X | |

| | | | | | | | | | | |
|---|---|---|---|---|---|---|---|---|---|---|
| skema_2_without_nutrition | 10 | boolean | 0 | FALSE | 0.005 | 2 | 0.995 | | | |
| skema_2_without_nutrition | 10 | count | 0 | FALSE | 0.005 | 6 | 0.995 | | | |
| skema_2_without_nutrition | 10 | summed | 0 | FALSE | 3.992 | 120 | 0.995 | | | |
| skema_2_without_nutrition | 30 | count | 0 | FALSE | 0.012 | 7 | 0.989 | X | | |
| skema_2_without_nutrition | 30 | summed | 0 | FALSE | 9.459 | 124 | 0.989 | | | |
| skema_2_without_nutrition | 30 | boolean | 0 | FALSE | 0.011 | 2 | 0.989 | X | | |
| skema_2_without_nutrition | 180 | count | 0 | FALSE | 0.033 | 11 | 0.975 | | | |
| skema_2_without_nutrition | 180 | boolean | 0 | FALSE | 0.025 | 2 | 0.975 | | | |
| skema_2_without_nutrition | 180 | summed | 0 | FALSE | 31.982 | 157 | 0.975 | | | |
| skema_2_without_nutrition | 365 | boolean | 0 | FALSE | 0.033 | 2 | 0.967 | | | |
| skema_2_without_nutrition | 365 | summed | 0 | FALSE | 49.338 | 185 | 0.967 | | | |
| skema_2_without_nutrition | 365 | count | 0 | FALSE | 0.048 | 11 | 0.967 | | | |
| skema_2_without_nutrition | 730 | summed | 0 | FALSE | 82.863 | 223 | 0.953 | X | | |
| skema_2_without_nutrition | 730 | boolean | 0 | FALSE | 0.047 | 2 | 0.953 | X | | |
| skema_2_without_nutrition | 730 | count | 0 | FALSE | 0.08 | 11 | 0.953 | X | | |
| skema_2_without_nutrition | 1 | boolean | 0 | FALSE | 0.027 | 2 | 0.973 | X | X | |
| skema_2_without_nutrition | 3 | boolean | 0 | FALSE | 0.028 | 2 | 0.972 | | X | |
| skema_2_without_nutrition | 7 | count | 0 | FALSE | 0.004 | 5 | 0.996 | X | | |
| skema_2_without_nutrition | 7 | boolean | 0 | FALSE | 0.004 | 2 | 0.996 | X | | |
| skema_2_without_nutrition | 7 | summed | 0 | FALSE | 2.973 | 120 | 0.996 | | | |
| skema_3 | 730 | count | 0 | FALSE | 0.218 | 57 | 0.947 | X | X | |
| skema_3 | 730 | summed | 0 | FALSE | 2.427 | 673 | 0.964 | X | X | |
| skema_3 | 730 | boolean | 0 | FALSE | 0.053 | 2 | 0.947 | X | X | |
| supervised_temporary_leave | 1 | count | 0 | FALSE | 0.038 | 4 | 0.963 | | | |
| supervised_temporary_leave | 1 | boolean | 0 | FALSE | 0.037 | 2 | 0.963 | | | |
| supervised_temporary_leave | 3 | count | 0 | FALSE | 0.109 | 5 | 0.902 | | | |
| supervised_temporary_leave | 3 | boolean | 0 | FALSE | 0.098 | 2 | 0.902 | | | |
| supervised_temporary_leave | 7 | count | 0 | FALSE | 0.23 | 7 | 0.814 | X | | |
| supervised_temporary_leave | 7 | boolean | 0 | FALSE | 0.186 | 2 | 0.814 | X | | |
| supervised_temporary_leave | 10 | boolean | 0 | FALSE | 0.235 | 2 | 0.765 | | | |
| supervised_temporary_leave | 10 | count | 0 | FALSE | 0.305 | 7 | 0.765 | | | |
| supervised_temporary_leave | 30 | boolean | 0 | FALSE | 0.411 | 2 | 0.589 | X | X | |
| supervised_temporary_leave | 30 | count | 0 | FALSE | 0.639 | 11 | 0.589 | X | X | |
| supervised_temporary_leave | 180 | boolean | 0 | FALSE | 0.611 | 2 | 0.389 | | X | |
| supervised_temporary_leave | 180 | count | 0 | FALSE | 1.387 | 21 | 0.389 | | X | |
| supervised_temporary_leave | 365 | count | 0 | FALSE | 1.895 | 31 | 0.331 | | X | |
| supervised_temporary_leave | 365 | boolean | 0 | FALSE | 0.669 | 2 | 0.331 | | X | |
| supervised_temporary_leave | 730 | count | 0 | FALSE | 2.599 | 47 | 0.283 | X | X | |
| supervised_temporary_leave | 730 | boolean | 0 | FALSE | 0.717 | 2 | 0.283 | X | X | |
| temporary_leave | 1 | count | 0 | FALSE | 0.088 | 5 | 0.915 | X | | |
| temporary_leave | 1 | boolean | 0 | FALSE | 0.085 | 2 | 0.915 | X | | |
| temporary_leave | 3 | boolean | 0 | FALSE | 0.204 | 2 | 0.796 | | X | |
| temporary_leave | 3 | count | 0 | FALSE | 0.243 | 6 | 0.796 | | | |
| temporary_leave | 7 | boolean | 0 | FALSE | 0.354 | 2 | 0.646 | X | X | |
| temporary_leave | 7 | count | 0 | FALSE | 0.495 | 9 | 0.646 | X | X | |
| temporary_leave | 10 | boolean | 0 | FALSE | 0.424 | 2 | 0.576 | | X | |
| temporary_leave | 10 | count | 0 | FALSE | 0.649 | 9 | 0.576 | | X | |
| temporary_leave | 30 | count | 0 | FALSE | 1.321 | 14 | 0.364 | X | X | |
| temporary_leave | 30 | boolean | 0 | FALSE | 0.636 | 2 | 0.364 | X | X | |
| temporary_leave | 180 | count | 0 | FALSE | 2.996 | 41 | 0.198 | | X | |
| temporary_leave | 180 | boolean | 0 | FALSE | 0.802 | 2 | 0.198 | | X | |
| temporary_leave | 365 | count | 0 | FALSE | 4.22 | 65 | 0.164 | | X | |
| temporary_leave | 365 | boolean | 0 | FALSE | 0.836 | 2 | 0.164 | | X | |
| temporary_leave | 730 | count | 0 | FALSE | 5.919 | 92 | 0.141 | X | X | |
| temporary_leave | 730 | boolean | 0 | FALSE | 0.859 | 2 | 0.141 | X | X | |
| tvangsindlaeggelse | 10 | summed | 0 | FALSE | 4.555 | 130 | 0.987 | | | |
| tvangsindlaeggelse | 10 | count | 0 | FALSE | 0.013 | 4 | 0.987 | | X | |
| tvangsindlaeggelse | 10 | boolean | 0 | FALSE | 0.013 | 2 | 0.987 | | X | |
| tvangsindlaeggelse | 30 | boolean | 0 | FALSE | 0.026 | 2 | 0.974 | | X | |
| tvangsindlaeggelse | 30 | count | 0 | FALSE | 0.026 | 6 | 0.974 | | X | |
| tvangsindlaeggelse | 30 | summed | 0 | FALSE | 9.244 | 131 | 0.974 | X | | |
| tvangsindlaeggelse | 180 | count | 0 | FALSE | 0.068 | 11 | 0.942 | | X | |
| tvangsindlaeggelse | 180 | boolean | 0 | FALSE | 0.058 | 2 | 0.942 | | X | |
| tvangsindlaeggelse | 180 | summed | 0 | FALSE | 30.517 | 164 | 0.942 | | | |
| tvangsindlaeggelse | 365 | boolean | 0 | FALSE | 0.076 | 2 | 0.924 | | X | |
| tvangsindlaeggelse | 365 | summed | 0 | FALSE | 49.005 | 194 | 0.924 | | X | |
| tvangsindlaeggelse | 365 | count | 0 | FALSE | 0.101 | 15 | 0.924 | | X | |
| tvangsindlaeggelse | 730 | boolean | 0 | FALSE | 0.105 | 2 | 0.895 | X | X | |
| tvangsindlaeggelse | 730 | summed | 0 | FALSE | 91.661 | 246 | 0.895 | | X | |
| tvangsindlaeggelse | 730 | count | 0 | FALSE | 0.165 | 21 | 0.895 | X | X | |
| tvangsindlaeggelse | 1 | boolean | 0 | FALSE | 0.05 | 2 | 0.95 | X | X | X |
| tvangsindlaeggelse | 3 | boolean | 0 | FALSE | 0.053 | 2 | 0.947 | | X | |
| tvangsindlaeggelse | 7 | boolean | 0 | FALSE | 0.011 | 2 | 0.989 | X | X | |
| tvangsindlaeggelse | 7 | summed | 0 | FALSE | 3.664 | 130 | 0.989 | | | |
| tvangsindlaeggelse | 7 | count | 0 | FALSE | 0.011 | 4 | 0.989 | X | X | |
| tvangstilbageholdelse | 10 | boolean | 0 | FALSE | 0.012 | 2 | 0.988 | | | |

| | | | | | | | | | | |
|---|---|---|---|---|---|---|---|---|---|---|
| tvangstilbageholdelse | 10 | summed | 0 | FALSE | 3.974 | 101 | 0.988 | | | |
| tvangstilbageholdelse | 10 | count | 0 | FALSE | 0.012 | 4 | 0.988 | | | |
| tvangstilbageholdelse | 30 | count | 0 | FALSE | 0.023 | 4 | 0.978 | X | | |
| tvangstilbageholdelse | 30 | summed | 0 | FALSE | 7.766 | 101 | 0.978 | X | | |
| tvangstilbageholdelse | 30 | boolean | 0 | FALSE | 0.022 | 2 | 0.978 | X | | |
| tvangstilbageholdelse | 180 | count | 0 | FALSE | 0.051 | 5 | 0.954 | | | |
| tvangstilbageholdelse | 180 | boolean | 0 | FALSE | 0.046 | 2 | 0.954 | | | |
| tvangstilbageholdelse | 180 | summed | 0 | FALSE | 20.715 | 110 | 0.954 | | | |
| tvangstilbageholdelse | 365 | summed | 0 | FALSE | 33.229 | 129 | 0.938 | | | |
| tvangstilbageholdelse | 365 | boolean | 0 | FALSE | 0.062 | 2 | 0.938 | | X | |
| tvangstilbageholdelse | 365 | count | 0 | FALSE | 0.073 | 7 | 0.938 | | X | |
| tvangstilbageholdelse | 730 | boolean | 0 | FALSE | 0.096 | 2 | 0.904 | X | X | |
| tvangstilbageholdelse | 730 | summed | 0 | FALSE | 69.229 | 170 | 0.904 | X | X | |
| tvangstilbageholdelse | 730 | count | 0 | FALSE | 0.127 | 11 | 0.904 | X | X | |
| tvangstilbageholdelse | 1 | boolean | 0 | FALSE | 0.036 | 2 | 0.964 | X | X | X |
| tvangstilbageholdelse | 3 | boolean | 0 | FALSE | 0.039 | 2 | 0.961 | | X | |
| tvangstilbageholdelse | 7 | summed | 0 | FALSE | 3.116 | 101 | 0.991 | X | | |
| tvangstilbageholdelse | 7 | boolean | 0 | FALSE | 0.009 | 2 | 0.991 | X | | |
| tvangstilbageholdelse | 7 | count | 0 | FALSE | 0.009 | 4 | 0.991 | X | | |
| unsupervised_temporary_leave | 1 | boolean | 0 | FALSE | 0.048 | 2 | 0.952 | X | | |
| unsupervised_temporary_leave | 1 | count | 0 | FALSE | 0.049 | 4 | 0.952 | X | | |
| unsupervised_temporary_leave | 3 | count | 0 | FALSE | 0.134 | 6 | 0.882 | | X | |
| unsupervised_temporary_leave | 3 | boolean | 0 | FALSE | 0.118 | 2 | 0.882 | | X | |
| unsupervised_temporary_leave | 7 | count | 0 | FALSE | 0.266 | 8 | 0.792 | X | X | |
| unsupervised_temporary_leave | 7 | boolean | 0 | FALSE | 0.208 | 2 | 0.792 | X | X | |
| unsupervised_temporary_leave | 10 | boolean | 0 | FALSE | 0.253 | 2 | 0.747 | | X | |
| unsupervised_temporary_leave | 10 | count | 0 | FALSE | 0.344 | 9 | 0.747 | | X | |
| unsupervised_temporary_leave | 30 | count | 0 | FALSE | 0.682 | 14 | 0.6 | X | X | |
| unsupervised_temporary_leave | 30 | boolean | 0 | FALSE | 0.4 | 2 | 0.6 | X | X | |
| unsupervised_temporary_leave | 180 | boolean | 0 | FALSE | 0.56 | 2 | 0.44 | | X | |
| unsupervised_temporary_leave | 180 | count | 0 | FALSE | 1.609 | 39 | 0.44 | | X | |
| unsupervised_temporary_leave | 365 | count | 0 | FALSE | 2.325 | 61 | 0.386 | | X | |
| unsupervised_temporary_leave | 365 | boolean | 0 | FALSE | 0.614 | 2 | 0.386 | | X | |
| unsupervised_temporary_leave | 730 | boolean | 0 | FALSE | 0.659 | 2 | 0.341 | X | X | |
| unsupervised_temporary_leave | 730 | count | 0 | FALSE | 3.321 | 89 | 0.341 | X | X | |
| weight_in_kg | 10 | latest | NaN | FALSE | 79.34 | 2042 | 0.514 | | | |
| weight_in_kg | 30 | latest | NaN | FALSE | 80.315 | 2050 | 0.315 | X | X | |
| weight_in_kg | 365 | latest | NaN | FALSE | 81.398 | 2077 | 0.104 | | X | |
| weight_in_kg | 730 | latest | NaN | FALSE | 81.457 | 2081 | 0.069 | X | X | |
| weight_in_kg | 180 | latest | NaN | FALSE | 81.233 | 2070 | 0.152 | | X | |

**Supplementary table 3: Description of clinical note (text) content by type.**

| Danish name | English name | Description |
|---|---|---|
| Aftaler, Psykiatri | Appointments, Psychiatry | Description of concrete care- and treatment-related appointments and agreements with and about the patient. |
| Aktuelt socialt, Psykiatri | Current social functioning | Description of the patient's current social situation, including relationship to family, civil status, residential-, occupational-, and economic conditions, and contact with the social services. Documentation of ongoing treatment plans in relation to the patient's social relationships. |
| Aktuelt psykisk | Subjective mental state | Description of the development, progress, and current status of the patient's mental illness. |
| Aktuelt somatisk, Psykiatri | Subjective physical state | Description of the patient's current and chronic somatic illnesses and symptoms. Information on current treatment in relation to the patient's physical condition. |
| Konklusion/vurdering | Conclusion/evaluation | Aggregation and interpretation of all findings and decisions based on an interview (e.g., in connection with an outpatient visit or during an inpatient stay). |
| Kontaktårsag | Reason for contact | The reason for the patient's in- or outpatient contact. |
| Objektivt psykisk | Objective mental state | Objective assessment of the patient's mental state, including state of |

| | | |
|---|---|---|
| | | consciousness, orientation, intelligence, psychomotor function, mood, delusions, psychotic symptoms, etc. |
| Observation af patient, Psykiatri | Observation of patient, Psychiatry | Description of an inpatient's mental symptoms, behaviour, reactions towards relatives, other patients, staff, etc. |
| Samtale med behandlingssigte | Conversation with aim of treatment | Documentation of the conversation's purpose and attendees. |
| Semistruktureret diagnostisk interview | Semi-structured diagnostic interview | Registration of semi-structured diagnostic interview, including a description of the method and documentation of result and conclusion. |
| Telefonnotat | Telephone note | Documentation of telephone conversations with a patient or the patient's guardian of nonclinical character. |

Adapted from Hansen et al. (2).

**Supplementary table 4: Hyperparameter search space**

| Hyperparameter search space | |
|---|---|
| **Alpha** | [1e-8-0.1] |
| **Lambda** | [1e-8-1] |
| **Gamma** | [1e-8-0.001] |
| **Learning rate** | [1e-8-0.1] |
| **Max depth** | [3-8] |
| **Number of estimators** | [100-1500] |
| **Scaling** | [Standardisation, no scaling] |
| **Imputation** | [Most frequent value imputation, mean value imputation] |
| **Predictor set** | [Structured predictors only, structured + text predictors] |
| **Predictor selection** | [ANOVA F-test univariate feature selection, no predictor selection] |
| **Predictor selection percentage** | [5%, 10%, 15%, 20%, 40%, 60%, 80%] |
| **Lookbehind combination** | [(1), (1, 3), (730), (1, 3, 10), (1, 7, 30, 730), (1, 3, 7, 10, 30, 180, 365, 730)] |

**Supplementary table 5: Optimal hyperparameters**

| Optimal hyperparameters | | |
|---|---|---|
| | **Mechanical restraint model** | **Composite restraint model** |
| **Alpha** | 0.0001286683 | 0.0000078439 |
| **Lambda** | 0.0384306469 | 0.0000075616 |
| **Gamma** | 0.0000000459 | 0.0000000835 |
| **Learning rate** | 0.0207201727 | 0.0397284173 |
| **Max depth** | 5 | 3 |
| **Number of estimators** | 409 | 357 |
| **Scaling** | No scaling | Standardisation |
| **Imputation** | Mean value imputation | Mean value imputation |
| **Predictor set** | Structured predictors only | Structured + text predictors |
| **Predictor selection** | ANOVA F-test univariate feature selection | ANOVA F-test univariate feature selection |
| **Predictor selection percentage** | 80% | 40% |
| **Lookbehind combination** | (1, 7, 30, 730) | (1, 3, 7, 10, 30, 180, 365, 730) |

**Supplementary table 6: Performance estimates for models on the training data**

| | **Mechanical restraint model** | **Composite restraint model** |
|---|---|---|
| **Area under the operator receiver characteristic** | 0.887 (SD = 0.009) | 0.894 (SD = 0.007) |

Mean area under the receiver operating characteristic across five cross-validation folds.

**Supplementary figure 1: Text vectorisation**

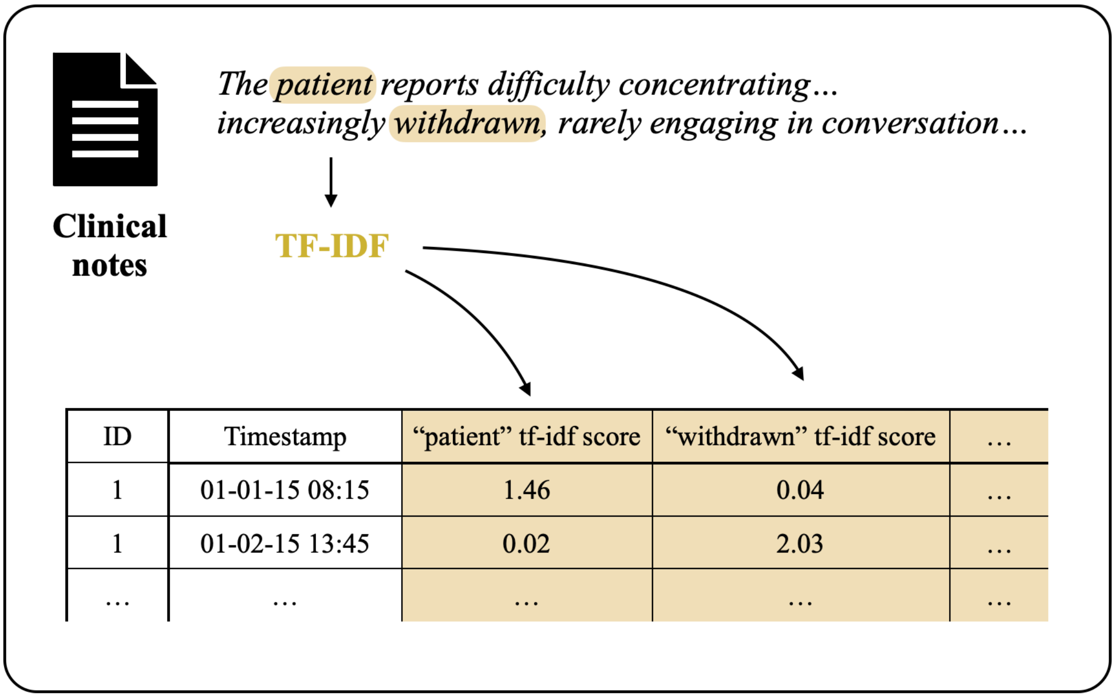

Clinical notes were vectorised using term frequency-inverse document frequency (TF-IDF).

**Supplementary figure 2: Performance across demographic characteristics**

A. Mechanical restraint model validated on mechanical restraint

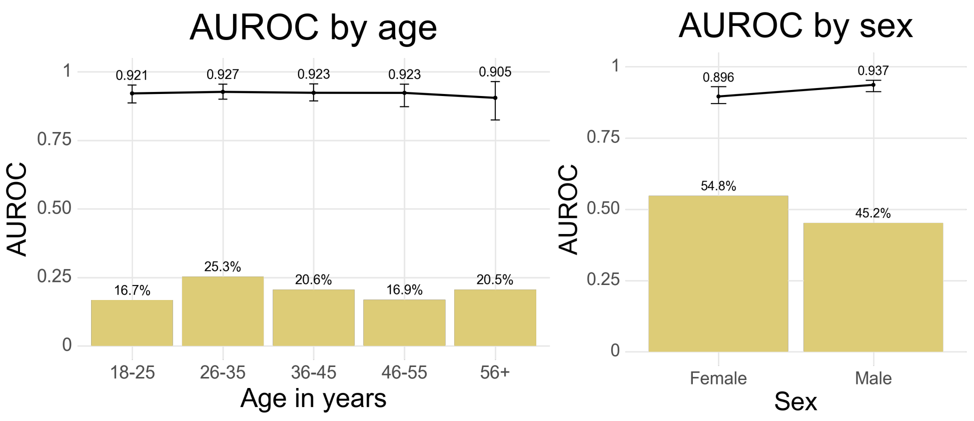

B. Composite restraint model validated on mechanical restraint

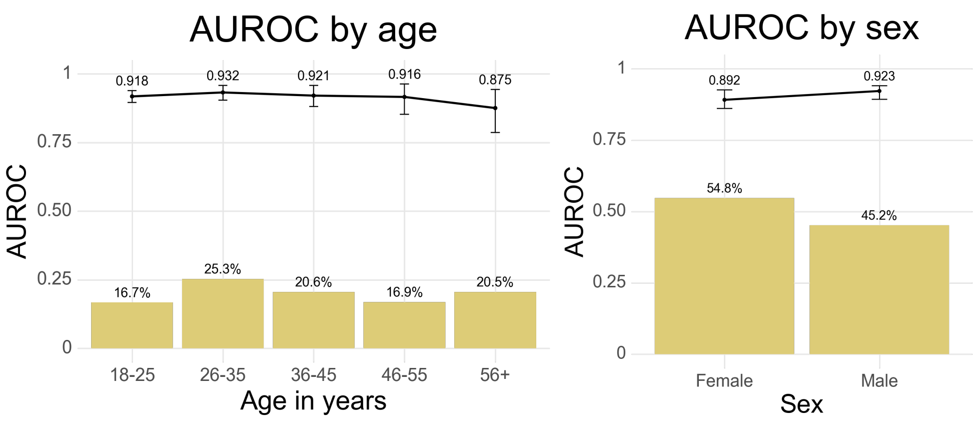

C. Composite restraint model validated on composite restraint

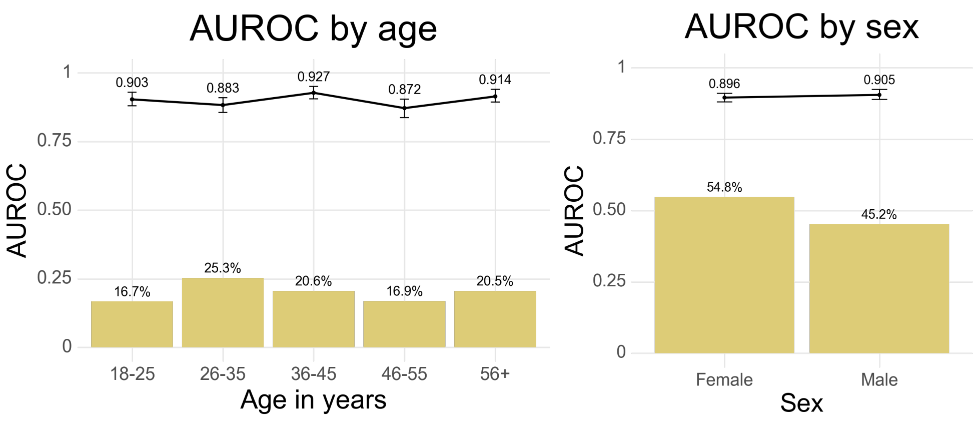

Area under the operating receiver characteristic (AUROC) performance stratified by sex and age groups in the test set. Bar heights indicate group proportion. Error bars denote 95% confidence intervals based on 100 bootstrap samples.

**Supplementary figure 3: Performance across temporal characteristics**

A. Mechanical restraint model validated on mechanical restraint

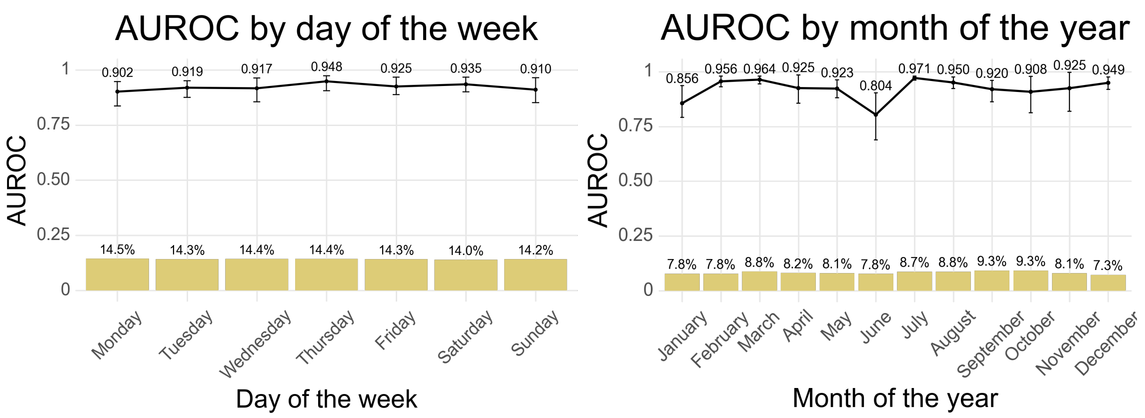

B. Composite restraint model validated on mechanical restraint

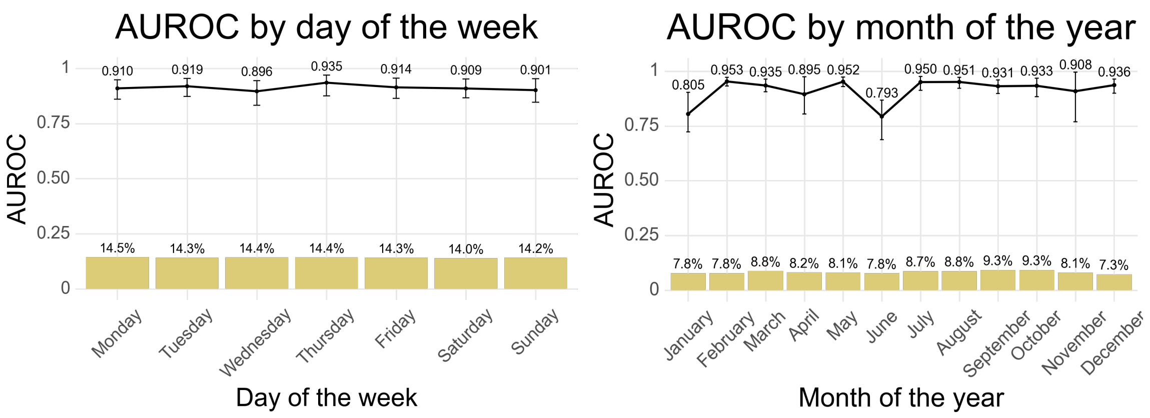

C. Composite restraint model validated on composite restraint

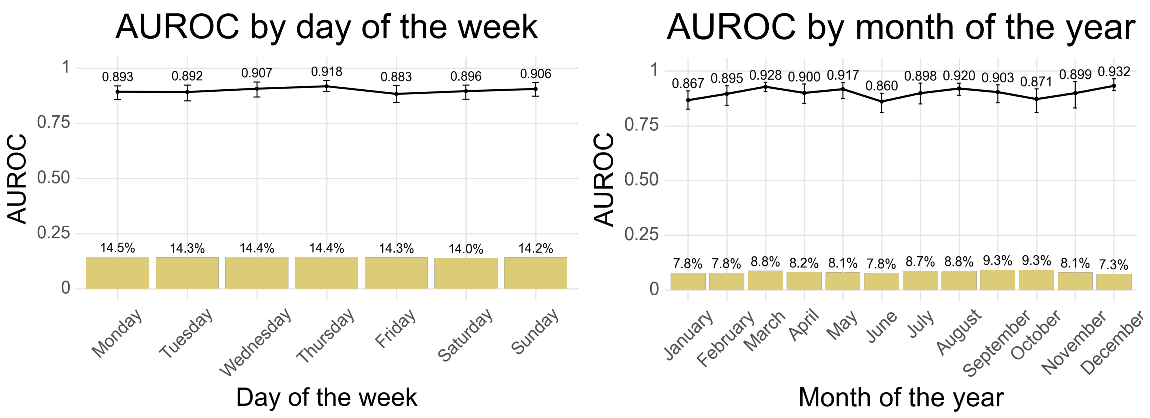

Area under the operating receiver characteristic (AUROC) performance stratified by days of the week and months of year in the test set. Bar heights indicate group proportion. Error bars denote 95% confidence intervals based on 100 bootstrap samples.

**Supplementary figure 4: Performance across geographical location**

A. Mechanical restraint model validated on mechanical restraint

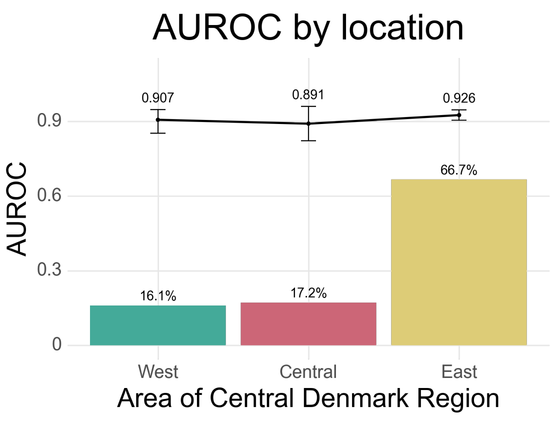

B. Composite restraint model validated on mechanical restraint

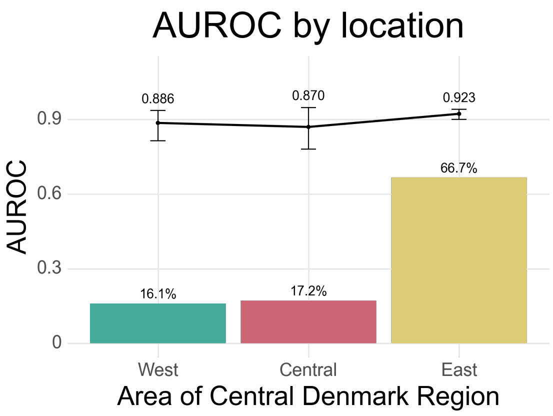

C. Composite restraint model validated on composite restraint

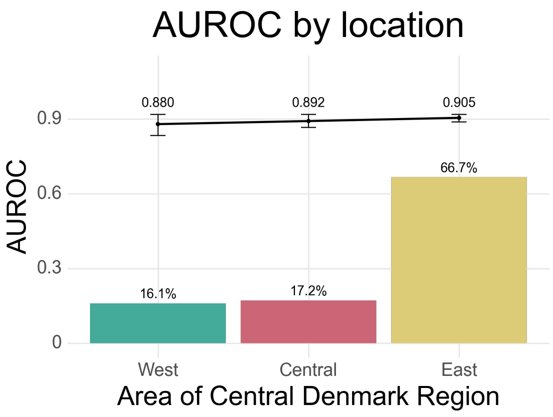

Area under the operating receiver characteristic (AUROC) performance stratified geographic location of admissions. Bar heights indicate group proportion. Error bars denote 95% confidence intervals based on 100 bootstrap samples.

**Supplementary figure 5: Temporal stability**

A. Mechanical restraint model validated on mechanical restraint

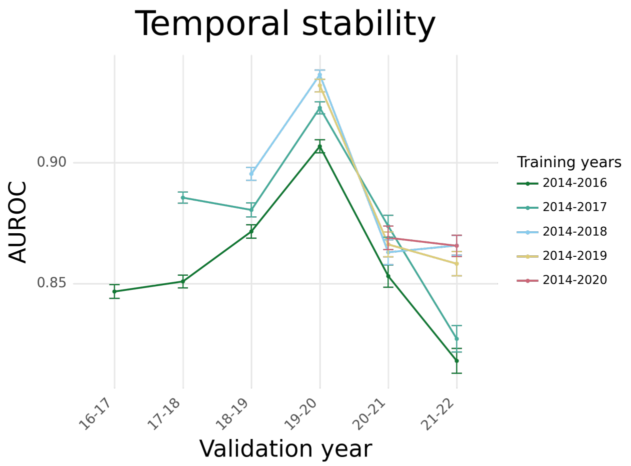

B. Composite restraint model validated on mechanical restraint

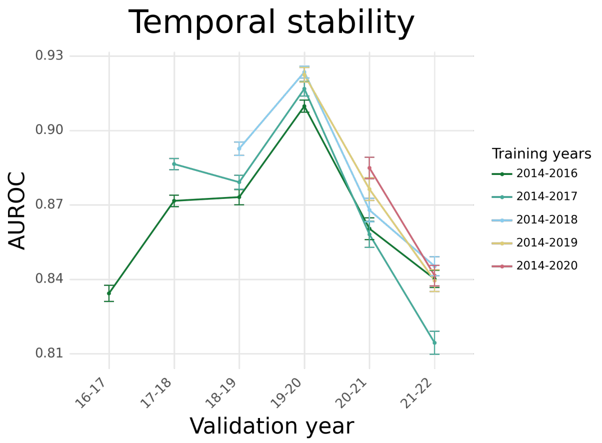

C. Composite restraint model validated on composite restraint

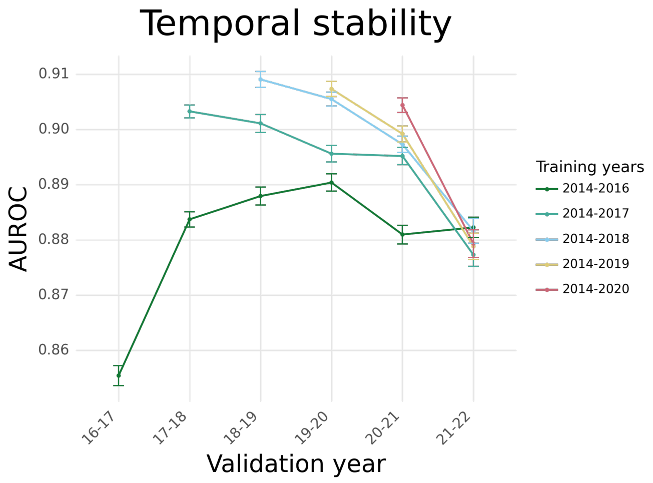

**Supplementary figure 6: Sensitivity across outcome types**

A. Mechanical restraint model validated on mechanical restraint

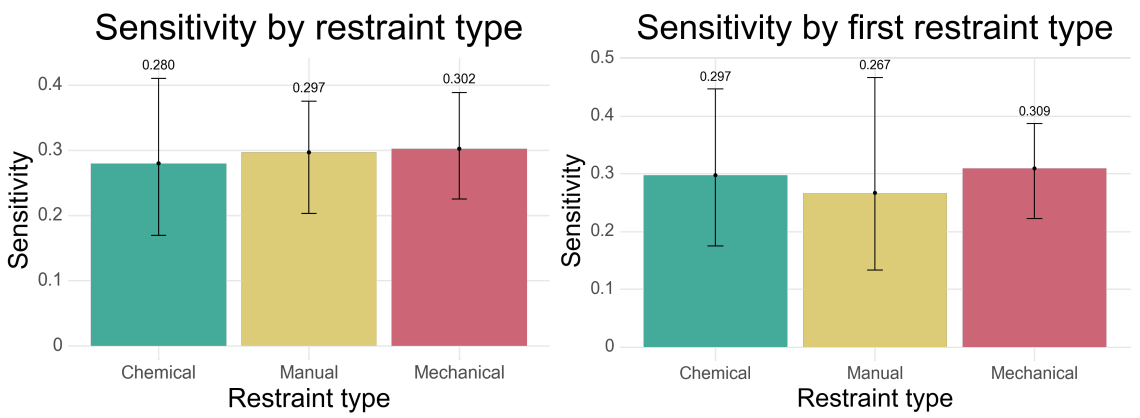

B. Composite restraint model validated on mechanical restraint

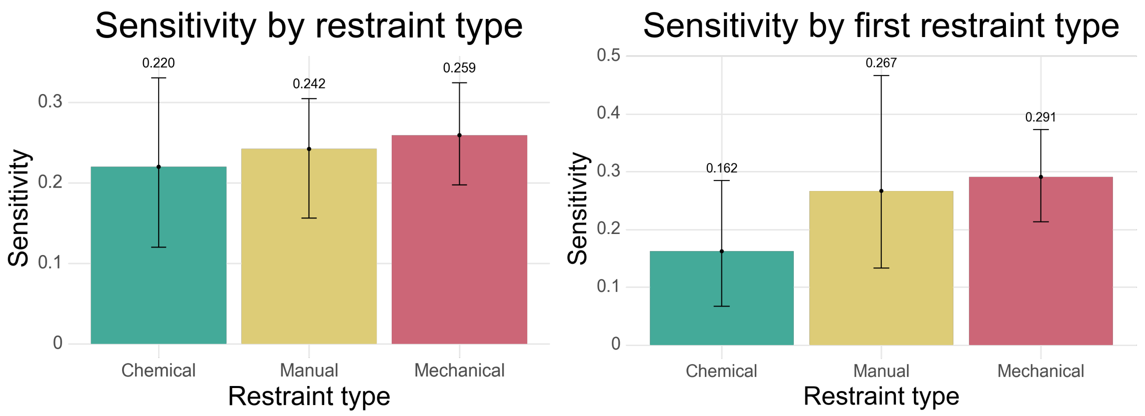

C. Composite restraint model validated on composite restraint

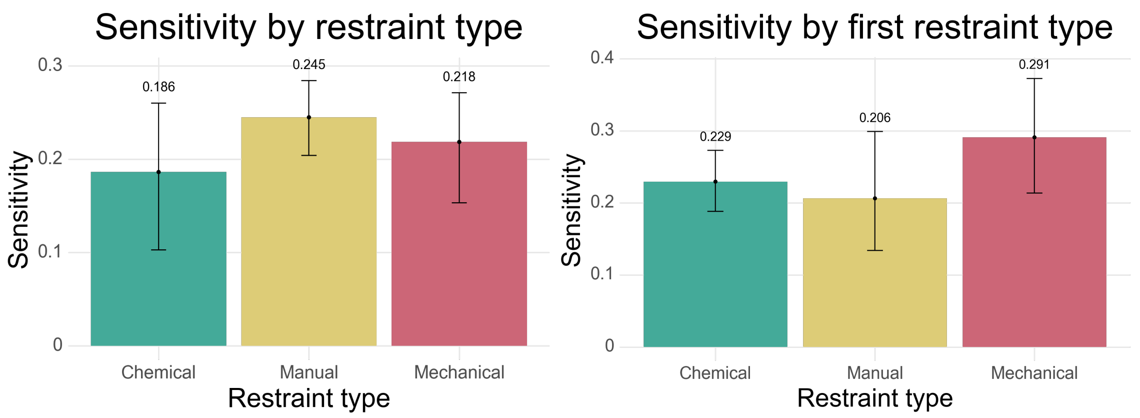

Sensitivity stratified by restraint type and index restraint type (first type recorded if multiple types occur within the lookahead period). Top 1% highest risk predictions classified as

positive predictions. Bar heights indicate group proportion. Error bars denote 95% confidence intervals based on 100 bootstrap samples.
